# Can longitudinal electronic health record data identify patients at higher risk of developing long COVID?

**DOI:** 10.1101/2024.02.08.24302528

**Authors:** Priya Shanmugam, Molly Bair, Emma Pendl-Robinson, Xindi C. Hu, N3C consortium

## Abstract

With hundreds of millions of COVID-19 infections to date, a considerable portion of the population has developed or will develop long COVID. Understanding the prevalence, risk factors, and healthcare costs of long COVID can be of significant societal importance. To investigate the utility of large-scale electronic health record (EHR) data in identifying and predicting long COVID, we analyzed a sample of 1.23 million participants from the National COVID Cohort Collaborative (N3C), a longitudinal EHR data repository from 80 sites in the US with over 8 million COVID-19 patients. We characterized the prevalence of long COVID using a few different types of definitions to illustrate their relative strengths and weaknesses. Then we developed machine learning models to predict the risk of developing long COVID using demographic factors and comorbidity in the EHR. The risk factors for long COVID include patient age; sex; smoking status; and comorbidities characterized by the Charlson Comorbidity Index (CCI). We were able to predict three types of long COVID with low to moderate levels of accuracy (AUC 0.599 – 0.734). We found that age and CCI were most predictive of long COVID diagnosis. Ongoing work includes applying the fair machine learning framework to the long COVID predictive models. We are implementing fairness and bias mitigation methods to model fitting through the following steps, selecting fairness metrics, preparing data and model, evaluating fairness metrics, applying bias mitigation methods to the dataset, and comparing model results and fairness metrics before and after the mitigation. The objective is to achieve equalized odds, a statistical notion that ensures classification algorithms do not discriminate against protected groups (such as sex and race/ethnicity). Results from the fairness-based machine learning will be included in the conference presentation.

## 1. Introduction

By November 2022, 94% of the US population was estimated to have been infected by SARS-CoV-2 at least once (Klaassen et al., 2022). This proportion continues to rise as COVID-19 remains as an important public health threat. Some people recover quickly after the initial infection, while others (between 10% to 30%) continue to experience symptoms even months after the initial infection (Herman et al., 2022). Long COVID is defined by the World Health Organization (WHO) as persistent symptoms and/or long-term complications following a probable or confirmed SARS-CoV-2 infection, usually 3 months from acute infection and lasting longer than 2 months with no probable alternative diagnosis (Nittas et al., 2022; World Health Organization, 2021). However, many challenges exist around diagnosing and managing patients with long COVID and identifying individuals who may have elevated risks of developing long COVID. Partially because the long-term effects of COVID-19 may exhibit differently for different people, or because tracking symptoms that can wax and wane over a long period of time can be difficult. Here we leverage a longitudinal electronic health record (EHR) data repository to test a few definitions of long COVID and develop a risk prediction model to identify patients at higher risks of developing long COVID. With hundreds of millions of COVID-19 infections to date, a considerable portion of the population has developed or will develop long COVID. Understanding the prevalence, risk factors, and healthcare costs of long COVID can be of significant societal importance. The healthcare sector needs to better understand the demand for medical care due to long COVID and be prepared to deliver that. Public health sector needs to be able to identify population segments at elevated risks for long COVID in order to design and implement targeted intervention strategies.

More than 80 symptoms have been identified in the literature to be potentially associated with long COVID (Nasserie et al., 2021). Like the initial SARS-CoV-2 infection, long COVID could affect multiple organs and body systems. The most common symptoms include fatigue, dyspnea, cardiac abnormalities, cognitive impairment, sleep disturbances, symptoms of post-traumatic stress disorder, muscle pain, concentration problems, and headache (Crook et al., 2021). In addition to a long list of symptoms, information on symptom severity, duration, and co-occurrence is generally lacking in the literature. Several approaches exist among published studies to characterize long COVID: some uses a very liberal definition of having at least one symptom which result in high percentage of patients estimated to have long COVID but is likely an overestimate (CDC, 2023), some focuses on the most salient symptoms like fatigue, headache, dyspnea and anosmia (Sudre et al., 2021), some focuses on symptom clusters to capture the overlapping symptoms (Global Burden of Disease Long COVID Collaborators, 2022). A wide range of prevalence estimates for long COVID may be a result of non-standardized definition, virulence of different SARS-CoV-2 variants, and population immunity. Prevalence of as high as 80% was observed in early studies, using a lenient definition of at least one symptom, among patients who were previously hospitalized (Nasserie et al., 2021). The Centers of Disease Control and Prevention (CDC) estimated that nearly one in five American adults who have had COVID-19 still have long COVID (CDC, 2022). More recently, the Global Burden of Disease Long COVID Collaborators estimated that 6.2% individuals have at least one of the three select-reported long COVID symptom clusters (Global Burden of Disease Long COVID Collaborators, 2022). As a result of the uncertainty around the prevalence of long COVID, the economic cost of long COVID is hard to fully grasp, but safe to assume it would be an enormous number. Cutler and Summers estimated the economic costs of long COVID to be $2.6 trillion in 2020, and later updated it to be $3.7 trillion as a result of higher prevalence of long COVID than previously assumed (D. Cutler, 2022; D. M. Cutler & Summers, 2020).

The enormity of the economic costs and societal impact of long COVID highlights the necessity to develop better methods for detecting, treating, and preventing long COVID. In this study, we analyzed data from the National COVID Cohort Collaborative (N3C), a longitudinal EHR data repository from 80 sites in the US with over 8 million COVID-19 patients (Haendel et al., 2021). We characterize the prevalence of long COVID using a few different types of definitions to illustrate their relative strengths and weaknesses. Then we develop machine learning models to predict the risk of developing long COVID using demographic factors and comorbidity in the EHR. We discuss model performance with a focus on features with strong predictive power that can be utilized to design early detection or targeted intervention strategies. We the discuss how EHR data derived risk prediction model can be used to enhance an individual COVID-19 risk calculator, 19andMe, to support individual and clinical decision-making. Ongoing work also includes assessing algorithmic bias and mitigating them using techniques including resampling and reweighting.

## 2. Our approach to predicting long COVID

Several features of long COVID make identifying positive cases using claims and EHR data challenging. First, some symptoms of long COVID may or may not be detectable, depending on which diagnostic test is used (Mancini et al., 2021). Second, due to early skepticism within the medical profession about the existence of COVID-19, there have been significant informal, patient-led efforts to report and track the various symptoms of long COVID (Patient Led Research Collaborative, 2023). Third, symptoms occurring during the long COVID episode must be separated from the initial episode of COVID itself, which may vary in length and severity. Fourth, like other medical conditions, a claims-based definition must be clinically relevant with respect to disease severity, and account for health conditions a patient may have had prior to their COVID and long COVID episodes.

Given these challenges, we tested several approaches to defining long COVID using data from the N3C Enclave. We first defined a six-month lookback & look-forward window around each patients’ index COVID-19 episode and limited our sample to patients who have healthcare utilization (defined as having a condition recorded in the N3C database) prior to, and after, these windows. This allowed us to accurately classify a lack of healthcare utilization during the lookback or look-forward windows as true non-utilization, as opposed to a patient death, relocation, or otherwise missing data. We then identified all OMOP concept ID condition codes that appear on a patient’s record between 90 and 180 days following their index COVID-19 diagnosis. Excluding conditions recorded within 90 days after an initial COVID-19 diagnosis ensured that we do not flag symptoms of COVID-19 as symptoms of long COVID. From the resulting set of symptoms, we removed any symptoms that appeared on a patient’s record during their lookback window. A drawback of this approach is that it reduces the likelihood that we detect long COVID cases among people with chronic conditions or comorbidities, who are actually most likely to contract long COVID. We additionally removed any symptoms during the look-forward period that do not appear on the list of 76 conditions in the N3C Long COVID OMOP condition code set. The remaining symptoms were considered as the patient’s long COVID symptom burden.

We considered that the N3C long COVID OMOP condition code set contains several potential redundancies, and that these conditions might be over-reported in claims and EHR data due to upcoding. For example, the following pairs of symptoms were similar to one another, yet each appeared as an independent item in the OMOP code set: sleep disturbances and insomnia, nausea and vomiting, body aches and muscle pain, and fatigue and exercise intolerance. We consolidated these redundancies in a patient’s look-forward period prior to calculating each patient’s long COVID symptom score. This reduced the number of long covid symptoms from 76 to 72.

### 3 symptom cluster approach

Lastly, we built off of the recent literature by calculating a cluster-specific score for each of three symptom clusters (Cognitive, Respiratory, and Fatigue) (Global Burden of Disease Long COVID Collaborators, 2022). We created three cluster-specific, binary long COVID definitions based on a cutoff of at least 3 distinct long COVID symptoms within the relevant cluster. This approach provided a few advantages: it built off the clinical literature identifying these three symptom clusters as distinct, allowing us to classify each patient into each subtype without concern for whether a patient exhibits symptoms from multiple clusters. Second, it allowed us to capture variation in the relationship between patient demographics and various long COVID subtypes. Third, it allowed us to draw distinctions between more and less severe symptom clusters of long COVID, and potentially identify predictive targets that capture high-morbidity or high-cost events.

## 3. Characteristics of Sample

We used the N3C de-identified “tier 2 access” data set.^1^ A limitation of our analysis is that the Patient Severity and Scores dataset, condition occurrence, and drug exposure datasets are frequently updated within the N3C data enclave and new records being added. Our analysis used the Patient Severity and Scores dataset version 70 which was released on March 19, 2022. Thus, all patients in our analysis were diagnosed on or before March 19, 2022. Therefore, our results did not capture recent changes in the COVID-19 pandemic including the rise in the prevalence of the omicron variant, vaccination, and vaccine booster doses.

For our analysis, we started with 13,151,716 patients from the Patient Severity and Scores database and filtered down to the patients with lab confirmed positive cases (3,724,542 patients). Finally, we selected the patients who had conditions occurrences within the 6 months preceding and proceeding diagnosis of COVID-19,^2^ whose COVID-19 visit start date was between September 1, 2020, and September 1, 2021, and who did not die from COVID-19. This provided us with our final analytic sample of 1,234,119 patients.

We selected covariates based on our literature review of covariates other researchers found were related to long COVID symptoms (Tsampasian et al., 2023). We identified the following covariates using the Patient Severity and Scores dataset: age, sex, race, ethnicity, and current or former smoking status. We categorized the patient’s age at diagnosis into 9 categories (0-9, 10-19, 20-29, 30-39, 40-49, 50-59, 60-69, 70-79, and 80+ years old) for patients missing age, we imputed age as the median age 41.2 which translates to the age category 40-49. We categorized sex as male, female, or other. Race was defined as White, Black or African American, Asian, Native Hawaiian or Other Pacific Islander, Other, and Missing/Unknown. Ethnicity was defined as Hispanic or Latino, Not Hispanic or Latino, and Ethnicity Missing or Unknown.

In addition, we identified the following covariates using the OMOP code set ids in the condition occurrence and drug exposure datasets for the dates prior to the covid diagnosis: COVID-19 vaccination, pregnancy, hypertension, obesity, immunocompromised, and Charlson Comorbidity Index (CCI) score (Glasheen et al., 2019). For each patient, if the OMOP codes for the comorbidity’s hypertension, obesity, immunocompromised, or CCI score appear in the condition occurrence data before the COVID-19 diagnosis, then we categorized the patient as having the respective comorbidity. We considered a patient vaccinated if a patient received one or more doses of a Pfizer, Moderna, or Johnson and Johnson vaccine before diagnosis of COVID-19 as recorded in the drug exposure dataset. We considered a patient pregnant if the condition start date was within a year of COVID-19 diagnosis. See Appendix A1 for full list of the OMOP codes we used to create the covariates.

Our sample was 56% (n = 692,547) female, 68% (n = 1,031,221) White, 2% (n = 25,083) received at least one dose of a COVID-19 vaccine, 22% (n = 27,3255) were 60 years old or older, see Table 1 for additional attributes of the analytic sample.

**Table 1:**
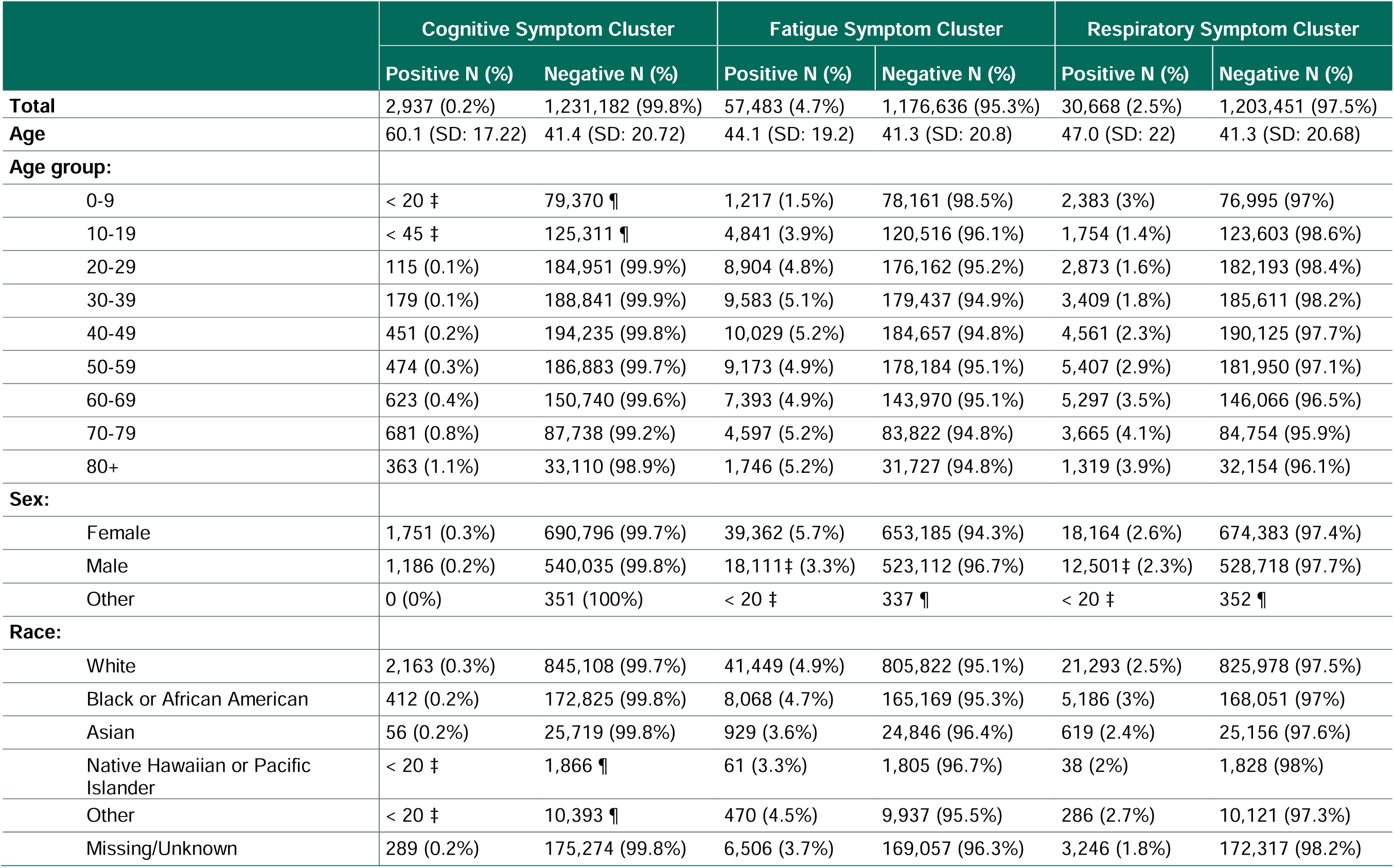
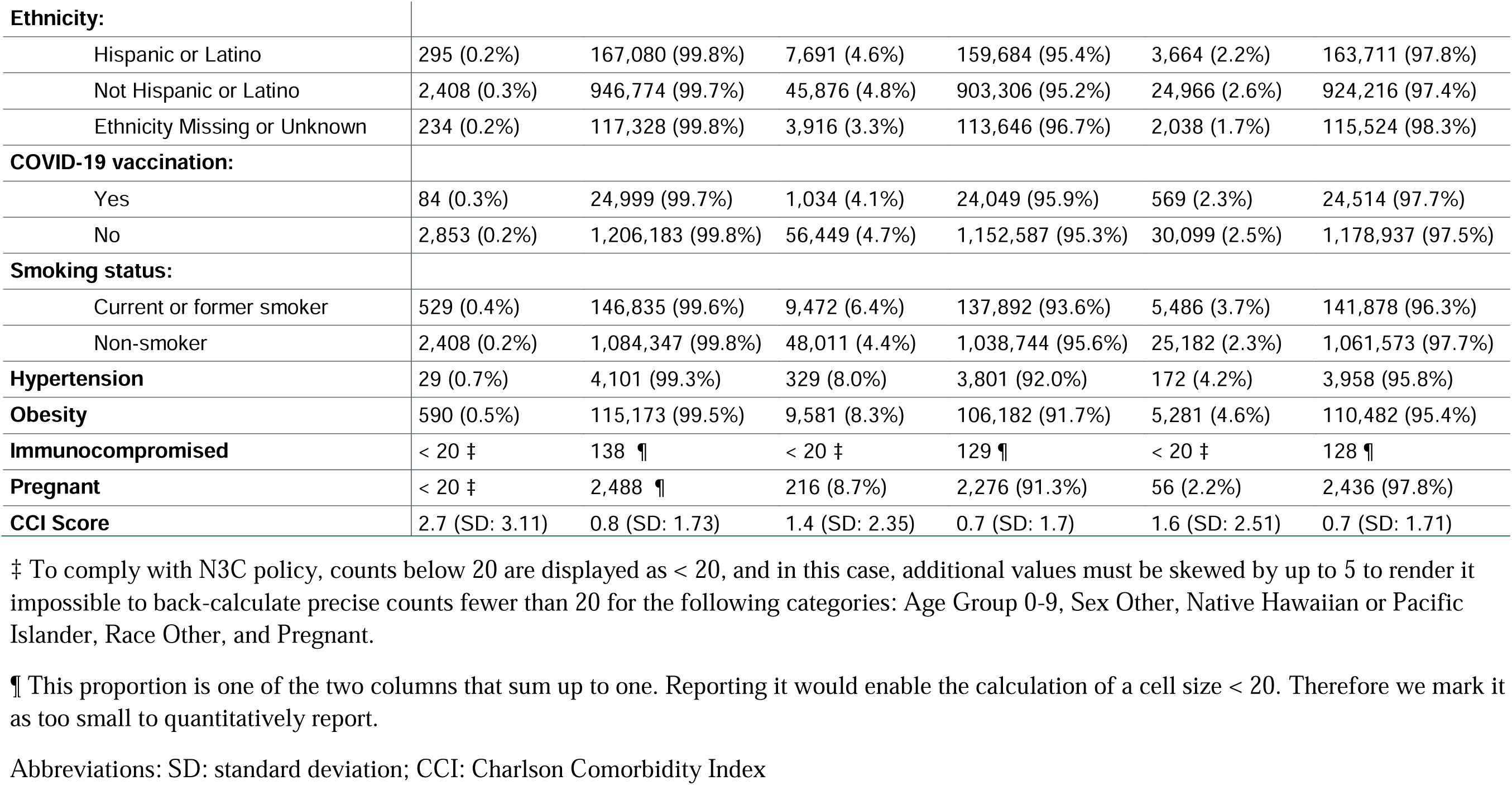
Analytic Sample.

Several limitations exist in our data. First, the observed conditions dataset only recorded diagnoses, signs, or symptoms of a condition either observed by a provider or reported by the patient (Blacketer, 2021). This limitation might have affected the makeup of the sample through: 1) underreporting conditions because patients might have had conditions treated at another facility not sharing data with N3C, 2) excluding the healthiest patients because they did not have any reported conditions in the six months before and after the COVID-19 diagnosis. The vaccination status had a similar limitation where vaccination data was reported by clinics, pharmacies, or patients (Blacketer, 2021). Due to this limitation, our sample may have underestimated the vaccination rates if patients received vaccinations at a facility not sharing data with N3C. The low vaccination rates in our data might have led to a weaker relationship between vaccination and long COVID symptoms in our findings. Due to our concerns with the vaccination variable, we choose not to include vaccination in our modeling.

## 4. Summary statistics for Cognitive, Fatigue, and Respiratory variations of Long COVID

We examined long COVID by three symptom clusters -- cognitive, fatigue, and respiratory -- as defined by the Global Burden of Disease Long COVID Collaborators (Global Burden of Disease Long COVID Collaborators, 2022). We classified patients as being positive for the individual symptom clusters if the patients had a qualifying symptom present between 3 to 6 months after having a lab confirmed case of COVID-19 and that symptom was not reported in the 6 months preceding their lab confirmed case of COVID-19. For a full list of the symptoms and OMOP codes we used as qualifying cognitive, fatigue, and respiratory long COVID symptoms, see Appendix A2, A3, and A4 respectively. Then we focused our analysis on the three separate symptom clusters and predicted the risk of a patient having had each symptom cluster of long COVID systems.

The fatigue long COVID symptom cluster was the most common in our sample with 4.7% (n = 57,483) of all patients, followed by respiratory cluster with 2.5% (n = 30,668) of our sample, and the cognitive symptom cluster with only 0.2% (n = 2,937) of the sample (Table 1). In general, for all three symptom clusters, there was an age gradient where patients in younger age groups have lower rates of symptom clusters than patients in older groups. Female patients had higher rates (5.7%) of the fatigue symptom cluster than male patients (3.3%) and there was no difference in rates between the sexes for the cognitive symptom clusters (female = 0.3%; male = 0.2%) and respiratory (female = 2.6%; male = 2.3%). Black and White patients had higher prevalence than other race groups for all three symptom clusters. There was not a meaningful difference in prevalence by ethnicity. Patients who had cognitive, fatigue, and respiratory symptom clusters had higher average CCI scores than those who did not experience said symptom clusters. The highest average CCI scores were patients who had cognitive cluster symptoms (mean = 2.7), compared to fatigue (mean = 1.4) and respiratory (mean = 1.4) symptom clusters. For other variables we did not observe large differences between groups.

## 5. Predictive model for long COVID outcomes

### Model training methods

We trained two types of machine learning models (a binary logistic regression model and a binary random forest model) on all three long COVID clusters separately: respiratory, fatigue, and cognitive. We used a 70-30 train-test split, resulting in 863,883 training observations and 370,236 test observations. To accommodate for long COVID being a rare outcome in all three of our clusters, we trained all models on down sampled data and tested them on the original, unbalanced data. To create the down sampled training data, we took five bootstrapped samples with replacement. Each bootstrapped sample had an equal number of positive and negative cases, which was also equal to the total number of positive cases in the original training data.

### Model performance

The random forest model outperformed the logistic regression model at predicting the respiratory and cognitive clusters, but the logistic regression model slightly outperformed the random forest model at predicting the fatigue outcome (Table 2). In all six models, the AUC remained fairly consistent between the train and test data, but the model precision (a measure of how often a model’s positive predictions are correct) dropped quite dramatically between the train and test data. This could be because long COVID was a rare outcome, or that there is low signal in the data.

**Table 2:**
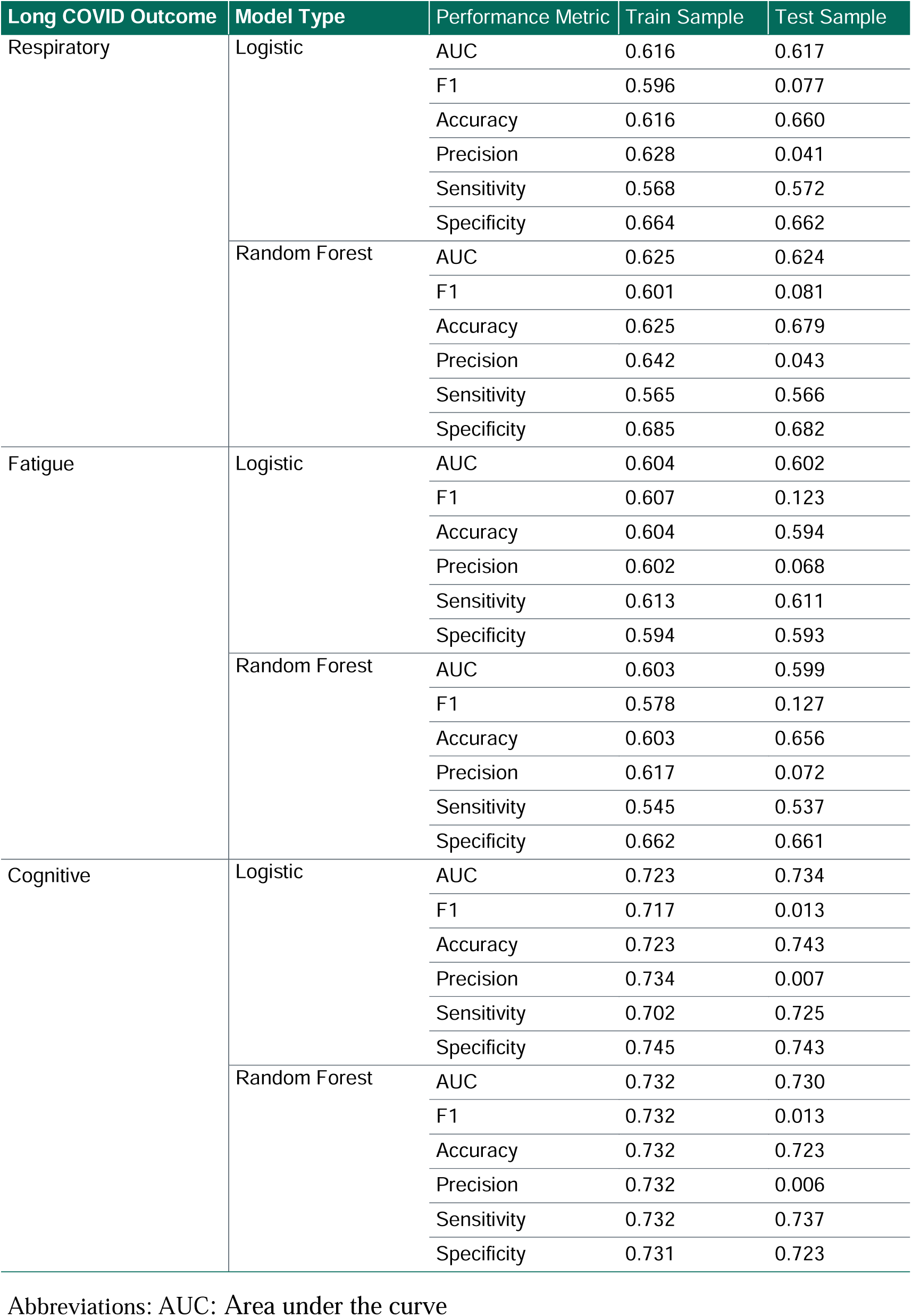
Model Performance Results.

| Long COVID Outcome | Model Type | Performance Metric | Train Sample | Test Sample |
| --- | --- | --- | --- | --- |
| Respiratory | Logistic | AUC | 0.616 | 0.617 |
|  |  | F1 | 0.596 | 0.077 |
|  |  | Accuracy | 0.616 | 0.660 |
|  |  | Precision | 0.628 | 0.041 |
|  |  | Sensitivity | 0.568 | 0.572 |
|  |  | Specificity | 0.664 | 0.662 |
|  | Random Forest | AUC | 0.625 | 0.624 |
|  |  | F1 | 0.601 | 0.081 |
|  |  | Accuracy | 0.625 | 0.679 |
|  |  | Precision | 0.642 | 0.043 |
|  |  | Sensitivity | 0.565 | 0.566 |
|  |  | Specificity | 0.685 | 0.682 |
| Fatigue | Logistic | AUC | 0.604 | 0.602 |
|  |  | F1 | 0.607 | 0.123 |
|  |  | Accuracy | 0.604 | 0.594 |
|  |  | Precision | 0.602 | 0.068 |
|  |  | Sensitivity | 0.613 | 0.611 |
|  |  | Specificity | 0.594 | 0.593 |
|  | Random Forest | AUC | 0.603 | 0.599 |
|  |  | F1 | 0.578 | 0.127 |
|  |  | Accuracy | 0.603 | 0.656 |
|  |  | Precision | 0.617 | 0.072 |
|  |  | Sensitivity | 0.545 | 0.537 |
|  |  | Specificity | 0.662 | 0.661 |
| Cognitive | Logistic | AUC | 0.723 | 0.734 |
|  |  | F1 | 0.717 | 0.013 |
|  |  | Accuracy | 0.723 | 0.743 |
|  |  | Precision | 0.734 | 0.007 |
|  |  | Sensitivity | 0.702 | 0.725 |
|  |  | Specificity | 0.745 | 0.743 |
|  | Random Forest | AUC | 0.732 | 0.730 |
|  |  | F1 | 0.732 | 0.013 |
|  |  | Accuracy | 0.732 | 0.723 |
|  |  | Precision | 0.732 | 0.006 |
|  |  | Sensitivity | 0.732 | 0.737 |
|  |  | Specificity | 0.731 | 0.723 |

Abbreviations: AUC: Area under the curve

### Covariates

To measure feature importance, we considered coefficient values for the binary logistic regression models and impurity-based variable importance scores for the random forest models (Tables 3 and 4). Among our six models, there were some variations in which covariates were most predictive. The CCI score was the most important feature in all three clusters for the random forest models. However, for the binary logistic regression models, all three clusters had different most important features (Table 3). For the respiratory cluster, the two most important features are belonging to the 0 – 9 age group and being a current or former smoker. For the fatigue cluster, the two most important features are belonging to the 0 – 9 age group and being male. Finally, for the cognitive cluster, the two most important features are being 80 years old or older and belonging to the 70 – 79 age group.

**Table 3:** Coefficient Values for the Binary Logistic Regression Models.

| Coefficient |  | Cognitive | Fatigue | Respiratory |
| --- | --- | --- | --- | --- |
| Age | 0-9 | -1.367 | -1.070 | 0.651 |
|  | 10-19 | -0.988 | -0.178 | -0.167 |
|  | 20-29 | -0.331 | 0.002 | -0.085 |
|  | 30-39 | Reference | Reference | Reference |
|  | 40-49 | 0.663 | -0.057 | 0.166 |
|  | 50-59 | 0.781 | -0.110 | 0.351 |
|  | 60-69 | 0.945 | -0.241 | 0.472 |
|  | 70-79 | 1.598 | -0.288 | 0.522 |
|  | 80+ | 1.892 | -0.287 | 0.504 |
| Sex | Female | Reference | Reference | Reference |
|  | Male | -0.211 | -0.539 | -0.176 |
|  | Other | -0.117 | -0.147 | -0.196 |
| Race | White | Reference | Reference | Reference |
|  | Black or African American | -0.054 | -0.178 | 0.084 |
|  | Asian | -0.064 | -0.274 | 0.019 |
|  | Native Hawaiian or Pacific Islander | -0.430 | -0.462 | -0.248 |
|  | Other | -0.454 | 0.111 | 0.052 |
|  | Missing/Unknown | -0.254 | -0.263 | -0.277 |
| Ethnicity | Not Hispanic or Latino | Reference | Reference | Reference |
|  | Hispanic or Latino | 0.084 | 0.063 | 0.016 |
|  | Ethnicity Missing or Unknown | -0.006 | -0.215 | -0.234 |
| COVID-19 vaccination | No | Reference | Reference | Reference |
|  | Yes | -0.397 | -0.406 | -0.338 |
| Smoking status: | Non-smoker | Reference | Reference | Reference |
|  | Current or Former smoker | 0.345 | 0.316 | 0.386 |
| Hypertension | No | Reference | Reference | Reference |
|  | Yes | 0.298 | 0.130 | -0.012 |
| Obesity | No | Reference | Reference | Reference |
|  | Yes | 0.431 | 0.429 | 0.413 |
| Immunocompromised | No | Reference | Reference | Reference |
|  | Yes | 0.214 | -0.056 | 0.092 |
| Pregnant | No | Reference | Reference | Reference |
|  | Yes | 0.293 | 0.286 | -0.076 |
| CCI Score |  | 0.237 | 0.160 | 0.166 |

Abbreviations: CCI: Charlson Comorbidity Index

**Table 4:** Impurity-Based Variable Importance Scores for the Random Forest Models.

| Variable Importance Score |  | Cognitive | Fatigue | Respiratory |
| --- | --- | --- | --- | --- |
| Age | 0-9 | 0.057 | 0.124 | 0.014 |
|  | 10-19 | 0.092 | 0.008 | 0.061 |
|  | 20-29 | 0.064 | 0.006 | 0.060 |
|  | 30-39 | Reference | Reference | Reference |
|  | 40-49 | 0.003 | 0.003 | 0.003 |
|  | 50-59 | 0.007 | 0.002 | 0.005 |
|  | 60-69 | 0.019 | 0.002 | 0.033 |
|  | 70-79 | 0.124 | 0.003 | 0.044 |
|  | 80+ | 0.078 | 0.002 | 0.012 |
| Sex | Female | Reference | Reference | Reference |
|  | Male | 0.007 | 0.214 | 0.008 |
|  | Other | 0.000 | 0.000 | 0.000 |
| Race | White | Reference | Reference | Reference |
|  | Black or African American | 0.004 | 0.003 | 0.007 |
|  | Asian | 0.003 | 0.003 | 0.001 |
|  | Native Hawaiian or Pacific Islander | 0.000 | 0.001 | 0.001 |
|  | Other | 0.002 | 0.001 | 0.001 |
|  | Missing/Unknown | 0.010 | 0.023 | 0.031 |
| Ethnicity | Not Hispanic or Latino | Reference | Reference | Reference |
|  | Hispanic or Latino | 0.006 | 0.004 | 0.004 |
|  | Ethnicity Missing or Unknown | 0.005 | 0.027 | 0.019 |
| COVID-19 vaccination | No | Reference | Reference | Reference |
|  | Yes | 0.003 | 0.003 | 0.003 |
| Smoking status: | Non-smoker | Reference | Reference | Reference |
|  | Current or Former smoker | 0.018 | 0.048 | 0.061 |
| Hypertension | No | Reference | Reference | Reference |
|  | Yes | 0.003 | 0.002 | 0.002 |
| Obesity | No | Reference | Reference | Reference |
|  | Yes | 0.045 | 0.128 | 0.122 |
| Immunocompromised | No | Reference | Reference | Reference |
|  | Yes | 0.000 | 0.000 | 0.000 |
| Pregnant | No | Reference | Reference | Reference |
|  | Yes | 0.001 | 0.001 | 0.001 |
| CCI Score |  | 0.445 | 0.394 | 0.507 |

601 Abbreviations: CCI: Charlson Comorbidity Index

### Comparisons with other long COVID predictive models

One challenge in comparing models is that there is not a consistent definition of long COVID that is universally accepted. The two papers described below offer two alternative definitions of long COVID.

One study also uses data from N3C to predict outcomes of long COVID. The authors consider visiting a long COVID clinic as an indicator that a patient has long COVID (Pfaff, 2022). This is a narrower definition of long COVID than our definition in this study. Their XGBoost model results in an AUC of 0.92 for all patients in their sample, 0.90 for hospitalized patients, and 0.85 for non-hospitalized patients. The simplicity of their measure could explain why their results are better than the results from our models. However, there are also drawbacks to defining long COVID as a visit to a long COVID clinic. a long COVID clinic is a very specific type of care and someone’s long COVID symptoms have to be pretty severe to be referred to a long COVID clinic. Thus, the Pfaff definition undercounts the number of patients with long COVID especially patients with more mild symptoms of long COVID.

Another study used age, sex, and the number of symptoms a patient experienced in the first week of infection to classify duration of COVID-19 as either short (less than 10 days) or long (28 days or more, which is shorter than the WHO recommendation) using k-means clustering (Sudre et al., 2021). The authors analyzed self-reported data from 4,182 cases of COVID-19 through the COVID Symptom Study app and used a random forest model and a logistic regression model to predict long COVID, as defined by having symptoms for 28 days or longer. The random forest model, which included first week symptoms and other comorbidities, performed moderately well with an AUC of 0.768. The logistic regression model was much simpler and included only age, sex, and number of symptoms in the first week, with an AUC of 0.767. One key difference between Sudre et al and our model is that Sudre et al included the number of symptoms in the first week which is likely a proxy for identify which patients had more severe initial COVID infections, whereas our model had limited variables representing the severity of initial infection.

## 6. Apply the fair machine learning framework to the long COVID predictive models

Timely risk assessment of long COVID outcomes can improve patient care and healthcare resource allocation. However, disparities across different sex, race/ethnicity, and social economic status in COVID-19 patient outcomes have been well-documented(Kharroubi & Diab-El-Harake, 2022; Webb Hooper et al., 2020). For example, while males have high risks for severe COVID outcomes such as death and ICU admission, females have been reported in the literature to have higher risks for long COVID (Cohen & van der Meulen Rodgers, 2023). In addition, compared to white patients, patients from racial/ethnic minority groups had significantly different odds of developing long COVID symptoms and conditions (Khullar et al., 2023). It is important to carefully examine model fairness across different population subgroups to achieve optimal and fair clinical decision-making.

We plan to confirm our models achieve similar performances across different sex/race/ethnicity groups by calculating the performance score using AUROC, F1 score, precision, recall, sensitivity, and specificity for males/females, different race groups including White, Black or African American, Asian, Native Hawaiian or Pacific Islander, and Other or Missing/Unknown, and different ethnicity (Hispanic and non-Hispanic) groups. This will allow us to detect if there is any bias in model performance. In addition to model performance, other fairness metrics will also be evaluated since the outcome (diagnoses codes in EHR) is not considered “ground truth”. We will also evaluate predictive equality, equal opportunity, and statistical parity of the model predictions.

If we detect disparity in fairness metrics, we plan to address it using various techniques including disparate impact remover (a preprocessing technique that edits input values to increase fairness between groups), reweighting (producing weights for each subgroup for each outcome class to achieve statistical parity), and resampling (oversampling or under-sampling to achieve statistical parity). We will compare the effects of different mitigation techniques and discuss their impact on the balance between model performance and fairness metrics. The method maximizing model predictive performance may be different from the method achieving the highest fairness metrics. We will discuss the trade-off between model performance and fairness in the context of using real-world EHR data to develop predictive models for long COVID.

## Conclusion

### Overarching takeaways regarding model performance

In this study, we explored the feasibility to train a predictive model to provide insights into the risk factors for long COVID. We leveraged a diverse and harmonized electronic health record data source and applied minimum filters to build a model with high potential to be applied to general population. After using techniques to handle unbalanced sample, our predictive models achieved moderate performance accuracy (AUC 0.599 – 0.730 for the random forest models, AUC 0.602 – 0.734 for the logistic regression models). It was interesting to observe that logistic regression models have comparable performance to random forest models, potentially because we included relatively limited number of predictors that are easy to acquire. Other predictive models developed in the past had stronger performances but they either leveraged clinical factors that can be hard to acquire such as blood oxygen, blood pH (Bennett et al., 2021), or they limited the population to a small and relatively homogenous groups of people, such as users of the COVID Symptom Study app or patients visiting a long COVID clinic (Pfaff, 2022; Sudre et al., 2021).

### Comparison with 19&Me severe COVID risk model

Previously we leveraged the same data source (N3C) and machine learning models to build a predictive model for severe COVID-19 outcomes. Severe COVID-19 outcome is defined as having a score of 6 or above using the Clinical Progression Scale (CPS) established by the World Health Organization for COVID-19 clinical research (Marshall et al., 2020). In a sample of 864,8080 COVID-19 positive patients, 15,401 (1.75%) has this outcome. Many risk factors are related to elevated COVID-19 risk, and others have published machine learning models that achieve high predictive performance (AUROC = 0.87) using a long list of predictors including demographics (age/sex/race), health conditions (comorbidities) and clinical characteristics (blood pH, respiratory rate, oxygen saturation…). We limit the predictors to those easily accessible in the 19andMe app (age/sex/race/comorbidities/smoking status/vaccination status). Since the outcome is highly unbalanced, we used down sampling and up weighting to handle the sample unbalance. We experimented with different modeling techniques such as random forest and logistic regression, with and without the downsampling techniques. The best performing model was the logistic regression with downsampled data (Table 2).

### Implications for future work: learnings on N3C data

The N3C is a nationally representative, harmonized data resource that we can leverage for COVID-19 research, among other data sources that our team already have experience with. In addition to the data tables that we explored in the current work, several other data tables such as visitation occurrence can be potentially useful for future similar work. In our current work, we can only create a flag variable for whether or not the patient has certain symptoms, without the ability to rate its severity. Information in the visitation occurrence table can be helpful because it has visit start and end date, which can then be used to characterize the severity of the condition. This can be a future area of modeling enhancement. Machine learning models have utilities in building predictive models for both severe COVID outcomes and long COVID. The severe COVID model had better performance, partially because of a clear definition of severe COVID and relative follow-up period. The model building process could benefit from close collaborations between data scientists, health service researchers and clinicians. The models developed in this study may be valuable to both payers to improve their understanding of the risks of their insured population, and to public health officials to better plan for the resources needed to improve population health in the aftermaths of COVID-19 pandemic.

## Data Availability

All data produced are available online at National COVID Cohort Collaborative Data Enclave

https://covid.cd2h.org

## Acknowledgements

This work is supported by Mathematica.

## N3C Attribution

The analyses described in this manuscript were conducted with data or tools accessed through the NCATS N3C Data Enclave https://covid.cd2h.org and N3C Attribution & Publication Policy v 1.2-2020-08-25b supported by NCATS Contract No. 75N95023D00001, Axle Informatics Subcontract: NCATS-P00438-B, and [insert additional funding agencies or sources and reference numbers as declared by the contributors in their form response above]. This research was possible because of the patients whose information is included within the data and the organizations (https://ncats.nih.gov/n3c/resources/data-contribution/data-transfer-agreement-signatories) and scientists who have contributed to the on-going development of this community resource [https://doi.org/10.1093/jamia/ocaa196].

Disclaimer The N3C Publication committee confirmed that this manuscript msid:1865.948 is in accordance with N3C data use and attribution policies; however, this content is solely the responsibility of the authors and does not necessarily represent the official views of the National Institutes of Health or the N3C program.

## IRB

The N3C data transfer to NCATS is performed under a Johns Hopkins University Reliance Protocol # IRB00249128 or individual site agreements with NIH. The N3C Data Enclave is managed under the authority of the NIH; information can be found at https://ncats.nih.gov/n3c/resources.

## Individual Acknowledgements For Core Contributors

We gratefully acknowledge the following core contributors to N3C:

Adam B. Wilcox, Adam M. Lee, Alexis Graves, Alfred (Jerrod) Anzalone, Amin Manna, Amit Saha, Amy Olex, Andrea Zhou, Andrew E. Williams, Andrew Southerland, Andrew T. Girvin, Anita Walden, Anjali A. Sharathkumar, Benjamin Amor, Benjamin Bates, Brian Hendricks, Brijesh Patel, Caleb Alexander, Carolyn Bramante, Cavin Ward-Caviness, Charisse Madlock-Brown, Christine Suver, Christopher Chute, Christopher Dillon, Chunlei Wu, Clare Schmitt, Cliff Takemoto, Dan Housman, Davera Gabriel, David A. Eichmann, Diego Mazzotti, Don Brown, Eilis Boudreau, Elaine Hill, Elizabeth Zampino, Emily Carlson Marti, Emily R. Pfaff, Evan French, Farrukh M Koraishy, Federico Mariona, Fred Prior, George Sokos, Greg Martin, Harold Lehmann, Heidi Spratt, Hemalkumar Mehta, Hongfang Liu, Hythem Sidky, J.W. Awori Hayanga, Jami Pincavitch, Jaylyn Clark, Jeremy Richard Harper, Jessica Islam, Jin Ge, Joel Gagnier, Joel H. Saltz, Joel Saltz, Johanna Loomba, John Buse, Jomol Mathew, Joni L. Rutter, Julie A. McMurry, Justin Guinney, Justin Starren, Karen Crowley, Katie Rebecca Bradwell, Kellie M. Walters, Ken Wilkins, Kenneth R. Gersing, Kenrick Dwain Cato, Kimberly Murray, Kristin Kostka, Lavance Northington, Lee Allan Pyles, Leonie Misquitta, Lesley Cottrell, Lili Portilla, Mariam Deacy, Mark M. Bissell, Marshall Clark, Mary Emmett, Mary Morrison Saltz, Matvey B. Palchuk, Melissa A. Haendel, Meredith Adams, Meredith Temple-O’Connor, Michael G. Kurilla, Michele Morris, Nabeel Qureshi, Nasia Safdar, Nicole Garbarini, Noha Sharafeldin, Ofer Sadan, Patricia A. Francis, Penny Wung Burgoon, Peter Robinson, Philip R.O. Payne, Rafael Fuentes, Randeep Jawa, Rebecca Erwin-Cohen, Rena Patel, Richard A. Moffitt, Richard L. Zhu, Rishi Kamaleswaran, Robert Hurley, Robert T. Miller, Saiju Pyarajan, Sam G. Michael, Samuel Bozzette, Sandeep Mallipattu, Satyanarayana Vedula, Scott Chapman, Shawn T. O’Neil, Soko Setoguchi, Stephanie S. Hong, Steve Johnson, Tellen D. Bennett, Tiffany Callahan, Umit Topaloglu, Usman Sheikh, Valery Gordon, Vignesh Subbian, Warren A. Kibbe, Wenndy Hernandez, Will Beasley, Will Cooper, William Hillegass, Xiaohan Tanner Zhang. Details of contributions available at covid.cd2h.org/core-contributors

## Data Partners with Released Data

The following institutions whose data is released or pending:

Available: Advocate Health Care Network — UL1TR002389: The Institute for Translational Medicine (ITM) • Aurora Health Care Inc — UL1TR002373: Wisconsin Network For Health Research • Boston University Medical Campus — UL1TR001430: Boston University Clinical and Translational Science Institute • Brown University — U54GM115677: Advance Clinical Translational Research (Advance-CTR) • Carilion Clinic — UL1TR003015: iTHRIV Integrated Translational health Research Institute of Virginia • Case Western Reserve University — UL1TR002548: The Clinical & Translational Science Collaborative of Cleveland (CTSC) • Charleston Area Medical Center — U54GM104942: West Virginia Clinical and Translational Science Institute (WVCTSI) • Children’s Hospital Colorado — UL1TR002535: Colorado Clinical and Translational Sciences Institute • Columbia University Irving Medical Center — UL1TR001873: Irving Institute for Clinical and Translational Research • Dartmouth College — None (Voluntary) Duke University — UL1TR002553: Duke Clinical and Translational Science Institute • George Washington Children’s Research Institute — UL1TR001876: Clinical and Translational Science Institute at Children’s National (CTSA-CN) • George Washington University — UL1TR001876: Clinical and Translational Science Institute at Children’s National (CTSA-CN) • Harvard Medical School — UL1TR002541: Harvard Catalyst • Indiana University School of Medicine — UL1TR002529: Indiana Clinical and Translational Science Institute • Johns Hopkins University — UL1TR003098: Johns Hopkins Institute for Clinical and Translational Research • Louisiana Public Health Institute — None (Voluntary) • Loyola Medicine — Loyola University Medical Center • Loyola University Medical Center — UL1TR002389: The Institute for Translational Medicine (ITM) • Maine Medical Center — U54GM115516: Northern New England Clinical & Translational Research (NNE-CTR) Network • Mary Hitchcock Memorial Hospital & Dartmouth Hitchcock Clinic — None (Voluntary) • Massachusetts General Brigham — UL1TR002541: Harvard Catalyst • Mayo Clinic Rochester — UL1TR002377: Mayo Clinic Center for Clinical and Translational Science (CCaTS) • Medical University of South Carolina — UL1TR001450: South Carolina Clinical & Translational Research Institute (SCTR) • MITRE Corporation — None (Voluntary) • Montefiore Medical Center — UL1TR002556: Institute for Clinical and Translational Research at Einstein and Montefiore • Nemours — U54GM104941: Delaware CTR ACCEL Program • NorthShore University HealthSystem — UL1TR002389: The Institute for Translational Medicine (ITM) • Northwestern University at Chicago — UL1TR001422: Northwestern University Clinical and Translational Science Institute (NUCATS) • OCHIN — INV-018455: Bill and Melinda Gates Foundation grant to Sage Bionetworks • Oregon Health & Science University — UL1TR002369: Oregon Clinical and Translational Research Institute • Penn State Health Milton S. Hershey Medical Center — UL1TR002014: Penn State Clinical and Translational Science Institute • Rush University Medical Center — UL1TR002389: The Institute for Translational Medicine (ITM) • Rutgers, The State University of New Jersey — UL1TR003017: New Jersey Alliance for Clinical and Translational Science • Stony Brook University — U24TR002306 • The Alliance at the University of Puerto Rico, Medical Sciences Campus — U54GM133807: Hispanic Alliance for Clinical and Translational Research (The Alliance) • The Ohio State University — UL1TR002733: Center for Clinical and Translational Science • The State University of New York at Buffalo — UL1TR001412: Clinical and Translational Science Institute • The University of Chicago — UL1TR002389: The Institute for Translational Medicine (ITM) • The University of Iowa — UL1TR002537: Institute for Clinical and Translational Science • The University of Miami Leonard M. Miller School of Medicine — UL1TR002736: University of Miami Clinical and Translational Science Institute • The University of Michigan at Ann Arbor — UL1TR002240: Michigan Institute for Clinical and Health Research • The University of Texas Health Science Center at Houston — UL1TR003167: Center for Clinical and Translational Sciences (CCTS) • The University of Texas Medical Branch at Galveston — UL1TR001439: The Institute for Translational Sciences • The University of Utah — UL1TR002538: Uhealth Center for Clinical and Translational Science • Tufts Medical Center — UL1TR002544: Tufts Clinical and Translational Science Institute • Tulane University — UL1TR003096: Center for Clinical and Translational Science • The Queens Medical Center — None (Voluntary) • University Medical Center New Orleans — U54GM104940: Louisiana Clinical and Translational Science (LA CaTS) Center • University of Alabama at Birmingham — UL1TR003096: Center for Clinical and Translational Science • University of Arkansas for Medical Sciences — UL1TR003107: UAMS Translational Research Institute • University of Cincinnati — UL1TR001425: Center for Clinical and Translational Science and Training • University of Colorado Denver, Anschutz Medical Campus — UL1TR002535: Colorado Clinical and Translational Sciences Institute • University of Illinois at Chicago — UL1TR002003: UIC Center for Clinical and Translational Science • University of Kansas Medical Center — UL1TR002366: Frontiers: University of Kansas Clinical and Translational Science Institute • University of Kentucky — UL1TR001998: UK Center for Clinical and Translational Science • University of Massachusetts Medical School Worcester — UL1TR001453: The UMass Center for Clinical and Translational Science (UMCCTS) • University Medical Center of Southern Nevada — None (voluntary) • University of Minnesota — UL1TR002494: Clinical and Translational Science Institute • University of Mississippi Medical Center — U54GM115428: Mississippi Center for Clinical and Translational Research (CCTR) • University of Nebraska Medical Center — U54GM115458: Great Plains IDeA-Clinical & Translational Research • University of North Carolina at Chapel Hill — UL1TR002489: North Carolina Translational and Clinical Science Institute • University of Oklahoma Health Sciences Center — U54GM104938: Oklahoma Clinical and Translational Science Institute (OCTSI) • University of Pittsburgh — UL1TR001857: The Clinical and Translational Science Institute (CTSI) • University of Pennsylvania — UL1TR001878: Institute for Translational Medicine and Therapeutics • University of Rochester — UL1TR002001: UR Clinical & Translational Science Institute • University of Southern California — UL1TR001855: The Southern California Clinical and Translational Science Institute (SC CTSI) • University of Vermont — U54GM115516: Northern New England Clinical & Translational Research (NNE-CTR) Network • University of Virginia — UL1TR003015: iTHRIV Integrated Translational health Research Institute of Virginia • University of Washington — UL1TR002319: Institute of Translational Health Sciences • University of Wisconsin-Madison — UL1TR002373: UW Institute for Clinical and Translational Research • Vanderbilt University Medical Center — UL1TR002243: Vanderbilt Institute for Clinical and Translational Research • Virginia Commonwealth University — UL1TR002649: C. Kenneth and Dianne Wright Center for Clinical and Translational Research • Wake Forest University Health Sciences — UL1TR001420: Wake Forest Clinical and Translational Science Institute • Washington University in St. Louis — UL1TR002345: Institute of Clinical and Translational Sciences • Weill Medical College of Cornell University — UL1TR002384: Weill Cornell Medicine Clinical and Translational Science Center • West Virginia University — U54GM104942: West Virginia Clinical and Translational Science Institute (WVCTSI)lJ Submitted: Icahn School of Medicine at Mount Sinai — UL1TR001433: ConduITS Institute for Translational Sciences • The University of Texas Health Science Center at Tyler — UL1TR003167: Center for Clinical and Translational Sciences (CCTS) • University of California, Davis — UL1TR001860: UCDavis Health Clinical and Translational Science Center • University of California, Irvine — UL1TR001414: The UC Irvine Institute for Clinical and Translational Science (ICTS) • University of California, Los Angeles — UL1TR001881: UCLA Clinical Translational Science Institute • University of California, San Diego — UL1TR001442: Altman Clinical and Translational Research Institute • University of California, San Francisco — UL1TR001872: UCSF Clinical and Translational Science InstitutelJ NYU Langone Health Clinical Science Core, Data Resource Core, and PASC Biorepository Core — OTA-21-015A: Post-Acute Sequelae of SARS-CoV-2 Infection Initiative (RECOVER)lJ Pending: Arkansas Children’s Hospital — UL1TR003107: UAMS Translational Research Institute • Baylor College of Medicine — None (Voluntary) • Children’s Hospital of Philadelphia — UL1TR001878: Institute for Translational Medicine and Therapeutics • Cincinnati Children’s Hospital Medical Center — UL1TR001425: Center for Clinical and Translational Science and Training • Emory University — UL1TR002378: Georgia Clinical and Translational Science Alliance • HonorHealth — None (Voluntary) • Loyola University Chicago — UL1TR002389: The Institute for Translational Medicine (ITM) • Medical College of Wisconsin — UL1TR001436: Clinical and Translational Science Institute of Southeast Wisconsin • MedStar Health Research Institute — None (Voluntary) • Georgetown University — UL1TR001409: The Georgetown-Howard Universities Center for Clinical and Translational Science (GHUCCTS) • MetroHealth — None (Voluntary) • Montana State University — U54GM115371: American Indian/Alaska Native CTR • NYU Langone Medical Center — UL1TR001445: Langone Health’s Clinical and Translational Science Institute • Ochsner Medical Center — U54GM104940: Louisiana Clinical and Translational Science (LA CaTS) Center • Regenstrief Institute — UL1TR002529: Indiana Clinical and Translational Science Institute • Sanford Research — None (Voluntary) • Stanford University — UL1TR003142: Spectrum: The Stanford Center for Clinical and Translational Research and Education • The Rockefeller University — UL1TR001866: Center for Clinical and Translational Science • The Scripps Research Institute — UL1TR002550: Scripps Research Translational Institute • University of Florida — UL1TR001427: UF Clinical and Translational Science Institute • University of New Mexico Health Sciences Center — UL1TR001449: University of New Mexico Clinical and Translational Science Center • University of Texas Health Science Center at San Antonio — UL1TR002645: Institute for Integration of Medicine and Science • Yale New Haven Hospital — UL1TR001863: Yale Center for Clinical Investigation

## Footnotes

1 Tire 2 access is the patient level EHR data where 17 patient identifier variables are removed and longitudinal data is data-shifted to safeguard privacy (Haendel et al., 2021). We selected patients for the analysis from the Patient Severity and Scores dataset (Release-v70-2022-03-19). The dataset contains patient level information from all sites. N3C identifies positive COVID-19 cases through their COVID-19 Phenotype Inclusion Criteria (Pfaff, 2022). Though this did not factor into our analysis, another feature of the N3C data is a 1:2 case to control ratio of patients with identifies lab-confirmed, suspected, and possible cases of COVID-19 to patients who have been screen and tested negative for COVID-19. In addition, we linked patient ids from the Patient Severity and Scores dataset with two additional datasets provided in N3C, the condition occurrence dataset (Release-v70-2022-03-19) and the drug exposures dataset (Release-v70-2022-03-19) to provide additional longitudinal information about the patients in the database. These condition occurrence dataset records diagnosis, signs, and symptoms of conditions and drug exposure dataset records introduction of drugs into the body of the patient overtime (Observational Health Data Sciences and Informatics, 2018).

2 For selecting the patients how has conditions occurrences within 6 months preceding and proceeding diagnosis, we used the condition occurrence dataset (Release-v70-2022-03-19)

## References

Bennett, T. D., Moffitt, R. A., Hajagos, J. G., Amor, B., Anand, A., Bissell, M. M., Bradwell, K. R., Bremer, C., Byrd, J. B., Denham, A., DeWitt, P. E., Gabriel, D., Garibaldi, B. T., Girvin, A. T., Guinney, J., Hill, E. L., Hong, S. S., Jimenez, H., Kavuluru, R., … National COVID Cohort Collaborative (N3C) Consortium. (2021). Clinical Characterization and Prediction of Clinical Severity of SARS-CoV-2 Infection Among US Adults Using Data From the US National COVID Cohort Collaborative. JAMA Network Open, 4(7), e2116901. 10.1001/jamanetworkopen.2021.16901

Blacketer, C. (2021, January 11). Chapter 4 The Common Data Model. The Book of OHDSI. https://ohdsi.github.io/TheBookOfOhdsi/

CDC. (2022, June 22). Nearly One in Five American Adults Who Have Had COVID-19 Still Have “Long COVID.” https://www.cdc.gov/nchs/pressroom/nchs_press_releases/2022/20220622.htm

CDC. (2023, January 4). Long COVID - Household Pulse Survey—COVID-19. https://www.cdc.gov/nchs/covid19/pulse/long-covid.htm

Cohen, J., & van der Meulen Rodgers, Y. (2023). An intersectional analysis of long COVID prevalence. International Journal for Equity in Health, 22, 261. 10.1186/s12939-023-02072-5

Crook, H., Raza, S., Nowell, J., Young, M., & Edison, P. (2021). Long covid—Mechanisms, risk factors, and management. BMJ, 374, n1648. 10.1136/bmj.n1648

Cutler, D. (2022, July 22). The Economic Cost of Long COVID: An Update. https://scholar.harvard.edu/files/cutler/files/long_covid_update_7-22.pdf

Cutler, D. M., & Summers, L. H. (2020). The COVID-19 Pandemic and the $16 Trillion Virus. JAMA, 324(15), 1495–1496. 10.1001/jama.2020.19759

Glasheen, W. P., Cordier, T., Gumpina, R., Haugh, G., Davis, J., & Renda, A. (2019). Charlson Comorbidity Index: ICD-9 Update and ICD-10 Translation. American Health & Drug Benefits, 12(4), Article 4.

Global Burden of Disease Long COVID Collaborators. (2022). Estimated Global Proportions of Individuals With Persistent Fatigue, Cognitive, and Respiratory Symptom Clusters Following Symptomatic COVID-19 in 2020 and 2021. JAMA, 328(16), 1604–1615. 10.1001/jama.2022.18931

Haendel, M. A., Chute, C. G., Bennett, T. D., Eichmann, D. A., Guinney, J., Kibbe, W. A., Payne, P. R. O., Pfaff, E. R., Robinson, P. N., Saltz, J. H., Spratt, H., Suver, C., Wilbanks, J., Wilcox, A. B., Williams, A. E., Wu, C., Blacketer, C., Bradford, R. L., Cimino, J. J., … N3C Consortium. (2021). The National COVID Cohort Collaborative (N3C): Rationale, design, infrastructure, and deployment. Journal of the American Medical Informatics Association: JAMIA, 28(3), 427–443. 10.1093/jamia/ocaa196

Herman, E., Shih, E., & Cheng, A. (2022). Long COVID: Rapid Evidence Review. American Family Physician, 106(5), 523–532.

Kharroubi, S. A., & Diab-El-Harake, M. (2022). Sex-differences in COVID-19 diagnosis, risk factors and disease comorbidities: A large US-based cohort study. Frontiers in Public Health, 10, 1029190. 10.3389/fpubh.2022.1029190

Khullar, D., Zhang, Y., Zang, C., Xu, Z., Wang, F., Weiner, M. G., Carton, T. W., Rothman, R. L., Block, J. P., & Kaushal, R. (2023). Racial/Ethnic Disparities in Post-acute Sequelae of SARS-CoV-2 Infection in New York: An EHR-Based Cohort Study from the RECOVER Program. Journal of General Internal Medicine, 38(5), 1127–1136. 10.1007/s11606-022-07997-1

Klaassen, F., Chitwood, M. H., Cohen, T., Pitzer, V. E., Russi, M., Swartwood, N. A., Salomon, J. A., & Menzies, N. A. (2022). *Changes in population immunity against infection and severe disease from SARS-CoV-2 Omicron variants in the United States between December 2021 and November* 2022 (p. 2022.11.19.22282525). medRxiv. 10.1101/2022.11.19.22282525

Mancini, D. M., Brunjes, D. L., Lala, A., Trivieri, M. G., Contreras, J. P., & Natelson, B. H. (2021). Use of Cardiopulmonary Stress Testing for Patients With Unexplained Dyspnea Post–Coronavirus Disease. JACC: Heart Failure, 9(12), 927–937. 10.1016/j.jchf.2021.10.002

Marshall, J. C., Murthy, S., Diaz, J., Adhikari, N. K., Angus, D. C., Arabi, Y. M., Baillie, K., Bauer, M., Berry, S., Blackwood, B., Bonten, M., Bozza, F., Brunkhorst, F., Cheng, A., Clarke, M., Dat, V. Q., de Jong, M., Denholm, J., Derde, L., … Zhang, J. (2020). A minimal common outcome measure set for COVID-19 clinical research. The Lancet Infectious Diseases, 20(8), e192–e197. 10.1016/S1473-3099(20)30483-7

Nasserie, T., Hittle, M., & Goodman, S. N. (2021). Assessment of the Frequency and Variety of Persistent Symptoms Among Patients With COVID-19: A Systematic Review. JAMA Network Open, 4(5), e2111417. 10.1001/jamanetworkopen.2021.11417

Nittas, V., Gao, M., West, E. A., Ballouz, T., Menges, D., Wulf Hanson, S., & Puhan, M. A. (2022). Long COVID Through a Public Health Lens: An Umbrella Review. Public Health Reviews, 43, 1604501. 10.3389/phrs.2022.1604501

Observational Health Data Sciences and Informatics. (2018). COVID-19 Clinical Data Warehouse Data Dictionary Based on OMOP Common Data Model Specifications Version 5.3. National Center for Advancing Translational Sciences (NCATS). https://ncats.nih.gov/files/OMOP_CDM_COVID.pdf

Patient Led Research Collaborative. (2023). About the Patient-Led Research Collaborative. Patient Led Research Collaborative – for Long COVID. https://patientresearchcovid19.com/

Pfaff, E. R. (2022, March 11). Latest Phenotype [GitHub]. National-COVID-Cohort-Collaborative. https://github.com/National-COVID-Cohort-Collaborative/Phenotype_Data_Acquisition/wiki/Latest-Phenotype

Sudre, C. H., Murray, B., Varsavsky, T., Graham, M. S., Penfold, R. S., Bowyer, R. C., Pujol, J. C., Klaser, K., Antonelli, M., Canas, L. S., Molteni, E., Modat, M., Jorge Cardoso, M., May, A., Ganesh, S., Davies, R., Nguyen, L. H., Drew, D. A., Astley, C. M., … Steves, C. J. (2021). Attributes and predictors of long COVID. Nature Medicine, 27(4), Article 4. 10.1038/s41591-021-01292-y

Tsampasian, V., Elghazaly, H., Chattopadhyay, R., Debski, M., Naing, T. K. P., Garg, P., Clark, A., Ntatsaki, E., & Vassiliou, V. S. (2023). Risk Factors Associated With Post−COVID-19 Condition: A Systematic Review and Meta-analysis. JAMA Internal Medicine. 10.1001/jamainternmed.2023.0750

Webb Hooper, M., Nápoles, A. M., & Pérez-Stable, E. J. (2020). COVID-19 and Racial/Ethnic Disparities. JAMA, 323(24), 2466–2467. 10.1001/jama.2020.8598

World Health Organization. (2021, October 6). A clinical case definition of post COVID-19 condition by a Delphi consensus, 6 October 2021. https://www.who.int/publications-detail-redirect/WHO-2019-nCoV-Post_COVID-19_condition-Clinical_case_definition-2021.1

