## Appendices for "Can longitudinal electronic health record data identify patients at higher risk of developing long COVID?"

Appendix

Appendix A1: Covariate OMOP Codes

Table of OMOP Codes used for covariate creation

| Covariate | codeset_id | concept_id | concepts_set_name | is_most_recent_verison | verison | concept_name | archive |
| --- | --- | --- | --- | --- | --- | --- | --- |
| hypertension | 93041866 | 316866 | ARIScience - Hypertension - JA | FALSE | 1 | Hypertensive disorder | FALSE |
| hypertension | 343580642 | 316866 | Atlas 807 N3C Essential Hypertension | TRUE | 2 | Hypertensive disorder | FALSE |
| hypertension | 779214702 | 316866 | Hypertension_Elixhauser_UC | TRUE | 1 | Hypertensive disorder | FALSE |
| hypertension | 859645468 | 316866 | hypertensive-disorder-107 | TRUE | 1 | Hypertensive disorder | FALSE |
| hypertension | 972807308 | 316866 | Pre-existing hypertension pregnancy | TRUE | 2 | Hypertensive disorder | FALSE |
| hypertension | 146797511 | 316866 | [LEGEND]Hypertension | TRUE | 1 | Hypertensive disorder | FALSE |
| hypertension | 179876469 | 316866 | hypertension-bdc | TRUE | 2 | Hypertensive disorder | FALSE |
| hypertension | 534314902 | 316866 | [Cardioonc] All CV Disease | TRUE | 1 | Hypertensive disorder | FALSE |
| hypertension | 119899917 | 316866 | [RHDT] Hypertension | TRUE | 2 | Hypertensive disorder | FALSE |
| hypertension | 862684480 | 316866 | hypertension-bdc | FALSE | 1 | Hypertensive disorder | FALSE |
| hypertension | 142602231 | 316866 | Hypertensive disorder | TRUE | 1 | Hypertensive disorder | FALSE |
| hypertension | 378884384 | 316866 | Hypertension_wide | TRUE | 1 | Hypertensive disorder | FALSE |
| hypertension | 537394386 | 316866 | jr_hypertension | TRUE | 1 | Hypertensive disorder | FALSE |
| hypertension | 799270877 | 316866 | ARIScience - Hypertension - JA | TRUE | 2 | Hypertensive disorder | FALSE |
| obesity | 229321427 | 433736 | obesity | TRUE | 1 | Obesity | FALSE |
| obesity | 236332904 | 433736 | UVA Obesity | FALSE | 1 | Obesity | FALSE |
| obesity | 548507987 | 433736 | cs_isc_obesity | TRUE | 1 | Obesity | FALSE |
| obesity | 611206754 | 433736 | Obesity | TRUE | 1 | Obesity | TRUE |
| obesity | 856365627 | 433736 | (RHDT) Obesity | TRUE | 1 | Obesity | FALSE |
| obesity | 25920448 | 433736 | [VSAC] Overweight or Obese | TRUE | 2 | Obesity | FALSE |
| obesity | 748628806 | 433736 | [neuro3x] obesity | TRUE | 1 | Obesity | FALSE |
| obesity | 140513760 | 433736 | [VSAC] Obesity | TRUE | 1 | Obesity | FALSE |
| obesity | 372000305 | 433736 | [VSAC] Overweight or Obese | FALSE | 1 | Obesity | FALSE |
| obesity | 652960807 | 433736 | [VSAC] Obesity Conditions | TRUE | 1 | Obesity | TRUE |
| obesity | 5957480 | 433736 | LWW_Obesity | TRUE | 2 | Obesity | FALSE |
| obesity | 533705532 | 433736 | ARIScience - Obesity | TRUE | 3 | Obesity | FALSE |
| obesity | 630695504 | 433736 | OBESITY | TRUE | 1 | Obesity | FALSE |
| obesity | 651016149 | 433736 | UVA Obesity | TRUE | 2 | Obesity | FALSE |
| obesity | 799016302 | 433736 | ARIScience - Obesity | FALSE | 1 | Obesity | FALSE |
| obesity | 835773409 | 433736 | ARIScience - Obesity | FALSE | 2 | Obesity | FALSE |
| immunocompromised | 829301938 | 4153516 | [DATOS] Immunocompromised Stated | TRUE | 2 | Patient immunocompromised | FALSE |
| immunocompromised | 370855955 | 4153516 | [DATOS] Immunocompromised Stated | FALSE | 1 | Patient immunocompromised | FALSE |
| pregnancy | 101607593 | 44837786 | Pregnancy status | TRUE | 1 | Pregnant state, incidental | FALSE |
| COVID-19 vaccination | 145590853 | 702866 | johnson and johnson vaccine | TRUE | 3 | SARS-COV-2 (COVID-19) vaccine, vector non-replicating, recombinant spike protein-Ad26, preservative free, 0.5 mL | FALSE |
| COVID-19 vaccination | 948645531 | 37003436 | vaccine_pfizer | TRUE | 1 | SARS-CoV-2 (COVID-19) vaccine, mRNA-BNT162b2 0.1 MG/ML Injectable Suspension | FALSE |
| COVID-19 vaccination | 935484078 | 37003436 | covid vaccine pfizer | TRUE | 1 | SARS-CoV-2 (COVID-19) vaccine, mRNA-BNT162b2 0.1 MG/ML Injectable Suspension | FALSE |
| COVID-19 vaccination | 837019313 | 37003432 | covid-19 vaccine - unknown type | TRUE | 1 | SARS-CoV-2 (COVID-19) vaccine, mRNA spike protein | FALSE |
| COVID-19 vaccination | 837019313 | 724904 | covid-19 vaccine - unknown type | TRUE | 1 | SARS-COV-2 (COVID-19) vaccine, UNSPECIFIED | FALSE |
| COVID-19 vaccination | 145590853 | 739906 | johnson and johnson vaccine | TRUE | 3 | SARS-COV-2 (COVID-19) vaccine, vector - Ad26 100000000000 UNT/ML Injectable Suspension | FALSE |
| COVID-19 vaccination | 247296404 | 739906 | johnson and johnson vaccine | FALSE | 1 | SARS-COV-2 (COVID-19) vaccine, vector - Ad26 100000000000 UNT/ML Injectable Suspension | FALSE |
| COVID-19 vaccination | 696924427 | 739906 | johnson and johnson vaccine | FALSE | 2 | SARS-COV-2 (COVID-19) vaccine, vector - Ad26 100000000000 UNT/ML Injectable Suspension | FALSE |
| COVID-19 vaccination | 696924427 | 724905 | johnson and johnson vaccine | FALSE | 2 | SARS-COV-2 (COVID-19) vaccine, vector non-replicating, recombinant spike protein-ChAdOx1, preservative free, 0.5 mL | FALSE |
| COVID-19 vaccination | 145590853 | 724905 | johnson and johnson vaccine | TRUE | 3 | SARS-COV-2 (COVID-19) vaccine, vector non-replicating, recombinant spike protein-ChAdOx1, preservative free, 0.5 mL | FALSE |
| COVID-19 vaccination | 705249656 | 37003518 | vaccine_moderna | TRUE | 1 | SARS-CoV-2 (COVID-19) vaccine, mRNA-1273 0.2 MG/ML Injectable Suspension | FALSE |
| COVID-19 vaccination | 100751863 | 37003518 | covid vaccine moderna | TRUE | 1 | SARS-CoV-2 (COVID-19) vaccine, mRNA-1273 0.2 MG/ML Injectable Suspension | FALSE |
| CCI score | 535274723 | 22839 | Charlson - Cancer | TRUE | 1 | Overlapping malignant neoplasm of larynx | FALSE |
| CCI score | 535274723 | 26361 | Charlson - Cancer | TRUE | 1 | Primary malignant neoplasm of pineal gland | FALSE |
| CCI score | 535274723 | 31509 | Charlson - Cancer | TRUE | 1 | Primary malignant neoplasm of tonsil | FALSE |
| CCI score | 535274723 | 78093 | Charlson - Cancer | TRUE | 1 | Primary malignant neoplasm of pleura | FALSE |
| CCI score | 535274723 | 79758 | Charlson - Cancer | TRUE | 1 | Primary malignant neoplasm of scrotum | FALSE |
| CCI score | 535274723 | 132572 | Charlson - Cancer | TRUE | 1 | Chronic myeloid leukemia in remission | FALSE |
| CCI score | 535274723 | 135764 | Charlson - Cancer | TRUE | 1 | Mycosis fungoides of lymph nodes of multiple sites | FALSE |
| CCI score | 535274723 | 136928 | Charlson - Cancer | TRUE | 1 | Hodgkin's disease, nodular sclerosis of intrathoracic lymph nodes | FALSE |
| CCI score | 535274723 | 137809 | Charlson - Cancer | TRUE | 1 | Primary malignant neoplasm of female breast | FALSE |
| CCI score | 535274723 | 192836 | Charlson - Cancer | TRUE | 1 | Primary malignant neoplasm of small intestine | FALSE |
| CCI score | 535274723 | 193138 | Charlson - Cancer | TRUE | 1 | Primary malignant neoplasm of lower third of esophagus | FALSE |
| CCI score | 535274723 | 193157 | Charlson - Cancer | TRUE | 1 | Hodgkin's disease, nodular sclerosis of spleen | FALSE |
| CCI score | 535274723 | 195197 | Charlson - Cancer | TRUE | 1 | Primary malignant neoplasm of vulva | FALSE |
| CCI score | 535274723 | 196044 | Charlson - Cancer | TRUE | 1 | Primary malignant neoplasm of stomach | FALSE |
| CCI score | 535274723 | 196047 | Charlson - Cancer | TRUE | 1 | Primary malignant neoplasm of parametrium | FALSE |
| CCI score | 535274723 | 197500 | Charlson - Cancer | TRUE | 1 | Primary malignant neoplasm of colon | FALSE |
| CCI score | 535274723 | 198085 | Charlson - Cancer | TRUE | 1 | Kaposi's sarcoma of gastrointestinal tract | FALSE |
| CCI score | 535274723 | 199747 | Charlson - Cancer | TRUE | 1 | Overlapping malignant neoplasm of small intestine | FALSE |
| CCI score | 535274723 | 199760 | Charlson - Cancer | TRUE | 1 | Hodgkin's disease, nodular sclerosis of intrapelvic lymph nodes | FALSE |
| CCI score | 535274723 | 316356 | Charlson - Cancer | TRUE | 1 | Hodgkin's disease of lymph nodes of multiple sites | FALSE |
| CCI score | 535274723 | 316644 | Charlson - Cancer | TRUE | 1 | Primary malignant neoplasm of heart | FALSE |
| CCI score | 535274723 | 321526 | Charlson - Cancer | TRUE | 1 | Monocytic leukemia | FALSE |
| CCI score | 535274723 | 378696 | Charlson - Cancer | TRUE | 1 | Primary malignant neoplasm of choroid | FALSE |
| CCI score | 535274723 | 432264 | Charlson - Cancer | TRUE | 1 | Primary malignant neoplasm of broad ligament | FALSE |
| CCI score | 535274723 | 433423 | Charlson - Cancer | TRUE | 1 | Primary malignant neoplasm of pancreatic duct | FALSE |
| CCI score | 535274723 | 433716 | Charlson - Cancer | TRUE | 1 | Primary malignant neoplasm of testis | FALSE |
| CCI score | 535274723 | 433975 | Charlson - Cancer | TRUE | 1 | Primary malignant neoplasm of cranial nerve | FALSE |
| CCI score | 535274723 | 434293 | Charlson - Cancer | TRUE | 1 | Primary malignant neoplasm of body of pancreas | FALSE |
| CCI score | 535274723 | 434587 | Charlson - Cancer | TRUE | 1 | Primary malignant neoplasm of dorsal surface of tongue | FALSE |
| CCI score | 535274723 | 435478 | Charlson - Cancer | TRUE | 1 | Primary malignant neoplasm of upper gum | FALSE |
| CCI score | 535274723 | 435754 | Charlson - Cancer | TRUE | 1 | Malignant tumor of ascending colon | FALSE |
| CCI score | 535274723 | 436926 | Charlson - Cancer | TRUE | 1 | Primary malignant neoplasm of cerebral meninges | FALSE |
| CCI score | 535274723 | 437224 | Charlson - Cancer | TRUE | 1 | Primary malignant neoplasm of greater curvature of stomach | FALSE |
| CCI score | 535274723 | 437798 | Charlson - Cancer | TRUE | 1 | Primary malignant neoplasm of splenic flexure of colon | FALSE |
| CCI score | 535274723 | 438367 | Charlson - Cancer | TRUE | 1 | Primary malignant neoplasm of nasal cavity | FALSE |
| CCI score | 535274723 | 438368 | Charlson - Cancer | TRUE | 1 | Primary malignant neoplasm of thymus | FALSE |
| CCI score | 535274723 | 438703 | Charlson - Cancer | TRUE | 1 | Hodgkin's disease, lymphocytic-histiocytic predominance of lymph nodes of axilla AND/OR upper limb | FALSE |
| CCI score | 535274723 | 438979 | Charlson - Cancer | TRUE | 1 | Primary malignant neoplasm of hepatic flexure of colon | FALSE |
| CCI score | 535274723 | 439270 | Charlson - Cancer | TRUE | 1 | Sézary's disease of intrapelvic lymph nodes | FALSE |
| CCI score | 535274723 | 439282 | Charlson - Cancer | TRUE | 1 | Hodgkin's disease, lymphocytic depletion of intrapelvic lymph nodes | FALSE |
| CCI score | 535274723 | 439392 | Charlson - Cancer | TRUE | 1 | Primary malignant neoplasm | FALSE |
| CCI score | 535274723 | 440344 | Charlson - Cancer | TRUE | 1 | Primary malignant neoplasm of lateral portion of floor of mouth | FALSE |
| CCI score | 535274723 | 441521 | Charlson - Cancer | TRUE | 1 | Malignant lymphoma of lymph nodes of axilla AND/OR upper limb | FALSE |
| CCI score | 535274723 | 442160 | Charlson - Cancer | TRUE | 1 | Hodgkin's disease, mixed cellularity of lymph nodes of inguinal region AND/OR lower limb | FALSE |
| CCI score | 535274723 | 442163 | Charlson - Cancer | TRUE | 1 | Hodgkin's disease, lymphocytic-histiocytic predominance of lymph nodes of head, face AND/OR neck | FALSE |
| CCI score | 535274723 | 443382 | Charlson - Cancer | TRUE | 1 | Malignant tumor of descending colon | FALSE |
| CCI score | 535274723 | 444224 | Charlson - Cancer | TRUE | 1 | Overlapping malignant neoplasm of lip, oral cavity and pharynx | FALSE |
| CCI score | 535274723 | 4001171 | Charlson - Cancer | TRUE | 1 | Liver cell carcinoma | FALSE |
| CCI score | 535274723 | 4002343 | Charlson - Cancer | TRUE | 1 | Overlapping malignant neoplasm of peripheral nerves and autonomic nervous system | FALSE |
| CCI score | 535274723 | 4002496 | Charlson - Cancer | TRUE | 1 | Mast cell leukemia (clinical) | FALSE |
| CCI score | 535274723 | 4003675 | Charlson - Cancer | TRUE | 1 | Primary malignant neoplasm of cloacogenic zone | FALSE |
| CCI score | 535274723 | 4033836 | Charlson - Cancer | TRUE | 1 | Melanoma in situ of eyelid, including canthus | FALSE |
| CCI score | 535274723 | 4038839 | Charlson - Cancer | TRUE | 1 | Hodgkin lymphoma, nodular lymphocyte predominance (clinical) | FALSE |
| CCI score | 535274723 | 4038845 | Charlson - Cancer | TRUE | 1 | Hairy cell leukemia (clinical) | FALSE |
| CCI score | 535274723 | 4079686 | Charlson - Cancer | TRUE | 1 | Acute megakaryoblastic leukemia | FALSE |
| CCI score | 535274723 | 4082487 | Charlson - Cancer | TRUE | 1 | Cutaneous/peripheral T-cell lymphoma | FALSE |
| CCI score | 535274723 | 4089665 | Charlson - Cancer | TRUE | 1 | Malignant tumor of peritoneum and retroperitoneum | FALSE |
| CCI score | 535274723 | 4089860 | Charlson - Cancer | TRUE | 1 | Malignant melanoma of lip | FALSE |
| CCI score | 535274723 | 4091464 | Charlson - Cancer | TRUE | 1 | Malignant neoplasm of nipple and areola of female breast | FALSE |
| CCI score | 535274723 | 4091490 | Charlson - Cancer | TRUE | 1 | Malignant neoplasm of cerebrum (excluding lobes and ventricles) | FALSE |
| CCI score | 535274723 | 4095312 | Charlson - Cancer | TRUE | 1 | Malignant tumor of nasopharynx | FALSE |
| CCI score | 535274723 | 4095432 | Charlson - Cancer | TRUE | 1 | Malignant neoplasm of liver and intrahepatic bile ducts | FALSE |
| CCI score | 535274723 | 4095748 | Charlson - Cancer | TRUE | 1 | Malignant neoplasm of fundus of corpus uteri | FALSE |
| CCI score | 535274723 | 4100425 | Charlson - Cancer | TRUE | 1 | Merkel cell carcinoma | FALSE |
| CCI score | 535274723 | 4114222 | Charlson - Cancer | TRUE | 1 | Malignant tumor of head and neck | FALSE |
| CCI score | 535274723 | 4130672 | Charlson - Cancer | TRUE | 1 | Neoplasm of multiple endocrine glands | FALSE |
| CCI score | 535274723 | 4147411 | Charlson - Cancer | TRUE | 1 | Follicular non-Hodgkin's lymphoma | FALSE |
| CCI score | 535274723 | 4156114 | Charlson - Cancer | TRUE | 1 | Primary malignant neoplasm of border of tongue | FALSE |
| CCI score | 535274723 | 4157456 | Charlson - Cancer | TRUE | 1 | Primary malignant neoplasm of skin head and neck | FALSE |
| CCI score | 535274723 | 4173974 | Charlson - Cancer | TRUE | 1 | B-cell prolymphocytic leukemia | FALSE |
| CCI score | 535274723 | 4180312 | Charlson - Cancer | TRUE | 1 | Neoplasm of extremity | FALSE |
| CCI score | 535274723 | 4212994 | Charlson - Cancer | TRUE | 1 | Extranodal NK/T-cell lymphoma, nasal type | FALSE |
| CCI score | 535274723 | 4244488 | Charlson - Cancer | TRUE | 1 | Malignant melanoma of skin of nose | FALSE |
| CCI score | 535274723 | 4246137 | Charlson - Cancer | TRUE | 1 | Primary malignant neoplasm of optic nerve | FALSE |
| CCI score | 535274723 | 4247719 | Charlson - Cancer | TRUE | 1 | Primary malignant neoplasm of ascending colon | FALSE |
| CCI score | 535274723 | 4299149 | Charlson - Cancer | TRUE | 1 | Anaplastic large T-cell systemic malignant lymphoma | FALSE |
| CCI score | 535274723 | 4312685 | Charlson - Cancer | TRUE | 1 | Primary malignant neoplasm of skin of lower limb | FALSE |
| CCI score | 535274723 | 37018934 | Charlson - Cancer | TRUE | 1 | Malignant carcinoid tumor of rectum | FALSE |
| CCI score | 535274723 | 40486465 | Charlson - Cancer | TRUE | 1 | Gastrointestinal stromal tumor of esophagus | FALSE |
| CCI score | 535274723 | 40492266 | Charlson - Cancer | TRUE | 1 | Gastrointestinal stromal tumor of stomach | FALSE |
| CCI score | 535274723 | 73153 | Charlson - Cancer | TRUE | 1 | Malignant neoplasm of lateral wall of urinary bladder | FALSE |
| CCI score | 535274723 | 76924 | Charlson - Cancer | TRUE | 1 | Primary malignant neoplasm of ureteric orifice of urinary bladder | FALSE |
| CCI score | 535274723 | 77812 | Charlson - Cancer | TRUE | 1 | Malignant neoplasm of anterior wall of urinary bladder | FALSE |
| CCI score | 535274723 | 80340 | Charlson - Cancer | TRUE | 1 | Primary malignant neoplasm of epididymis | FALSE |
| CCI score | 535274723 | 133147 | Charlson - Cancer | TRUE | 1 | Primary malignant neoplasm of skin of trunk | FALSE |
| CCI score | 535274723 | 133158 | Charlson - Cancer | TRUE | 1 | Plasma cell leukemia in remission | FALSE |
| CCI score | 535274723 | 133711 | Charlson - Cancer | TRUE | 1 | Overlapping malignant neoplasm of female breast | FALSE |
| CCI score | 535274723 | 133969 | Charlson - Cancer | TRUE | 1 | Primary malignant neoplasm of vermilion border of lip | FALSE |
| CCI score | 535274723 | 134603 | Charlson - Cancer | TRUE | 1 | Chronic myeloid leukemia | FALSE |
| CCI score | 535274723 | 139753 | Charlson - Cancer | TRUE | 1 | Primary malignant neoplasm of parathyroid gland | FALSE |
| CCI score | 535274723 | 140958 | Charlson - Cancer | TRUE | 1 | Kaposi's sarcoma of palate | FALSE |
| CCI score | 535274723 | 141524 | Charlson - Cancer | TRUE | 1 | Mycosis fungoides of lymph nodes of axilla AND/OR upper limb | FALSE |
| CCI score | 535274723 | 192847 | Charlson - Cancer | TRUE | 1 | Primary malignant neoplasm of isthmus of uterus | FALSE |
| CCI score | 535274723 | 193418 | Charlson - Cancer | TRUE | 1 | Primary malignant neoplasm of spleen | FALSE |
| CCI score | 535274723 | 195760 | Charlson - Cancer | TRUE | 1 | Sézary's disease of spleen | FALSE |
| CCI score | 535274723 | 196049 | Charlson - Cancer | TRUE | 1 | Primary malignant neoplasm of urethra | FALSE |
| CCI score | 535274723 | 196359 | Charlson - Cancer | TRUE | 1 | Primary malignant neoplasm of uterine cervix | FALSE |
| CCI score | 535274723 | 197803 | Charlson - Cancer | TRUE | 1 | Overlapping malignant neoplasm of stomach | FALSE |
| CCI score | 535274723 | 198704 | Charlson - Cancer | TRUE | 1 | Hodgkin's disease of spleen | FALSE |
| CCI score | 535274723 | 200355 | Charlson - Cancer | TRUE | 1 | Hodgkin's disease of intra-abdominal lymph nodes | FALSE |
| CCI score | 535274723 | 200963 | Charlson - Cancer | TRUE | 1 | Primary malignant neoplasm of paraurethral glands | FALSE |
| CCI score | 535274723 | 254583 | Charlson - Cancer | TRUE | 1 | Kaposi's sarcoma of lung | FALSE |
| CCI score | 535274723 | 259748 | Charlson - Cancer | TRUE | 1 | Primary malignant neoplasm of accessory sinus | FALSE |
| CCI score | 535274723 | 313980 | Charlson - Cancer | TRUE | 1 | Hodgkin's disease, lymphocytic depletion of lymph nodes of multiple sites | FALSE |
| CCI score | 535274723 | 375479 | Charlson - Cancer | TRUE | 1 | Kaposi's sarcoma of soft tissue | FALSE |
| CCI score | 535274723 | 377811 | Charlson - Cancer | TRUE | 1 | Primary malignant neoplasm of retina (primary) | FALSE |
| CCI score | 535274723 | 432558 | Charlson - Cancer | TRUE | 1 | Overlapping malignant neoplasm of nasopharynx | FALSE |
| CCI score | 535274723 | 432845 | Charlson - Cancer | TRUE | 1 | Primary malignant neoplasm of central portion of female breast | FALSE |
| CCI score | 535274723 | 432848 | Charlson - Cancer | TRUE | 1 | Primary malignant neoplasm of cerebral ventricle | FALSE |
| CCI score | 535274723 | 433143 | Charlson - Cancer | TRUE | 1 | Primary malignant neoplasm of appendix | FALSE |
| CCI score | 535274723 | 433161 | Charlson - Cancer | TRUE | 1 | Hodgkin's disease, lymphocytic depletion of lymph nodes of head, face AND/OR neck | FALSE |
| CCI score | 535274723 | 433704 | Charlson - Cancer | TRUE | 1 | Primary malignant neoplasm of retromolar area | FALSE |
| CCI score | 535274723 | 434289 | Charlson - Cancer | TRUE | 1 | Primary malignant neoplasm of lingual tonsil | FALSE |
| CCI score | 535274723 | 434577 | Charlson - Cancer | TRUE | 1 | Primary malignant neoplasm of hypopharyngeal aspect of aryepiglottic fold | FALSE |
| CCI score | 535274723 | 434877 | Charlson - Cancer | TRUE | 1 | Hodgkin's disease of lymph nodes of axilla AND/OR upper limb | FALSE |
| CCI score | 535274723 | 435203 | Charlson - Cancer | TRUE | 1 | Hodgkin's disease, mixed cellularity of extranodal AND/OR solid organ site | FALSE |
| CCI score | 535274723 | 435493 | Charlson - Cancer | TRUE | 1 | Primary malignant neoplasm of short bone of upper limb | FALSE |
| CCI score | 535274723 | 436059 | Charlson - Cancer | TRUE | 1 | Multiple myeloma in remission | FALSE |
| CCI score | 535274723 | 436353 | Charlson - Cancer | TRUE | 1 | Primary malignant neoplasm of upper outer quadrant of female breast | FALSE |
| CCI score | 535274723 | 438080 | Charlson - Cancer | TRUE | 1 | Primary malignant neoplasm of anterior wall of nasopharynx | FALSE |
| CCI score | 535274723 | 438086 | Charlson - Cancer | TRUE | 1 | Primary malignant neoplasm of parietal lobe | FALSE |
| CCI score | 535274723 | 438095 | Charlson - Cancer | TRUE | 1 | Overlapping malignant neoplasm of eye and adnexa (primary) | FALSE |
| CCI score | 535274723 | 438370 | Charlson - Cancer | TRUE | 1 | Primary malignant neoplasm of glans penis | FALSE |
| CCI score | 535274723 | 438373 | Charlson - Cancer | TRUE | 1 | Hodgkin's disease, lymphocytic-histiocytic predominance of extranodal AND/OR solid organ site | FALSE |
| CCI score | 535274723 | 438693 | Charlson - Cancer | TRUE | 1 | Primary malignant neoplasm of mediastinum | FALSE |
| CCI score | 535274723 | 438694 | Charlson - Cancer | TRUE | 1 | Primary malignant neoplasm of hard palate | FALSE |
| CCI score | 535274723 | 438977 | Charlson - Cancer | TRUE | 1 | Primary malignant neoplasm of ribs and/or sternum and/or clavicle | FALSE |
| CCI score | 535274723 | 439293 | Charlson - Cancer | TRUE | 1 | Burkitt's lymphoma of spleen | FALSE |
| CCI score | 535274723 | 440345 | Charlson - Cancer | TRUE | 1 | Primary malignant neoplasm of lateral wall of oropharynx | FALSE |
| CCI score | 535274723 | 440649 | Charlson - Cancer | TRUE | 1 | Primary malignant neoplasm of head of pancreas | FALSE |
| CCI score | 535274723 | 440956 | Charlson - Cancer | TRUE | 1 | Primary malignant neoplasm of upper inner quadrant of female breast | FALSE |
| CCI score | 535274723 | 440965 | Charlson - Cancer | TRUE | 1 | Hodgkin's disease of intrathoracic lymph nodes | FALSE |
| CCI score | 535274723 | 441223 | Charlson - Cancer | TRUE | 1 | Primary malignant neoplasm of superior wall of nasopharynx | FALSE |
| CCI score | 535274723 | 441513 | Charlson - Cancer | TRUE | 1 | Primary malignant neoplasm of axillary tail of breast | FALSE |
| CCI score | 535274723 | 442169 | Charlson - Cancer | TRUE | 1 | Mycosis fungoides of intrapelvic lymph nodes | FALSE |
| CCI score | 535274723 | 4001172 | Charlson - Cancer | TRUE | 1 | Hepatoblastoma | FALSE |
| CCI score | 535274723 | 4002340 | Charlson - Cancer | TRUE | 1 | Primary malignant neoplasm of meninges | FALSE |
| CCI score | 535274723 | 4003184 | Charlson - Cancer | TRUE | 1 | Acute panmyelosis with myelofibrosis | FALSE |
| CCI score | 535274723 | 4003684 | Charlson - Cancer | TRUE | 1 | Overlapping malignant neoplasm of male breast | FALSE |
| CCI score | 535274723 | 4003830 | Charlson - Cancer | TRUE | 1 | Diffuse non-Hodgkin's lymphoma | FALSE |
| CCI score | 535274723 | 4038838 | Charlson - Cancer | TRUE | 1 | Non-Hodgkin's lymphoma (clinical) | FALSE |
| CCI score | 535274723 | 4038843 | Charlson - Cancer | TRUE | 1 | Hodgkin's disease, mixed cellularity (clinical) | FALSE |
| CCI score | 535274723 | 4092223 | Charlson - Cancer | TRUE | 1 | Malignant neoplasm of orbital bone | FALSE |
| CCI score | 535274723 | 4094260 | Charlson - Cancer | TRUE | 1 | Malignant neoplasm of peripheral nerves and autonomic nervous system | FALSE |
| CCI score | 535274723 | 4094548 | Charlson - Cancer | TRUE | 1 | Extramedullary plasmacytoma | FALSE |
| CCI score | 535274723 | 4094872 | Charlson - Cancer | TRUE | 1 | Malignant neoplasm, overlapping lesion of accessory sinuses | FALSE |
| CCI score | 535274723 | 4098597 | Charlson - Cancer | TRUE | 1 | Waldenström macroglobulinemia | FALSE |
| CCI score | 535274723 | 4111023 | Charlson - Cancer | TRUE | 1 | Malignant tumor of anus and anal canal | FALSE |
| CCI score | 535274723 | 4155171 | Charlson - Cancer | TRUE | 1 | Primary malignant neoplasm of lip | FALSE |
| CCI score | 535274723 | 4162859 | Charlson - Cancer | TRUE | 1 | Primary malignant neoplasm of adrenal medulla | FALSE |
| CCI score | 535274723 | 4170421 | Charlson - Cancer | TRUE | 1 | Gastrointestinal stromal tumor | FALSE |
| CCI score | 535274723 | 4217892 | Charlson - Cancer | TRUE | 1 | Hand-Schüller-Christian disease | FALSE |
| CCI score | 535274723 | 4246808 | Charlson - Cancer | TRUE | 1 | Primary malignant neoplasm of ciliary body (primary) | FALSE |
| CCI score | 535274723 | 4247336 | Charlson - Cancer | TRUE | 1 | Primary malignant neoplasm of middle ear | FALSE |
| CCI score | 535274723 | 4264693 | Charlson - Cancer | TRUE | 1 | Mast cell malignancy | FALSE |
| CCI score | 535274723 | 40481522 | Charlson - Cancer | TRUE | 1 | Primary mediastinal (thymic) large B-cell lymphoma | FALSE |
| CCI score | 535274723 | 40481524 | Charlson - Cancer | TRUE | 1 | Acute myeloid leukemia with t(9:11)(p22;q23); MLLT3-MLL | FALSE |
| CCI score | 535274723 | 40482893 | Charlson - Cancer | TRUE | 1 | Extranodal marginal zone B-cell lymphoma of mucosa-associated lymphoid tissue (MALT-lymphoma) | FALSE |
| CCI score | 535274723 | 40487528 | Charlson - Cancer | TRUE | 1 | Non-Hodgkin's lymphoma of extranodal site | FALSE |
| CCI score | 535274723 | 40492267 | Charlson - Cancer | TRUE | 1 | Gastrointestinal stromal tumor of small intestine | FALSE |
| CCI score | 535274723 | 45770892 | Charlson - Cancer | TRUE | 1 | Primary malignant neoplasm of uterus | FALSE |
| CCI score | 535274723 | 26052 | Charlson - Cancer | TRUE | 1 | Primary malignant neoplasm of larynx | FALSE |
| CCI score | 535274723 | 28083 | Charlson - Cancer | TRUE | 1 | Primary malignant neoplasm of pharynx | FALSE |
| CCI score | 535274723 | 76349 | Charlson - Cancer | TRUE | 1 | Primary malignant neoplasm of urinary system | FALSE |
| CCI score | 535274723 | 80045 | Charlson - Cancer | TRUE | 1 | Primary malignant neoplasm of anus | FALSE |
| CCI score | 535274723 | 132832 | Charlson - Cancer | TRUE | 1 | Primary malignant neoplasm of inner aspect of lip | FALSE |
| CCI score | 535274723 | 132853 | Charlson - Cancer | TRUE | 1 | Lymphoid leukemia | FALSE |
| CCI score | 535274723 | 133154 | Charlson - Cancer | TRUE | 1 | Plasma cell leukemia | FALSE |
| CCI score | 535274723 | 133419 | Charlson - Cancer | TRUE | 1 | Overlapping malignant melanoma of skin | FALSE |
| CCI score | 535274723 | 133424 | Charlson - Cancer | TRUE | 1 | Primary malignant neoplasm of thyroid gland | FALSE |
| CCI score | 535274723 | 134290 | Charlson - Cancer | TRUE | 1 | Primary malignant neoplasm of palate | FALSE |
| CCI score | 535274723 | 134305 | Charlson - Cancer | TRUE | 1 | Acute lymphoid leukemia | FALSE |
| CCI score | 535274723 | 134596 | Charlson - Cancer | TRUE | 1 | Lymphoid leukemia in remission | FALSE |
| CCI score | 535274723 | 135489 | Charlson - Cancer | TRUE | 1 | Primary malignant neoplasm of male breast | FALSE |
| CCI score | 535274723 | 135759 | Charlson - Cancer | TRUE | 1 | Sézary's disease of extranodal AND/OR solid organ site | FALSE |
| CCI score | 535274723 | 135766 | Charlson - Cancer | TRUE | 1 | Leukemia in remission | FALSE |
| CCI score | 535274723 | 136655 | Charlson - Cancer | TRUE | 1 | Hodgkin's disease, mixed cellularity of intrathoracic lymph nodes | FALSE |
| CCI score | 535274723 | 136915 | Charlson - Cancer | TRUE | 1 | Primary malignant neoplasm of skin of lip | FALSE |
| CCI score | 535274723 | 138378 | Charlson - Cancer | TRUE | 1 | Sézary's disease of lymph nodes of axilla AND/OR upper limb | FALSE |
| CCI score | 535274723 | 140664 | Charlson - Cancer | TRUE | 1 | Sézary's disease of lymph nodes of multiple sites | FALSE |
| CCI score | 535274723 | 141243 | Charlson - Cancer | TRUE | 1 | Hodgkin's disease, lymphocytic depletion of intrathoracic lymph nodes | FALSE |
| CCI score | 535274723 | 192560 | Charlson - Cancer | TRUE | 1 | Malignant lymphoma of lymph nodes of inguinal region AND/OR lower limb | FALSE |
| CCI score | 535274723 | 194589 | Charlson - Cancer | TRUE | 1 | Primary malignant neoplasm of biliary tract | FALSE |
| CCI score | 535274723 | 194593 | Charlson - Cancer | TRUE | 1 | Mycosis fungoides of lymph nodes of inguinal region AND/OR lower limb | FALSE |
| CCI score | 535274723 | 194876 | Charlson - Cancer | TRUE | 1 | Hodgkin's disease, lymphocytic-histiocytic predominance of intrapelvic lymph nodes | FALSE |
| CCI score | 535274723 | 195480 | Charlson - Cancer | TRUE | 1 | Primary malignant neoplasm of renal pelvis | FALSE |
| CCI score | 535274723 | 196055 | Charlson - Cancer | TRUE | 1 | Burkitt's lymphoma of lymph nodes of inguinal region and lower limb | FALSE |
| CCI score | 535274723 | 197225 | Charlson - Cancer | TRUE | 1 | Primary malignant neoplasm of clitoris | FALSE |
| CCI score | 535274723 | 197507 | Charlson - Cancer | TRUE | 1 | Primary malignant neoplasm of male genital organ | FALSE |
| CCI score | 535274723 | 198082 | Charlson - Cancer | TRUE | 1 | Overlapping malignant neoplasm of body of uterus | FALSE |
| CCI score | 535274723 | 198374 | Charlson - Cancer | TRUE | 1 | Nodular lymphoma of spleen | FALSE |
| CCI score | 535274723 | 200052 | Charlson - Cancer | TRUE | 1 | Primary malignant neoplasm of uterine adnexa | FALSE |
| CCI score | 535274723 | 200054 | Charlson - Cancer | TRUE | 1 | Primary malignant neoplasm of ureter | FALSE |
| CCI score | 535274723 | 201242 | Charlson - Cancer | TRUE | 1 | Hodgkin's disease, lymphocytic-histiocytic predominance of intra-abdominal lymph nodes | FALSE |
| CCI score | 535274723 | 201813 | Charlson - Cancer | TRUE | 1 | Mycosis fungoides of intra-abdominal lymph nodes | FALSE |
| CCI score | 535274723 | 257503 | Charlson - Cancer | TRUE | 1 | Primary malignant neoplasm of main bronchus | FALSE |
| CCI score | 535274723 | 260336 | Charlson - Cancer | TRUE | 1 | Primary malignant neoplasm of glottis | FALSE |
| CCI score | 535274723 | 312846 | Charlson - Cancer | TRUE | 1 | Kaposi's sarcoma of lymph nodes | FALSE |
| CCI score | 535274723 | 315763 | Charlson - Cancer | TRUE | 1 | Hodgkin's disease, nodular sclerosis of lymph nodes of multiple sites | FALSE |
| CCI score | 535274723 | 317510 | Charlson - Cancer | TRUE | 1 | Leukemia | FALSE |
| CCI score | 535274723 | 375490 | Charlson - Cancer | TRUE | 1 | Primary malignant neoplasm of cornea (primary) | FALSE |
| CCI score | 535274723 | 378081 | Charlson - Cancer | TRUE | 1 | Overlapping malignant neoplasm of soft tissues | FALSE |
| CCI score | 535274723 | 432271 | Charlson - Cancer | TRUE | 1 | Carcinoid tumor of ileum | FALSE |
| CCI score | 535274723 | 433709 | Charlson - Cancer | TRUE | 1 | Primary malignant neoplasm of tonsillar fossa | FALSE |
| CCI score | 535274723 | 433976 | Charlson - Cancer | TRUE | 1 | Primary malignant neoplasm of temporal lobe | FALSE |
| CCI score | 535274723 | 434592 | Charlson - Cancer | TRUE | 1 | B-cell lymphoma (clinical) | FALSE |
| CCI score | 535274723 | 435190 | Charlson - Cancer | TRUE | 1 | Primary malignant neoplasm of pyriform sinus | FALSE |
| CCI score | 535274723 | 435474 | Charlson - Cancer | TRUE | 1 | Primary malignant neoplasm of posterior hypopharyngeal wall | FALSE |
| CCI score | 535274723 | 435484 | Charlson - Cancer | TRUE | 1 | Primary malignant neoplasm of trigone of urinary bladder | FALSE |
| CCI score | 535274723 | 435485 | Charlson - Cancer | TRUE | 1 | Primary malignant neoplasm of ill-defined site | FALSE |
| CCI score | 535274723 | 435751 | Charlson - Cancer | TRUE | 1 | Primary malignant neoplasm of fundus of stomach | FALSE |
| CCI score | 535274723 | 436348 | Charlson - Cancer | TRUE | 1 | Primary malignant neoplasm of anal canal | FALSE |
| CCI score | 535274723 | 436635 | Charlson - Cancer | TRUE | 1 | Primary malignant neoplasm of sigmoid colon | FALSE |
| CCI score | 535274723 | 436640 | Charlson - Cancer | TRUE | 1 | Primary malignant neoplasm of long bone of lower limb | FALSE |
| CCI score | 535274723 | 436923 | Charlson - Cancer | TRUE | 1 | Primary malignant neoplasm of urinary bladder neck | FALSE |
| CCI score | 535274723 | 438692 | Charlson - Cancer | TRUE | 1 | Primary malignant neoplasm of lateral wall of nasopharynx | FALSE |
| CCI score | 535274723 | 438698 | Charlson - Cancer | TRUE | 1 | Malignant lymphoma of lymph nodes of head, face AND/OR neck | FALSE |
| CCI score | 535274723 | 439266 | Charlson - Cancer | TRUE | 1 | Sézary's disease of lymph nodes of inguinal region and lower limb | FALSE |
| CCI score | 535274723 | 439285 | Charlson - Cancer | TRUE | 1 | Hodgkin's disease, lymphocytic depletion of intra-abdominal lymph nodes | FALSE |
| CCI score | 535274723 | 440047 | Charlson - Cancer | TRUE | 1 | Primary malignant neoplasm of tonsillar pillar | FALSE |
| CCI score | 535274723 | 440335 | Charlson - Cancer | TRUE | 1 | Primary malignant neoplasm of lower gum | FALSE |
| CCI score | 535274723 | 441520 | Charlson - Cancer | TRUE | 1 | Primary malignant neoplasm of round ligament of uterus | FALSE |
| CCI score | 535274723 | 441805 | Charlson - Cancer | TRUE | 1 | Primary malignant neoplasm of endocervix | FALSE |
| CCI score | 535274723 | 441806 | Charlson - Cancer | TRUE | 1 | Primary malignant neoplasm of brain stem | FALSE |
| CCI score | 535274723 | 442151 | Charlson - Cancer | TRUE | 1 | Hodgkin's disease, lymphocytic depletion of lymph nodes of axilla AND/OR upper limb | FALSE |
| CCI score | 535274723 | 443380 | Charlson - Cancer | TRUE | 1 | Malignant tumor of thymus | FALSE |
| CCI score | 535274723 | 444203 | Charlson - Cancer | TRUE | 1 | Primary malignant neoplasm of articular cartilage | FALSE |
| CCI score | 535274723 | 4001329 | Charlson - Cancer | TRUE | 1 | Follicular non-Hodgkin's lymphoma, mixed small cleaved cell and large cell (clinical) | FALSE |
| CCI score | 535274723 | 4003028 | Charlson - Cancer | TRUE | 1 | Primary malignant neoplasm of descended testis | FALSE |
| CCI score | 535274723 | 4003029 | Charlson - Cancer | TRUE | 1 | Overlapping malignant neoplasm of vulva | FALSE |
| CCI score | 535274723 | 4038846 | Charlson - Cancer | TRUE | 1 | Langerhans cell histiocytosis, disseminated (clinical) | FALSE |
| CCI score | 535274723 | 4041799 | Charlson - Cancer | TRUE | 1 | Sézary's disease (clinical) | FALSE |
| CCI score | 535274723 | 4079282 | Charlson - Cancer | TRUE | 1 | Atypical chronic myeloid leukemia | FALSE |
| CCI score | 535274723 | 4079683 | Charlson - Cancer | TRUE | 1 | T-cell prolymphocytic leukemia | FALSE |
| CCI score | 535274723 | 4091486 | Charlson - Cancer | TRUE | 1 | Malignant neoplasm of overlapping lesion of urinary organs | FALSE |
| CCI score | 535274723 | 4095168 | Charlson - Cancer | TRUE | 1 | Malignant neoplasm, overlapping lesion of bladder | FALSE |
| CCI score | 535274723 | 4097283 | Charlson - Cancer | TRUE | 1 | Malignant neoplasm of peripheral nerves of lower limb, including hip | FALSE |
| CCI score | 535274723 | 4097561 | Charlson - Cancer | TRUE | 1 | Burkitt's lymphoma of lymph nodes of axilla and upper limb | FALSE |
| CCI score | 535274723 | 4114221 | Charlson - Cancer | TRUE | 1 | Malignant tumor of unknown origin | FALSE |
| CCI score | 535274723 | 4149851 | Charlson - Cancer | TRUE | 1 | Malignant melanoma of upper limb | FALSE |
| CCI score | 535274723 | 4173963 | Charlson - Cancer | TRUE | 1 | B-cell acute lymphoblastic leukemia | FALSE |
| CCI score | 535274723 | 4246802 | Charlson - Cancer | TRUE | 1 | Primary malignant neoplasm of bone of skull | FALSE |
| CCI score | 535274723 | 4300704 | Charlson - Cancer | TRUE | 1 | Diffuse large B-cell lymphoma (nodal/systemic with skin involvement) | FALSE |
| CCI score | 535274723 | 36684820 | Charlson - Cancer | TRUE | 1 | Primary malignant neoplasm of axillary tail of right female breast | FALSE |
| CCI score | 535274723 | 40481357 | Charlson - Cancer | TRUE | 1 | Large cell lymphoma of intrapelvic lymph nodes | FALSE |
| CCI score | 535274723 | 40481901 | Charlson - Cancer | TRUE | 1 | Mantle cell lymphoma | FALSE |
| CCI score | 535274723 | 40486896 | Charlson - Cancer | TRUE | 1 | Primary malignant neoplasm of extrahepatic bile duct | FALSE |
| CCI score | 535274723 | 40650479 | Charlson - Cancer | TRUE | 1 | Primary malignant neoplasm of posterior wall of urinary bladder | FALSE |
| CCI score | 535274723 | 24296 | Charlson - Cancer | TRUE | 1 | Primary malignant neoplasm of pituitary gland | FALSE |
| CCI score | 535274723 | 25748 | Charlson - Cancer | TRUE | 1 | Overlapping malignant neoplasm of esophagus | FALSE |
| CCI score | 535274723 | 75488 | Charlson - Cancer | TRUE | 1 | Primary malignant neoplasm of vertebral column | FALSE |
| CCI score | 535274723 | 81237 | Charlson - Cancer | TRUE | 1 | Primary malignant neoplasm of upper limb bones and scapula | FALSE |
| CCI score | 535274723 | 132841 | Charlson - Cancer | TRUE | 1 | Malignant lymphoma of lymph nodes of multiple sites | FALSE |
| CCI score | 535274723 | 132850 | Charlson - Cancer | TRUE | 1 | Myeloid leukemia in remission | FALSE |
| CCI score | 535274723 | 135491 | Charlson - Cancer | TRUE | 1 | Primary malignant neoplasm of spinal cord | FALSE |
| CCI score | 535274723 | 135496 | Charlson - Cancer | TRUE | 1 | Acute leukemia in remission | FALSE |
| CCI score | 535274723 | 136639 | Charlson - Cancer | TRUE | 1 | Primary malignant neoplasm of sphenoidal sinus | FALSE |
| CCI score | 535274723 | 138074 | Charlson - Cancer | TRUE | 1 | Primary malignant neoplasm of vermilion border of upper lip | FALSE |
| CCI score | 535274723 | 140046 | Charlson - Cancer | TRUE | 1 | Primary malignant neoplasm of ethmoidal sinus | FALSE |
| CCI score | 535274723 | 140057 | Charlson - Cancer | TRUE | 1 | Chronic leukemia | FALSE |
| CCI score | 535274723 | 140666 | Charlson - Cancer | TRUE | 1 | Myeloid leukemia | FALSE |
| CCI score | 535274723 | 140955 | Charlson - Cancer | TRUE | 1 | Overlapping malignant neoplasm of floor of mouth | FALSE |
| CCI score | 535274723 | 193155 | Charlson - Cancer | TRUE | 1 | Carcinoid tumor of large intestine | FALSE |
| CCI score | 535274723 | 193434 | Charlson - Cancer | TRUE | 1 | Hodgkin's disease, nodular sclerosis of intra-abdominal lymph nodes | FALSE |
| CCI score | 535274723 | 194594 | Charlson - Cancer | TRUE | 1 | Hodgkin's disease, mixed cellularity of intrapelvic lymph nodes | FALSE |
| CCI score | 535274723 | 194878 | Charlson - Cancer | TRUE | 1 | Nodular lymphoma of extranodal AND/OR solid organ site | FALSE |
| CCI score | 535274723 | 195483 | Charlson - Cancer | TRUE | 1 | Primary malignant neoplasm of penis | FALSE |
| CCI score | 535274723 | 196048 | Charlson - Cancer | TRUE | 1 | Primary malignant neoplasm of vagina | FALSE |
| CCI score | 535274723 | 197806 | Charlson - Cancer | TRUE | 1 | Primary malignant neoplasm of gallbladder | FALSE |
| CCI score | 535274723 | 198088 | Charlson - Cancer | TRUE | 1 | Nodular lymphoma of intrapelvic lymph nodes | FALSE |
| CCI score | 535274723 | 198091 | Charlson - Cancer | TRUE | 1 | Primary malignant neoplasm of retroperitoneum | FALSE |
| CCI score | 535274723 | 198092 | Charlson - Cancer | TRUE | 1 | Overlapping malignant neoplasm of retroperitoneum and peritoneum | FALSE |
| CCI score | 535274723 | 198988 | Charlson - Cancer | TRUE | 1 | Primary malignant neoplasm of pelvis | FALSE |
| CCI score | 535274723 | 201231 | Charlson - Cancer | TRUE | 1 | Malignant neoplasm of connective and soft tissue of pelvis | FALSE |
| CCI score | 535274723 | 201517 | Charlson - Cancer | TRUE | 1 | Primary malignant neoplasm of urachus | FALSE |
| CCI score | 535274723 | 201518 | Charlson - Cancer | TRUE | 1 | Primary malignant neoplasm of lower limb | FALSE |
| CCI score | 535274723 | 201519 | Charlson - Cancer | TRUE | 1 | Primary malignant neoplasm of liver | FALSE |
| CCI score | 535274723 | 201801 | Charlson - Cancer | TRUE | 1 | Primary malignant neoplasm of fallopian tube | FALSE |
| CCI score | 535274723 | 258375 | Charlson - Cancer | TRUE | 1 | Overlapping malignant neoplasm of bronchus and lung | FALSE |
| CCI score | 535274723 | 261236 | Charlson - Cancer | TRUE | 1 | Primary malignant neoplasm of upper lobe, bronchus or lung | FALSE |
| CCI score | 535274723 | 315481 | Charlson - Cancer | TRUE | 1 | Burkitt's lymphoma of lymph nodes of multiple sites | FALSE |
| CCI score | 535274723 | 372567 | Charlson - Cancer | TRUE | 1 | Primary malignant neoplasm of lacrimal gland | FALSE |
| CCI score | 535274723 | 379756 | Charlson - Cancer | TRUE | 1 | Primary malignant neoplasm of mandible | FALSE |
| CCI score | 535274723 | 432559 | Charlson - Cancer | TRUE | 1 | Primary malignant neoplasm of occipital lobe | FALSE |
| CCI score | 535274723 | 432837 | Charlson - Cancer | TRUE | 1 | Primary malignant neoplasm of cecum | FALSE |
| CCI score | 535274723 | 432838 | Charlson - Cancer | TRUE | 1 | Primary malignant neoplasm of cardia of stomach | FALSE |
| CCI score | 535274723 | 432843 | Charlson - Cancer | TRUE | 1 | Primary malignant neoplasm of tail of pancreas | FALSE |
| CCI score | 535274723 | 436054 | Charlson - Cancer | TRUE | 1 | Primary malignant neoplasm of undescended testis | FALSE |
| CCI score | 535274723 | 436344 | Charlson - Cancer | TRUE | 1 | Primary malignant neoplasm of posterior wall of nasopharynx | FALSE |
| CCI score | 535274723 | 437238 | Charlson - Cancer | TRUE | 1 | Carcinoid tumor of appendix | FALSE |
| CCI score | 535274723 | 438090 | Charlson - Cancer | TRUE | 1 | Overlapping malignant neoplasm of rectum, anus and anal canal | FALSE |
| CCI score | 535274723 | 438691 | Charlson - Cancer | TRUE | 1 | Overlapping malignant neoplasm of oropharynx | FALSE |
| CCI score | 535274723 | 439265 | Charlson - Cancer | TRUE | 1 | Sézary's disease of intrathoracic lymph nodes | FALSE |
| CCI score | 535274723 | 439267 | Charlson - Cancer | TRUE | 1 | Sézary's disease of intra-abdominal lymph nodes | FALSE |
| CCI score | 535274723 | 439746 | Charlson - Cancer | TRUE | 1 | Primary malignant neoplasm of hypopharynx | FALSE |
| CCI score | 535274723 | 440058 | Charlson - Cancer | TRUE | 1 | Malignant lymphoma of extranodal AND/OR solid organ site | FALSE |
| CCI score | 535274723 | 441515 | Charlson - Cancer | TRUE | 1 | Primary malignant neoplasm of lower outer quadrant of female breast | FALSE |
| CCI score | 535274723 | 442131 | Charlson - Cancer | TRUE | 1 | Primary malignant neoplasm of head | FALSE |
| CCI score | 535274723 | 442157 | Charlson - Cancer | TRUE | 1 | Hodgkin's disease, nodular sclerosis of lymph nodes of inguinal region AND/OR lower limb | FALSE |
| CCI score | 535274723 | 442159 | Charlson - Cancer | TRUE | 1 | Hodgkin's disease, mixed cellularity of spleen | FALSE |
| CCI score | 535274723 | 443381 | Charlson - Cancer | TRUE | 1 | Malignant tumor of sigmoid colon | FALSE |
| CCI score | 535274723 | 760936 | Charlson - Cancer | TRUE | 1 | Plasma cell leukemia in relapse | FALSE |
| CCI score | 535274723 | 4002357 | Charlson - Cancer | TRUE | 1 | Follicular non-Hodgkin's lymphoma, small cleaved cell (clinical) | FALSE |
| CCI score | 535274723 | 4003175 | Charlson - Cancer | TRUE | 1 | Primary malignant neoplasm of peripheral nerves of upper limb | FALSE |
| CCI score | 535274723 | 4003693 | Charlson - Cancer | TRUE | 1 | Overlapping malignant neoplasm of brain | FALSE |
| CCI score | 535274723 | 4038841 | Charlson - Cancer | TRUE | 1 | Lymphocyte-rich classical Hodgkin lymphoma | FALSE |
| CCI score | 535274723 | 4040379 | Charlson - Cancer | TRUE | 1 | Malignant mast cell tumor (clinical) | FALSE |
| CCI score | 535274723 | 4040380 | Charlson - Cancer | TRUE | 1 | Mycosis fungoides (clinical) | FALSE |
| CCI score | 535274723 | 4041800 | Charlson - Cancer | TRUE | 1 | Burkitt's lymphoma (clinical) | FALSE |
| CCI score | 535274723 | 4089777 | Charlson - Cancer | TRUE | 1 | Malignant neoplasm of connective and soft tissue of hip and lower limb | FALSE |
| CCI score | 535274723 | 4091467 | Charlson - Cancer | TRUE | 1 | Malignant neoplasm of axillary tail of female breast | FALSE |
| CCI score | 535274723 | 4092513 | Charlson - Cancer | TRUE | 1 | Malignant neoplasm, overlapping lesion of breast | FALSE |
| CCI score | 535274723 | 4094863 | Charlson - Cancer | TRUE | 1 | Malignant tumor of Meckel's diverticulum | FALSE |
| CCI score | 535274723 | 4095018 | Charlson - Cancer | TRUE | 1 | Malignant neoplasm of connective and soft tissue of abdomen | FALSE |
| CCI score | 535274723 | 4095589 | Charlson - Cancer | TRUE | 1 | Malignant melanoma of ear and/or external auditory canal | FALSE |
| CCI score | 535274723 | 4095592 | Charlson - Cancer | TRUE | 1 | Malignant melanoma of scalp and/or neck | FALSE |
| CCI score | 535274723 | 4115271 | Charlson - Cancer | TRUE | 1 | Sarcoma of liver | FALSE |
| CCI score | 535274723 | 4147164 | Charlson - Cancer | TRUE | 1 | Malignant tumor of lymphoid hemopoietic and related tissue | FALSE |
| CCI score | 535274723 | 4151263 | Charlson - Cancer | TRUE | 1 | Malignant melanoma of lower limb | FALSE |
| CCI score | 535274723 | 4160342 | Charlson - Cancer | TRUE | 1 | Neoplasm of eye region | FALSE |
| CCI score | 535274723 | 4187851 | Charlson - Cancer | TRUE | 1 | Primary malignant neoplasm of breast lower inner quadrant | FALSE |
| CCI score | 535274723 | 4189938 | Charlson - Cancer | TRUE | 1 | Acute monocytic/monoblastic leukemia | FALSE |
| CCI score | 535274723 | 4216139 | Charlson - Cancer | TRUE | 1 | Plasmacytoma | FALSE |
| CCI score | 535274723 | 4246127 | Charlson - Cancer | TRUE | 1 | Malignant neoplasm of liver | FALSE |
| CCI score | 535274723 | 4246141 | Charlson - Cancer | TRUE | 1 | Primary malignant neoplasm of pelvic bone | FALSE |
| CCI score | 535274723 | 4247836 | Charlson - Cancer | TRUE | 1 | Primary malignant neoplasm of major salivary gland | FALSE |
| CCI score | 535274723 | 4247842 | Charlson - Cancer | TRUE | 1 | Primary malignant neoplasm of myometrium | FALSE |
| CCI score | 535274723 | 4311499 | Charlson - Cancer | TRUE | 1 | Primary malignant neoplasm of respiratory tract | FALSE |
| CCI score | 535274723 | 4334322 | Charlson - Cancer | TRUE | 1 | Malignant neoplasm of lower respiratory tract | FALSE |
| CCI score | 535274723 | 37017321 | Charlson - Cancer | TRUE | 1 | Malignant carcinoid tumor of kidney | FALSE |
| CCI score | 535274723 | 37018875 | Charlson - Cancer | TRUE | 1 | Malignant neuroendocrine tumor | FALSE |
| CCI score | 535274723 | 40492268 | Charlson - Cancer | TRUE | 1 | Myelodysplastic/myeloproliferative disease | FALSE |
| CCI score | 535274723 | 45767695 | Charlson - Cancer | TRUE | 1 | Metastatic carcinoid tumor | FALSE |
| CCI score | 535274723 | 46271402 | Charlson - Cancer | TRUE | 1 | Malignant carcinoid tumor of small intestine | FALSE |
| CCI score | 535274723 | 27235 | Charlson - Cancer | TRUE | 1 | Primary malignant neoplasm of carotid body | FALSE |
| CCI score | 535274723 | 74582 | Charlson - Cancer | TRUE | 1 | Primary malignant neoplasm of rectum | FALSE |
| CCI score | 535274723 | 79749 | Charlson - Cancer | TRUE | 1 | Primary malignant neoplasm of body of penis | FALSE |
| CCI score | 535274723 | 80665 | Charlson - Cancer | TRUE | 1 | Malignant neoplasm of thorax | FALSE |
| CCI score | 535274723 | 81239 | Charlson - Cancer | TRUE | 1 | Primary malignant neoplasm of upper limb | FALSE |
| CCI score | 535274723 | 132258 | Charlson - Cancer | TRUE | 1 | Primary malignant neoplasm of frontal sinus | FALSE |
| CCI score | 535274723 | 133420 | Charlson - Cancer | TRUE | 1 | Primary malignant neoplasm of endocrine gland | FALSE |
| CCI score | 535274723 | 133710 | Charlson - Cancer | TRUE | 1 | Overlapping malignant neoplasm of lip | FALSE |
| CCI score | 535274723 | 134579 | Charlson - Cancer | TRUE | 1 | Primary malignant neoplasm of buccal mucosa | FALSE |
| CCI score | 535274723 | 134879 | Charlson - Cancer | TRUE | 1 | Mycosis fungoides of intrathoracic lymph nodes | FALSE |
| CCI score | 535274723 | 135204 | Charlson - Cancer | TRUE | 1 | Mycosis fungoides of extranodal AND/OR solid organ site | FALSE |
| CCI score | 535274723 | 137219 | Charlson - Cancer | TRUE | 1 | Primary malignant neoplasm of inner aspect of lower lip | FALSE |
| CCI score | 535274723 | 137800 | Charlson - Cancer | TRUE | 1 | Primary malignant neoplasm of maxillary sinus | FALSE |
| CCI score | 535274723 | 138351 | Charlson - Cancer | TRUE | 1 | Primary malignant neoplasm of inner aspect of upper lip | FALSE |
| CCI score | 535274723 | 138377 | Charlson - Cancer | TRUE | 1 | Mycosis fungoides of lymph nodes of head, face AND/OR neck | FALSE |
| CCI score | 535274723 | 140967 | Charlson - Cancer | TRUE | 1 | Myeloid sarcoma | FALSE |
| CCI score | 535274723 | 141816 | Charlson - Cancer | TRUE | 1 | Acute lymphoid leukemia in remission | FALSE |
| CCI score | 535274723 | 192261 | Charlson - Cancer | TRUE | 1 | Overlapping malignant neoplasm of pancreas | FALSE |
| CCI score | 535274723 | 193422 | Charlson - Cancer | TRUE | 1 | Primary malignant neoplasm of body of stomach | FALSE |
| CCI score | 535274723 | 195195 | Charlson - Cancer | TRUE | 1 | Malignant lymphoma of intrapelvic lymph nodes | FALSE |
| CCI score | 535274723 | 195482 | Charlson - Cancer | TRUE | 1 | Primary malignant neoplasm of intestinal tract | FALSE |
| CCI score | 535274723 | 195761 | Charlson - Cancer | TRUE | 1 | Hodgkin's disease of intrapelvic lymph nodes | FALSE |
| CCI score | 535274723 | 198381 | Charlson - Cancer | TRUE | 1 | Hodgkin's disease, lymphocytic-histiocytic predominance of spleen | FALSE |
| CCI score | 535274723 | 200051 | Charlson - Cancer | TRUE | 1 | Primary malignant neoplasm of ovary | FALSE |
| CCI score | 535274723 | 200343 | Charlson - Cancer | TRUE | 1 | Malignant lymphoma of spleen | FALSE |
| CCI score | 535274723 | 200349 | Charlson - Cancer | TRUE | 1 | Malignant lymphoma of intra-abdominal lymph nodes | FALSE |
| CCI score | 535274723 | 200962 | Charlson - Cancer | TRUE | 1 | Primary malignant neoplasm of prostate | FALSE |
| CCI score | 535274723 | 201238 | Charlson - Cancer | TRUE | 1 | Primary malignant neoplasm of female genital organ | FALSE |
| CCI score | 535274723 | 252840 | Charlson - Cancer | TRUE | 1 | Primary malignant neoplasm of upper respiratory tract | FALSE |
| CCI score | 535274723 | 254282 | Charlson - Cancer | TRUE | 1 | Primary malignant neoplasm of soft palate | FALSE |
| CCI score | 535274723 | 256633 | Charlson - Cancer | TRUE | 1 | Primary malignant neoplasm of base of tongue | FALSE |
| CCI score | 535274723 | 258981 | Charlson - Cancer | TRUE | 1 | Kaposi's sarcoma of skin | FALSE |
| CCI score | 535274723 | 261808 | Charlson - Cancer | TRUE | 1 | Primary malignant neoplasm of vestibule of mouth | FALSE |
| CCI score | 535274723 | 315202 | Charlson - Cancer | TRUE | 1 | Hodgkin's disease, mixed cellularity of lymph nodes of multiple sites | FALSE |
| CCI score | 535274723 | 320347 | Charlson - Cancer | TRUE | 1 | Nodular lymphoma of lymph nodes of multiple sites | FALSE |
| CCI score | 535274723 | 374874 | Charlson - Cancer | TRUE | 1 | Primary malignant neoplasm of eye | FALSE |
| CCI score | 535274723 | 376647 | Charlson - Cancer | TRUE | 1 | Primary malignant neoplasm of soft tissues | FALSE |
| CCI score | 535274723 | 380055 | Charlson - Cancer | TRUE | 1 | Primary malignant neoplasm of brain | FALSE |
| CCI score | 535274723 | 432260 | Charlson - Cancer | TRUE | 1 | Primary malignant neoplasm of upper third of esophagus | FALSE |
| CCI score | 535274723 | 432262 | Charlson - Cancer | TRUE | 1 | Primary malignant neoplasm of trachea | FALSE |
| CCI score | 535274723 | 432833 | Charlson - Cancer | TRUE | 1 | Primary malignant neoplasm of oropharynx | FALSE |
| CCI score | 535274723 | 434299 | Charlson - Cancer | TRUE | 1 | Hodgkin's disease, mixed cellularity of lymph nodes of axilla AND/OR upper limb | FALSE |
| CCI score | 535274723 | 434302 | Charlson - Cancer | TRUE | 1 | Hodgkin's disease, nodular sclerosis of extranodal AND/OR solid organ site | FALSE |
| CCI score | 535274723 | 434584 | Charlson - Cancer | TRUE | 1 | Kaposi's sarcoma (clinical) | FALSE |
| CCI score | 535274723 | 434590 | Charlson - Cancer | TRUE | 1 | Malignant melanoma of skin of eyelid | FALSE |
| CCI score | 535274723 | 434881 | Charlson - Cancer | TRUE | 1 | Peripheral T-cell lymphoma (clinical) | FALSE |
| CCI score | 535274723 | 435487 | Charlson - Cancer | TRUE | 1 | Primary malignant neoplasm of posterior mediastinum | FALSE |
| CCI score | 535274723 | 435752 | Charlson - Cancer | TRUE | 1 | Primary malignant neoplasm of duodenum | FALSE |
| CCI score | 535274723 | 435753 | Charlson - Cancer | TRUE | 1 | Malignant lymphoma of intrathoracic lymph nodes | FALSE |
| CCI score | 535274723 | 436042 | Charlson - Cancer | TRUE | 1 | Primary malignant neoplasm of anterior two-thirds of tongue | FALSE |
| CCI score | 535274723 | 436043 | Charlson - Cancer | TRUE | 1 | Overlapping malignant neoplasm of tongue | FALSE |
| CCI score | 535274723 | 436352 | Charlson - Cancer | TRUE | 1 | Primary malignant neoplasm of laryngeal cartilage | FALSE |
| CCI score | 535274723 | 436643 | Charlson - Cancer | TRUE | 1 | Primary malignant neoplasm of anterior aspect of epiglottis | FALSE |
| CCI score | 535274723 | 436651 | Charlson - Cancer | TRUE | 1 | Hodgkin's disease of extranodal AND/OR solid organ site | FALSE |
| CCI score | 535274723 | 437498 | Charlson - Cancer | TRUE | 1 | Primary malignant neoplasm of tongue | FALSE |
| CCI score | 535274723 | 437501 | Charlson - Cancer | TRUE | 1 | Primary malignant neoplasm of labia majora | FALSE |
| CCI score | 535274723 | 437504 | Charlson - Cancer | TRUE | 1 | Burkitt's tumor of lymph nodes of head, face AND/OR neck | FALSE |
| CCI score | 535274723 | 437818 | Charlson - Cancer | TRUE | 1 | Hodgkin's disease of lymph nodes of head, face AND/OR neck | FALSE |
| CCI score | 535274723 | 438089 | Charlson - Cancer | TRUE | 1 | Primary malignant neoplasm of pyloric antrum | FALSE |
| CCI score | 535274723 | 438360 | Charlson - Cancer | TRUE | 1 | Primary malignant neoplasm of vallecula | FALSE |
| CCI score | 535274723 | 438699 | Charlson - Cancer | TRUE | 1 | Primary malignant neoplasm of rectosigmoid junction | FALSE |
| CCI score | 535274723 | 439281 | Charlson - Cancer | TRUE | 1 | Hodgkin's disease, lymphocytic depletion of spleen | FALSE |
| CCI score | 535274723 | 439404 | Charlson - Cancer | TRUE | 1 | Primary malignant neoplasm of oral cavity | FALSE |
| CCI score | 535274723 | 440044 | Charlson - Cancer | TRUE | 1 | Overlapping malignant neoplasm of hypopharynx | FALSE |
| CCI score | 535274723 | 440351 | Charlson - Cancer | TRUE | 1 | Hodgkin's disease, lymphocytic-histiocytic predominance of intrathoracic lymph nodes | FALSE |
| CCI score | 535274723 | 441800 | Charlson - Cancer | TRUE | 1 | Primary malignant neoplasm of descending colon | FALSE |
| CCI score | 535274723 | 442150 | Charlson - Cancer | TRUE | 1 | Hodgkin's disease, lymphocytic depletion of lymph nodes of inguinal region AND/OR lower limb | FALSE |
| CCI score | 535274723 | 442162 | Charlson - Cancer | TRUE | 1 | Hodgkin's disease, lymphocytic-histiocytic predominance of lymph nodes of inguinal region AND/OR lower limb | FALSE |
| CCI score | 535274723 | 443384 | Charlson - Cancer | TRUE | 1 | Malignant tumor of transverse colon | FALSE |
| CCI score | 535274723 | 443719 | Charlson - Cancer | TRUE | 1 | Relapsing chronic myeloid leukemia | FALSE |
| CCI score | 535274723 | 4001170 | Charlson - Cancer | TRUE | 1 | Overlapping malignant neoplasm of palate | FALSE |
| CCI score | 535274723 | 4001328 | Charlson - Cancer | TRUE | 1 | Diffuse non-Hodgkin's lymphoma, lymphoblastic (clinical) | FALSE |
| CCI score | 535274723 | 4001664 | Charlson - Cancer | TRUE | 1 | Intrahepatic bile duct carcinoma | FALSE |
| CCI score | 535274723 | 4001666 | Charlson - Cancer | TRUE | 1 | Mesothelioma of peritoneum | FALSE |
| CCI score | 535274723 | 4002494 | Charlson - Cancer | TRUE | 1 | Histiocytic sarcoma (clinical) | FALSE |
| CCI score | 535274723 | 4002498 | Charlson - Cancer | TRUE | 1 | Overlapping malignant neoplasm of tonsil | FALSE |
| CCI score | 535274723 | 4003021 | Charlson - Cancer | TRUE | 1 | Angiosarcoma of liver | FALSE |
| CCI score | 535274723 | 4003179 | Charlson - Cancer | TRUE | 1 | Primary malignant neoplasm of peripheral nerves of trunk | FALSE |
| CCI score | 535274723 | 4003833 | Charlson - Cancer | TRUE | 1 | Follicular non-Hodgkin's lymphoma, large cell (clinical) | FALSE |
| CCI score | 535274723 | 4003834 | Charlson - Cancer | TRUE | 1 | Malignant immunoproliferative disease (clinical) | FALSE |
| CCI score | 535274723 | 4038842 | Charlson - Cancer | TRUE | 1 | Hodgkin's disease, nodular sclerosis (clinical) | FALSE |
| CCI score | 535274723 | 4082311 | Charlson - Cancer | TRUE | 1 | B-cell chronic lymphocytic leukemia | FALSE |
| CCI score | 535274723 | 4092358 | Charlson - Cancer | TRUE | 1 | Malignant neoplasm of connective and soft tissue of thorax | FALSE |
| CCI score | 535274723 | 4097284 | Charlson - Cancer | TRUE | 1 | Malignant neoplasm of peripheral nerve of pelvis | FALSE |
| CCI score | 535274723 | 4114198 | Charlson - Cancer | TRUE | 1 | Malignant tumor of olfactory tract | FALSE |
| CCI score | 535274723 | 4131761 | Charlson - Cancer | TRUE | 1 | Neoplasm of placenta | FALSE |
| CCI score | 535274723 | 4157454 | Charlson - Cancer | TRUE | 1 | Primary malignant neoplasm of lower lobe, bronchus or lung | FALSE |
| CCI score | 535274723 | 4187850 | Charlson - Cancer | TRUE | 1 | Primary malignant neoplasm of breast upper outer quadrant | FALSE |
| CCI score | 535274723 | 4225982 | Charlson - Cancer | TRUE | 1 | Mature T-cell AND/OR NK cell neoplasm | FALSE |
| CCI score | 535274723 | 4244051 | Charlson - Cancer | TRUE | 1 | Malignant melanoma of skin of breast | FALSE |
| CCI score | 535274723 | 4246029 | Charlson - Cancer | TRUE | 1 | Primary malignant neoplasm of cauda equina | FALSE |
| CCI score | 535274723 | 4311480 | Charlson - Cancer | TRUE | 1 | Primary malignant neoplasm of ileum | FALSE |
| CCI score | 535274723 | 4312698 | Charlson - Cancer | TRUE | 1 | Primary malignant neoplasm of trunk | FALSE |
| CCI score | 535274723 | 36716501 | Charlson - Cancer | TRUE | 1 | Primary malignant epithelial neoplasm of nasopharynx | FALSE |
| CCI score | 535274723 | 40479608 | Charlson - Cancer | TRUE | 1 | Mantle cell lymphoma of spleen | FALSE |
| CCI score | 535274723 | 40482847 | Charlson - Cancer | TRUE | 1 | Juvenile myelomonocytic leukemia | FALSE |
| CCI score | 535274723 | 40490328 | Charlson - Cancer | TRUE | 1 | Gastrointestinal stromal tumor of large intestine | FALSE |
| CCI score | 535274723 | 25486 | Charlson - Cancer | TRUE | 1 | Primary malignant neoplasm of islets of Langerhans | FALSE |
| CCI score | 535274723 | 76914 | Charlson - Cancer | TRUE | 1 | Primary malignant neoplasm of bone | FALSE |
| CCI score | 535274723 | 79740 | Charlson - Cancer | TRUE | 1 | Overlapping malignant neoplasm of colon | FALSE |
| CCI score | 535274723 | 133438 | Charlson - Cancer | TRUE | 1 | Chronic lymphoid leukemia in remission | FALSE |
| CCI score | 535274723 | 133713 | Charlson - Cancer | TRUE | 1 | Malignant melanoma of skin of face | FALSE |
| CCI score | 535274723 | 135762 | Charlson - Cancer | TRUE | 1 | Acute myeloid leukemia in remission | FALSE |
| CCI score | 535274723 | 135765 | Charlson - Cancer | TRUE | 1 | Sézary's disease of lymph nodes of head, face AND/OR neck | FALSE |
| CCI score | 535274723 | 138708 | Charlson - Cancer | TRUE | 1 | Acute leukemia | FALSE |
| CCI score | 535274723 | 140352 | Charlson - Cancer | TRUE | 1 | Acute myeloid leukemia, disease | FALSE |
| CCI score | 535274723 | 140950 | Charlson - Cancer | TRUE | 1 | Primary malignant neoplasm of gum | FALSE |
| CCI score | 535274723 | 141232 | Charlson - Cancer | TRUE | 1 | Malignant melanoma of skin | FALSE |
| CCI score | 535274723 | 192268 | Charlson - Cancer | TRUE | 1 | Hodgkin's disease, mixed cellularity of intra-abdominal lymph nodes | FALSE |
| CCI score | 535274723 | 193428 | Charlson - Cancer | TRUE | 1 | Burkitt's lymphoma of intrapelvic lymph nodes | FALSE |
| CCI score | 535274723 | 196360 | Charlson - Cancer | TRUE | 1 | Primary malignant neoplasm of bladder | FALSE |
| CCI score | 535274723 | 196645 | Charlson - Cancer | TRUE | 1 | Overlapping malignant neoplasm of male genital organs | FALSE |
| CCI score | 535274723 | 197506 | Charlson - Cancer | TRUE | 1 | Malignant neoplasm of abdomen | FALSE |
| CCI score | 535274723 | 198695 | Charlson - Cancer | TRUE | 1 | Overlapping malignant neoplasm of biliary tract | FALSE |
| CCI score | 535274723 | 198985 | Charlson - Cancer | TRUE | 1 | Primary malignant neoplasm of kidney | FALSE |
| CCI score | 535274723 | 200338 | Charlson - Cancer | TRUE | 1 | Nodular lymphoma of intra-abdominal lymph nodes | FALSE |
| CCI score | 535274723 | 256646 | Charlson - Cancer | TRUE | 1 | Primary malignant neoplasm of middle lobe, bronchus or lung | FALSE |
| CCI score | 535274723 | 259755 | Charlson - Cancer | TRUE | 1 | Primary malignant neoplasm of subglottis | FALSE |
| CCI score | 535274723 | 314600 | Charlson - Cancer | TRUE | 1 | Hodgkin's disease, nodular sclerosis of lymph nodes of axilla AND/OR upper limb | FALSE |
| CCI score | 535274723 | 432254 | Charlson - Cancer | TRUE | 1 | Primary malignant neoplasm of Waldeyer's ring | FALSE |
| CCI score | 535274723 | 432263 | Charlson - Cancer | TRUE | 1 | Primary malignant neoplasm of lower inner quadrant of female breast | FALSE |
| CCI score | 535274723 | 432844 | Charlson - Cancer | TRUE | 1 | Primary malignant neoplasm of anterior mediastinum | FALSE |
| CCI score | 535274723 | 434285 | Charlson - Cancer | TRUE | 1 | Primary malignant neoplasm of uvula | FALSE |
| CCI score | 535274723 | 434292 | Charlson - Cancer | TRUE | 1 | Primary malignant neoplasm of lesser curvature of stomach | FALSE |
| CCI score | 535274723 | 434588 | Charlson - Cancer | TRUE | 1 | Primary malignant neoplasm of parotid gland | FALSE |
| CCI score | 535274723 | 434880 | Charlson - Cancer | TRUE | 1 | Primary malignant neoplasm of short bone of lower limb | FALSE |
| CCI score | 535274723 | 435492 | Charlson - Cancer | TRUE | 1 | Nodular lymphoma of intrathoracic lymph nodes | FALSE |
| CCI score | 535274723 | 435758 | Charlson - Cancer | TRUE | 1 | Burkitt's tumor of extranodal AND/OR solid organ site | FALSE |
| CCI score | 535274723 | 436045 | Charlson - Cancer | TRUE | 1 | Malignant neoplasm of lip, oral cavity and pharynx | FALSE |
| CCI score | 535274723 | 436358 | Charlson - Cancer | TRUE | 1 | Primary malignant neoplasm of exocervix | FALSE |
| CCI score | 535274723 | 436913 | Charlson - Cancer | TRUE | 1 | Primary malignant neoplasm of jejunum | FALSE |
| CCI score | 535274723 | 436922 | Charlson - Cancer | TRUE | 1 | Primary malignant neoplasm of postcricoid region | FALSE |
| CCI score | 535274723 | 437233 | Charlson - Cancer | TRUE | 1 | Multiple myeloma | FALSE |
| CCI score | 535274723 | 437805 | Charlson - Cancer | TRUE | 1 | Primary malignant neoplasm of middle third of esophagus | FALSE |
| CCI score | 535274723 | 438982 | Charlson - Cancer | TRUE | 1 | Primary malignant neoplasm of anterior portion of floor of mouth | FALSE |
| CCI score | 535274723 | 439738 | Charlson - Cancer | TRUE | 1 | Primary malignant neoplasm of sublingual gland | FALSE |
| CCI score | 535274723 | 439739 | Charlson - Cancer | TRUE | 1 | Primary malignant neoplasm of posterior wall of oropharynx | FALSE |
| CCI score | 535274723 | 441225 | Charlson - Cancer | TRUE | 1 | Primary malignant neoplasm of ampulla of Vater | FALSE |
| CCI score | 535274723 | 441802 | Charlson - Cancer | TRUE | 1 | Primary malignant neoplasm of labia minora | FALSE |
| CCI score | 535274723 | 442122 | Charlson - Cancer | TRUE | 1 | Primary malignant neoplasm of paraganglion | FALSE |
| CCI score | 535274723 | 442134 | Charlson - Cancer | TRUE | 1 | Primary malignant neoplasm of craniopharyngeal duct | FALSE |
| CCI score | 535274723 | 442158 | Charlson - Cancer | TRUE | 1 | Hodgkin's disease, nodular sclerosis of lymph nodes of head, face AND/OR neck | FALSE |
| CCI score | 535274723 | 443391 | Charlson - Cancer | TRUE | 1 | Malignant tumor of cecum | FALSE |
| CCI score | 535274723 | 443565 | Charlson - Cancer | TRUE | 1 | Carcinoid tumor of gastrointestinal tract | FALSE |
| CCI score | 535274723 | 4002497 | Charlson - Cancer | TRUE | 1 | Acute promyelocytic leukemia, FAB M3 | FALSE |
| CCI score | 535274723 | 4003027 | Charlson - Cancer | TRUE | 1 | Overlapping malignant neoplasm of penis | FALSE |
| CCI score | 535274723 | 4003188 | Charlson - Cancer | TRUE | 1 | Adult T-cell leukemia/lymphoma | FALSE |
| CCI score | 535274723 | 4003674 | Charlson - Cancer | TRUE | 1 | Primary malignant neoplasm of branchial cleft | FALSE |
| CCI score | 535274723 | 4003836 | Charlson - Cancer | TRUE | 1 | Immunoproliferative small intestinal disease (clinical) | FALSE |
| CCI score | 535274723 | 4033318 | Charlson - Cancer | TRUE | 1 | Overlapping malignant neoplasm of bone and articular cartilage of limbs | FALSE |
| CCI score | 535274723 | 4038835 | Charlson - Cancer | TRUE | 1 | Hodgkin's disease (clinical) | FALSE |
| CCI score | 535274723 | 4044708 | Charlson - Cancer | TRUE | 1 | Langerhans cell histiocytosis, unifocal (clinical) | FALSE |
| CCI score | 535274723 | 4054513 | Charlson - Cancer | TRUE | 1 | Neoplasm of accessory sinus | FALSE |
| CCI score | 535274723 | 4091768 | Charlson - Cancer | TRUE | 1 | Burkitt's lymphoma of intrathoracic lymph nodes | FALSE |
| CCI score | 535274723 | 4092515 | Charlson - Cancer | TRUE | 1 | Malignant neoplasm, overlapping lesion of cervix uteri | FALSE |
| CCI score | 535274723 | 4097560 | Charlson - Cancer | TRUE | 1 | Burkitt's lymphoma of intra-abdominal lymph nodes | FALSE |
| CCI score | 535274723 | 4110889 | Charlson - Cancer | TRUE | 1 | Malignant tumor of acoustic vestibular nerve | FALSE |
| CCI score | 535274723 | 4111917 | Charlson - Cancer | TRUE | 1 | Malignant mesothelioma of pleura | FALSE |
| CCI score | 535274723 | 4133599 | Charlson - Cancer | TRUE | 1 | Chronic myelomonocytic leukemia | FALSE |
| CCI score | 535274723 | 4139358 | Charlson - Cancer | TRUE | 1 | Erythroleukemia, FAB M6 in remission | FALSE |
| CCI score | 535274723 | 4153890 | Charlson - Cancer | TRUE | 1 | Malignant melanoma of trunk | FALSE |
| CCI score | 535274723 | 4162115 | Charlson - Cancer | TRUE | 1 | Primary malignant neoplasm of adrenal cortex | FALSE |
| CCI score | 535274723 | 4162253 | Charlson - Cancer | TRUE | 1 | Primary malignant neoplasm of breast | FALSE |
| CCI score | 535274723 | 4162860 | Charlson - Cancer | TRUE | 1 | Primary malignant neoplasm of body of uterus | FALSE |
| CCI score | 535274723 | 4188545 | Charlson - Cancer | TRUE | 1 | Primary malignant neoplasm of breast lower outer quadrant | FALSE |
| CCI score | 535274723 | 4247238 | Charlson - Cancer | TRUE | 1 | Primary malignant neoplasm of endometrium | FALSE |
| CCI score | 535274723 | 4288751 | Charlson - Cancer | TRUE | 1 | Heavy chain disease | FALSE |
| CCI score | 535274723 | 4301668 | Charlson - Cancer | TRUE | 1 | Primary cutaneous follicular center B-cell lymphoma | FALSE |
| CCI score | 535274723 | 36684817 | Charlson - Cancer | TRUE | 1 | Primary malignant neoplasm of axillary tail of left female breast | FALSE |
| CCI score | 535274723 | 37396742 | Charlson - Cancer | TRUE | 1 | Aggressive systemic mastocytosis | FALSE |
| CCI score | 535274723 | 40482859 | Charlson - Cancer | TRUE | 1 | Malignant carcinoid tumor | FALSE |
| CCI score | 535274723 | 40483761 | Charlson - Cancer | TRUE | 1 | Acute myeloid leukemia with myelodysplasia-related changes | FALSE |
| CCI score | 535274723 | 40486171 | Charlson - Cancer | TRUE | 1 | Diffuse follicle center lymphoma | FALSE |
| CCI score | 535274723 | 40488812 | Charlson - Cancer | TRUE | 1 | Sarcoma of dendritic cells (accessory cells) | FALSE |
| CCI score | 535274723 | 46271647 | Charlson - Cancer | TRUE | 1 | Malignant carcinoid tumor of stomach | FALSE |
| CCI score | 535274723 | 26638 | Charlson - Cancer | TRUE | 1 | Primary malignant neoplasm of esophagus | FALSE |
| CCI score | 535274723 | 132565 | Charlson - Cancer | TRUE | 1 | Primary malignant neoplasm of vermilion border of lower lip | FALSE |
| CCI score | 535274723 | 132575 | Charlson - Cancer | TRUE | 1 | Myeloid sarcoma in remission | FALSE |
| CCI score | 535274723 | 134295 | Charlson - Cancer | TRUE | 1 | Primary malignant neoplasm of spinal meninges | FALSE |
| CCI score | 535274723 | 134597 | Charlson - Cancer | TRUE | 1 | Chronic leukemia in remission | FALSE |
| CCI score | 535274723 | 135750 | Charlson - Cancer | TRUE | 1 | Primary malignant neoplasm of floor of mouth | FALSE |
| CCI score | 535274723 | 136917 | Charlson - Cancer | TRUE | 1 | Primary malignant neoplasm of skin of face | FALSE |
| CCI score | 535274723 | 138099 | Charlson - Cancer | TRUE | 1 | Erythroleukemia, FAB M6 | FALSE |
| CCI score | 535274723 | 139750 | Charlson - Cancer | TRUE | 1 | Primary malignant neoplasm of skin | FALSE |
| CCI score | 535274723 | 192255 | Charlson - Cancer | TRUE | 1 | Primary malignant neoplasm of pylorus | FALSE |
| CCI score | 535274723 | 193719 | Charlson - Cancer | TRUE | 1 | Primary malignant neoplasm of spermatic cord | FALSE |
| CCI score | 535274723 | 196051 | Charlson - Cancer | TRUE | 1 | Overlapping malignant neoplasm of female genital organs | FALSE |
| CCI score | 535274723 | 197799 | Charlson - Cancer | TRUE | 1 | Primary malignant neoplasm of the peritoneum | FALSE |
| CCI score | 535274723 | 198100 | Charlson - Cancer | TRUE | 1 | Carcinoid tumor of small intestine | FALSE |
| CCI score | 535274723 | 198104 | Charlson - Cancer | TRUE | 1 | Primary malignant neoplasm of adrenal gland | FALSE |
| CCI score | 535274723 | 199754 | Charlson - Cancer | TRUE | 1 | Primary malignant neoplasm of pancreas | FALSE |
| CCI score | 535274723 | 255192 | Charlson - Cancer | TRUE | 1 | Primary malignant neoplasm of commissure of lip | FALSE |
| CCI score | 535274723 | 261514 | Charlson - Cancer | TRUE | 1 | Primary malignant neoplasm of supraglottis | FALSE |
| CCI score | 535274723 | 321234 | Charlson - Cancer | TRUE | 1 | Overlapping malignant neoplasm of heart, mediastinum and pleura | FALSE |
| CCI score | 535274723 | 321522 | Charlson - Cancer | TRUE | 1 | Hodgkin's disease, lymphocytic-histiocytic predominance of lymph nodes of multiple sites | FALSE |
| CCI score | 535274723 | 373151 | Charlson - Cancer | TRUE | 1 | Primary malignant neoplasm of conjunctiva (primary) | FALSE |
| CCI score | 535274723 | 432257 | Charlson - Cancer | TRUE | 1 | Primary malignant neoplasm of transverse colon | FALSE |
| CCI score | 535274723 | 432267 | Charlson - Cancer | TRUE | 1 | Hodgkin's disease, lymphocytic depletion of extranodal AND/OR solid organ site | FALSE |
| CCI score | 535274723 | 433149 | Charlson - Cancer | TRUE | 1 | Primary malignant neoplasm of cerebellum | FALSE |
| CCI score | 535274723 | 434300 | Charlson - Cancer | TRUE | 1 | Neuroendocrine tumor | FALSE |
| CCI score | 535274723 | 436652 | Charlson - Cancer | TRUE | 1 | Hodgkin's disease of lymph nodes of inguinal region AND/OR lower limb | FALSE |
| CCI score | 535274723 | 437220 | Charlson - Cancer | TRUE | 1 | Primary malignant neoplasm of ventral surface of tongue | FALSE |
| CCI score | 535274723 | 440339 | Charlson - Cancer | TRUE | 1 | Primary malignant neoplasm of prepuce | FALSE |
| CCI score | 535274723 | 440658 | Charlson - Cancer | TRUE | 1 | Primary malignant neoplasm of skin of upper limb | FALSE |
| CCI score | 535274723 | 441233 | Charlson - Cancer | TRUE | 1 | Primary malignant neoplasm of frontal lobe | FALSE |
| CCI score | 535274723 | 442161 | Charlson - Cancer | TRUE | 1 | Hodgkin's disease, mixed cellularity of lymph nodes of head, face AND/OR neck | FALSE |
| CCI score | 535274723 | 442168 | Charlson - Cancer | TRUE | 1 | Mycosis fungoides of spleen | FALSE |
| CCI score | 535274723 | 4002356 | Charlson - Cancer | TRUE | 1 | Diffuse non-Hodgkin's lymphoma, small cell (clinical) | FALSE |
| CCI score | 535274723 | 4032870 | Charlson - Cancer | TRUE | 1 | Malignant mesothelioma of pericardium | FALSE |
| CCI score | 535274723 | 4033891 | Charlson - Cancer | TRUE | 1 | Mesothelioma (malignant, clinical disorder) | FALSE |
| CCI score | 535274723 | 4041798 | Charlson - Cancer | TRUE | 1 | Hodgkin's disease, lymphocytic depletion (clinical) | FALSE |
| CCI score | 535274723 | 4091469 | Charlson - Cancer | TRUE | 1 | Malignant neoplasm of nipple and areola of male breast | FALSE |
| CCI score | 535274723 | 4091621 | Charlson - Cancer | TRUE | 1 | Malignant neoplasm of peripheral nerve of abdomen | FALSE |
| CCI score | 535274723 | 4092235 | Charlson - Cancer | TRUE | 1 | Malignant neoplasm of connective and soft tissue of upper limb and shoulder | FALSE |
| CCI score | 535274723 | 4094262 | Charlson - Cancer | TRUE | 1 | Malignant neoplasm of peripheral nerve of thorax | FALSE |
| CCI score | 535274723 | 4095892 | Charlson - Cancer | TRUE | 1 | Malignant neoplasm of peripheral nerves of head, face and neck | FALSE |
| CCI score | 535274723 | 4137687 | Charlson - Cancer | TRUE | 1 | Acute promyelocytic leukemia, FAB M3, in remission | FALSE |
| CCI score | 535274723 | 4158563 | Charlson - Cancer | TRUE | 1 | Primary malignant neoplasm of breast upper inner quadrant | FALSE |
| CCI score | 535274723 | 4179720 | Charlson - Cancer | TRUE | 1 | Primary malignant neoplasm of gastrointestinal tract | FALSE |
| CCI score | 535274723 | 4247822 | Charlson - Cancer | TRUE | 1 | Primary malignant neoplasm of central nervous system | FALSE |
| CCI score | 535274723 | 4308811 | Charlson - Cancer | TRUE | 1 | Neoplasm of soft tissue | FALSE |
| CCI score | 535274723 | 40481907 | Charlson - Cancer | TRUE | 1 | Carcinoid tumor | FALSE |
| CCI score | 535274723 | 40488896 | Charlson - Cancer | TRUE | 1 | Anaplastic large cell lymphoma, ALK negative | FALSE |
| CCI score | 535274723 | 42709931 | Charlson - Cancer | TRUE | 1 | Primary malignant neoplasm of dome of urinary bladder | FALSE |
| CCI score | 535274723 | 45765770 | Charlson - Cancer | TRUE | 1 | Follicular non-Hodgkin's lymphoma diffuse follicle center sub-type grade 1 | FALSE |
| CCI score | 359043664 | 321319 | Charlson - CHF | TRUE | 1 | Cardiomyopathy | FALSE |
| CCI score | 359043664 | 4004279 | Charlson - CHF | TRUE | 1 | High output heart failure | FALSE |
| CCI score | 359043664 | 4242669 | Charlson - CHF | TRUE | 1 | Biventricular congestive heart failure | FALSE |
| CCI score | 359043664 | 40480603 | Charlson - CHF | TRUE | 1 | Acute systolic heart failure | FALSE |
| CCI score | 359043664 | 318773 | Charlson - CHF | TRUE | 1 | Dilated cardiomyopathy secondary to alcohol | FALSE |
| CCI score | 359043664 | 439696 | Charlson - CHF | TRUE | 1 | Hypertensive heart and renal disease with (congestive) heart failure | FALSE |
| CCI score | 359043664 | 443580 | Charlson - CHF | TRUE | 1 | Systolic heart failure | FALSE |
| CCI score | 359043664 | 443587 | Charlson - CHF | TRUE | 1 | Diastolic heart failure | FALSE |
| CCI score | 359043664 | 444101 | Charlson - CHF | TRUE | 1 | Hypertensive heart failure | FALSE |
| CCI score | 359043664 | 4014159 | Charlson - CHF | TRUE | 1 | Chronic right-sided heart failure | FALSE |
| CCI score | 359043664 | 4172864 | Charlson - CHF | TRUE | 1 | Neonatal cardiac failure | FALSE |
| CCI score | 359043664 | 4163710 | Charlson - CHF | TRUE | 1 | Dilated cardiomyopathy | FALSE |
| CCI score | 359043664 | 4229440 | Charlson - CHF | TRUE | 1 | Chronic congestive heart failure | FALSE |
| CCI score | 359043664 | 4233424 | Charlson - CHF | TRUE | 1 | Acute right-sided heart failure | FALSE |
| CCI score | 359043664 | 44782718 | Charlson - CHF | TRUE | 1 | Acute combined systolic and diastolic heart failure | FALSE |
| CCI score | 359043664 | 44782733 | Charlson - CHF | TRUE | 1 | Acute on chronic combined systolic and diastolic heart failure | FALSE |
| CCI score | 359043664 | 320746 | Charlson - CHF | TRUE | 1 | Cardiomyopathy associated with another disorder | FALSE |
| CCI score | 359043664 | 439846 | Charlson - CHF | TRUE | 1 | Left heart failure | FALSE |
| CCI score | 359043664 | 4190773 | Charlson - CHF | TRUE | 1 | Restrictive cardiomyopathy | FALSE |
| CCI score | 359043664 | 4273632 | Charlson - CHF | TRUE | 1 | Right ventricular failure | FALSE |
| CCI score | 359043664 | 40479576 | Charlson - CHF | TRUE | 1 | Chronic diastolic heart failure | FALSE |
| CCI score | 359043664 | 40481043 | Charlson - CHF | TRUE | 1 | Acute on chronic diastolic heart failure | FALSE |
| CCI score | 359043664 | 316139 | Charlson - CHF | TRUE | 1 | Heart failure | FALSE |
| CCI score | 359043664 | 40480602 | Charlson - CHF | TRUE | 1 | Acute on chronic systolic heart failure | FALSE |
| CCI score | 359043664 | 44782719 | Charlson - CHF | TRUE | 1 | Chronic combined systolic and diastolic heart failure | FALSE |
| CCI score | 359043664 | 319835 | Charlson - CHF | TRUE | 1 | Congestive heart failure | FALSE |
| CCI score | 359043664 | 4110961 | Charlson - CHF | TRUE | 1 | Generalized ischemic myocardial dysfunction | FALSE |
| CCI score | 359043664 | 4195785 | Charlson - CHF | TRUE | 1 | Right heart failure secondary to left heart failure | FALSE |
| CCI score | 359043664 | 37309625 | Charlson - CHF | TRUE | 1 | Acute on chronic right-sided congestive heart failure | FALSE |
| CCI score | 359043664 | 40479192 | Charlson - CHF | TRUE | 1 | Chronic systolic heart failure | FALSE |
| CCI score | 359043664 | 40481042 | Charlson - CHF | TRUE | 1 | Acute diastolic heart failure | FALSE |
| CCI score | 359043664 | 40482727 | Charlson - CHF | TRUE | 1 | Combined systolic and diastolic dysfunction | FALSE |
| CCI score | 78746470 | 4182210 | Charlson - Dementia | TRUE | 1 | Dementia | FALSE |
| CCI score | 78746470 | 37109056 | Charlson - Dementia | TRUE | 1 | Vascular dementia without behavioral disturbance | FALSE |
| CCI score | 78746470 | 43530666 | Charlson - Dementia | TRUE | 1 | Dementia with behavioral disturbance | FALSE |
| CCI score | 78746470 | 373179 | Charlson - Dementia | TRUE | 1 | Senile degeneration of brain | FALSE |
| CCI score | 78746470 | 378419 | Charlson - Dementia | TRUE | 1 | Alzheimer's disease | FALSE |
| CCI score | 78746470 | 4220313 | Charlson - Dementia | TRUE | 1 | Primary degenerative dementia of the Alzheimer type, senile onset | FALSE |
| CCI score | 78746470 | 37018688 | Charlson - Dementia | TRUE | 1 | Vascular dementia with behavioral disturbance | FALSE |
| CCI score | 78746470 | 4218017 | Charlson - Dementia | TRUE | 1 | Primary degenerative dementia of the Alzheimer type, presenile onset | FALSE |
| CCI score | 78746470 | 374888 | Charlson - Dementia | TRUE | 1 | Dementia associated with another disease | FALSE |
| CCI score | 78746470 | 443605 | Charlson - Dementia | TRUE | 1 | Vascular dementia | FALSE |
| CCI score | 719585646 | 439770 | Charlson - DM | TRUE | 1 | Ketoacidosis due to type 1 diabetes mellitus | FALSE |
| CCI score | 719585646 | 443412 | Charlson - DM | TRUE | 1 | Type 1 diabetes mellitus without complication | FALSE |
| CCI score | 719585646 | 4214376 | Charlson - DM | TRUE | 1 | Hyperglycemia | FALSE |
| CCI score | 719585646 | 4224254 | Charlson - DM | TRUE | 1 | Ketoacidotic coma due to type 1 diabetes mellitus | FALSE |
| CCI score | 719585646 | 4228443 | Charlson - DM | TRUE | 1 | Ketoacidotic coma due to type 2 diabetes mellitus | FALSE |
| CCI score | 719585646 | 442793 | Charlson - DM | TRUE | 1 | Complication due to diabetes mellitus | FALSE |
| CCI score | 719585646 | 4008576 | Charlson - DM | TRUE | 1 | Diabetes mellitus without complication | FALSE |
| CCI score | 719585646 | 4152858 | Charlson - DM | TRUE | 1 | Type 1 diabetes mellitus with arthropathy | FALSE |
| CCI score | 719585646 | 43531616 | Charlson - DM | TRUE | 1 | Dermopathy due to type 2 diabetes mellitus | FALSE |
| CCI score | 719585646 | 45757363 | Charlson - DM | TRUE | 1 | Hypoglycemia due to type 2 diabetes mellitus | FALSE |
| CCI score | 719585646 | 45769832 | Charlson - DM | TRUE | 1 | Dermopathy due to type 1 diabetes mellitus | FALSE |
| CCI score | 719585646 | 4029423 | Charlson - DM | TRUE | 1 | Hypoglycemia due to diabetes mellitus | FALSE |
| CCI score | 719585646 | 4032787 | Charlson - DM | TRUE | 1 | Hyperosmolarity | FALSE |
| CCI score | 719585646 | 4042502 | Charlson - DM | TRUE | 1 | Disease of mouth | FALSE |
| CCI score | 719585646 | 4099214 | Charlson - DM | TRUE | 1 | Type 1 diabetes mellitus with ulcer | FALSE |
| CCI score | 719585646 | 4228112 | Charlson - DM | TRUE | 1 | Hypoglycemic coma due to type 1 diabetes mellitus | FALSE |
| CCI score | 719585646 | 201530 | Charlson - DM | TRUE | 1 | Hyperosmolar coma due to type 2 diabetes mellitus | FALSE |
| CCI score | 719585646 | 201820 | Charlson - DM | TRUE | 1 | Diabetes mellitus | FALSE |
| CCI score | 719585646 | 4099651 | Charlson - DM | TRUE | 1 | Type 2 diabetes mellitus with ulcer | FALSE |
| CCI score | 719585646 | 4159742 | Charlson - DM | TRUE | 1 | Diabetic foot ulcer | FALSE |
| CCI score | 719585646 | 4226798 | Charlson - DM | TRUE | 1 | Hypoglycemic coma due to diabetes mellitus | FALSE |
| CCI score | 719585646 | 37016349 | Charlson - DM | TRUE | 1 | Hyperglycemia due to type 2 diabetes mellitus | FALSE |
| CCI score | 719585646 | 45770902 | Charlson - DM | TRUE | 1 | Ulcer of lower limb due to type 1 diabetes mellitus | FALSE |
| CCI score | 719585646 | 46270483 | Charlson - DM | TRUE | 1 | Arthropathy due to metabolic disorder | FALSE |
| CCI score | 719585646 | 134398 | Charlson - DM | TRUE | 1 | Periodontal disease | FALSE |
| CCI score | 719585646 | 443734 | Charlson - DM | TRUE | 1 | Ketoacidosis due to type 2 diabetes mellitus | FALSE |
| CCI score | 719585646 | 4033942 | Charlson - DM | TRUE | 1 | Diabetic dermopathy | FALSE |
| CCI score | 719585646 | 4095288 | Charlson - DM | TRUE | 1 | Ketoacidotic coma due to diabetes mellitus | FALSE |
| CCI score | 719585646 | 4198296 | Charlson - DM | TRUE | 1 | Type 2 diabetes mellitus with neuropathic arthropathy | FALSE |
| CCI score | 719585646 | 43530690 | Charlson - DM | TRUE | 1 | Foot ulcer due to type 2 diabetes mellitus | FALSE |
| CCI score | 719585646 | 4009303 | Charlson - DM | TRUE | 1 | Diabetic ketoacidosis without coma | FALSE |
| CCI score | 719585646 | 4114427 | Charlson - DM | TRUE | 1 | Neuropathic arthropathy due to diabetes mellitus | FALSE |
| CCI score | 719585646 | 4193704 | Charlson - DM | TRUE | 1 | Type 2 diabetes mellitus without complication | FALSE |
| CCI score | 719585646 | 4227657 | Charlson - DM | TRUE | 1 | Skin ulcer due to diabetes mellitus | FALSE |
| CCI score | 719585646 | 37016348 | Charlson - DM | TRUE | 1 | Hyperglycemia due to type 1 diabetes mellitus | FALSE |
| CCI score | 719585646 | 45769830 | Charlson - DM | TRUE | 1 | Neuropathic arthropathy due to type 1 diabetes mellitus | FALSE |
| CCI score | 719585646 | 201254 | Charlson - DM | TRUE | 1 | Type 1 diabetes mellitus | FALSE |
| CCI score | 719585646 | 201826 | Charlson - DM | TRUE | 1 | Type 2 diabetes mellitus | FALSE |
| CCI score | 719585646 | 443727 | Charlson - DM | TRUE | 1 | Diabetic ketoacidosis | FALSE |
| CCI score | 719585646 | 4196141 | Charlson - DM | TRUE | 1 | Arthropathy due to type 2 diabetes mellitus | FALSE |
| CCI score | 719585646 | 4226238 | Charlson - DM | TRUE | 1 | Hyperosmolar coma due to diabetes mellitus | FALSE |
| CCI score | 719585646 | 36714116 | Charlson - DM | TRUE | 1 | Hypoglycemic coma due to type 2 diabetes mellitus | FALSE |
| CCI score | 719585646 | 45769876 | Charlson - DM | TRUE | 1 | Hypoglycemia due to type 1 diabetes mellitus | FALSE |
| CCI score | 403438288 | 192279 | Charlson - DMcx | TRUE | 1 | Disorder of kidney due to diabetes mellitus | FALSE |
| CCI score | 403438288 | 200687 | Charlson - DMcx | TRUE | 1 | Renal disorder due to type 1 diabetes mellitus | FALSE |
| CCI score | 403438288 | 318712 | Charlson - DMcx | TRUE | 1 | Peripheral circulatory disorder due to type 1 diabetes mellitus | FALSE |
| CCI score | 403438288 | 378743 | Charlson - DMcx | TRUE | 1 | Mild nonproliferative retinopathy due to diabetes mellitus | FALSE |
| CCI score | 403438288 | 4143857 | Charlson - DMcx | TRUE | 1 | Lumbosacral radiculoplexus neuropathy due to type 1 diabetes mellitus | FALSE |
| CCI score | 403438288 | 4174977 | Charlson - DMcx | TRUE | 1 | Retinopathy due to diabetes mellitus | FALSE |
| CCI score | 403438288 | 4222876 | Charlson - DMcx | TRUE | 1 | Gangrene due to type 2 diabetes mellitus | FALSE |
| CCI score | 403438288 | 4225656 | Charlson - DMcx | TRUE | 1 | Cataract due to diabetes mellitus type 1 | FALSE |
| CCI score | 403438288 | 37016180 | Charlson - DMcx | TRUE | 1 | Moderate nonproliferative retinopathy due to type 1 diabetes mellitus | FALSE |
| CCI score | 403438288 | 321822 | Charlson - DMcx | TRUE | 1 | Peripheral vascular disorder due to diabetes mellitus | FALSE |
| CCI score | 403438288 | 376065 | Charlson - DMcx | TRUE | 1 | Disorder of nervous system due to type 2 diabetes mellitus | FALSE |
| CCI score | 403438288 | 380096 | Charlson - DMcx | TRUE | 1 | Proliferative retinopathy due to diabetes mellitus | FALSE |
| CCI score | 403438288 | 380097 | Charlson - DMcx | TRUE | 1 | Macular edema due to diabetes mellitus | FALSE |
| CCI score | 403438288 | 4140466 | Charlson - DMcx | TRUE | 1 | Lumbosacral radiculoplexus neuropathy due to type 2 diabetes mellitus | FALSE |
| CCI score | 403438288 | 4175440 | Charlson - DMcx | TRUE | 1 | Autonomic neuropathy due to diabetes mellitus | FALSE |
| CCI score | 403438288 | 4191611 | Charlson - DMcx | TRUE | 1 | Lumbosacral radiculoplexus neuropathy due to diabetes mellitus | FALSE |
| CCI score | 403438288 | 4223303 | Charlson - DMcx | TRUE | 1 | Gangrene due to type 1 diabetes mellitus | FALSE |
| CCI score | 403438288 | 4225055 | Charlson - DMcx | TRUE | 1 | Mononeuropathy due to type 1 diabetes mellitus | FALSE |
| CCI score | 403438288 | 4227210 | Charlson - DMcx | TRUE | 1 | Retinopathy due to type 1 diabetes mellitus | FALSE |
| CCI score | 403438288 | 43530685 | Charlson - DMcx | TRUE | 1 | Proliferative retinopathy due to type 2 diabetes mellitus | FALSE |
| CCI score | 403438288 | 43531578 | Charlson - DMcx | TRUE | 1 | Chronic kidney disease due to type 2 diabetes mellitus | FALSE |
| CCI score | 403438288 | 376114 | Charlson - DMcx | TRUE | 1 | Severe nonproliferative retinopathy due to diabetes mellitus | FALSE |
| CCI score | 403438288 | 443731 | Charlson - DMcx | TRUE | 1 | Renal disorder due to type 2 diabetes mellitus | FALSE |
| CCI score | 403438288 | 4044391 | Charlson - DMcx | TRUE | 1 | Neuropathy due to diabetes mellitus | FALSE |
| CCI score | 403438288 | 43530656 | Charlson - DMcx | TRUE | 1 | Nonproliferative retinopathy due to type 2 diabetes mellitus | FALSE |
| CCI score | 403438288 | 377552 | Charlson - DMcx | TRUE | 1 | Moderate nonproliferative retinopathy due to diabetes mellitus | FALSE |
| CCI score | 403438288 | 443729 | Charlson - DMcx | TRUE | 1 | Peripheral circulatory disorder due to type 2 diabetes mellitus | FALSE |
| CCI score | 403438288 | 443730 | Charlson - DMcx | TRUE | 1 | Disorder of nervous system due to diabetes mellitus | FALSE |
| CCI score | 403438288 | 4131908 | Charlson - DMcx | TRUE | 1 | Peripheral angiopathy due to diabetes mellitus | FALSE |
| CCI score | 403438288 | 4338896 | Charlson - DMcx | TRUE | 1 | Traction retinal detachment involving macula | FALSE |
| CCI score | 403438288 | 45763584 | Charlson - DMcx | TRUE | 1 | Proliferative retinopathy due to type 1 diabetes mellitus | FALSE |
| CCI score | 403438288 | 376979 | Charlson - DMcx | TRUE | 1 | Cataract due to diabetes mellitus | FALSE |
| CCI score | 403438288 | 443733 | Charlson - DMcx | TRUE | 1 | Disorder of eye due to type 2 diabetes mellitus | FALSE |
| CCI score | 403438288 | 443767 | Charlson - DMcx | TRUE | 1 | Disorder of eye due to diabetes mellitus | FALSE |
| CCI score | 403438288 | 4338901 | Charlson - DMcx | TRUE | 1 | Traction detachment of retina due to diabetes mellitus | FALSE |
| CCI score | 403438288 | 45757435 | Charlson - DMcx | TRUE | 1 | Mild nonproliferative retinopathy due to type 2 diabetes mellitus | FALSE |
| CCI score | 403438288 | 45763583 | Charlson - DMcx | TRUE | 1 | Nonproliferative diabetic retinopathy due to type 1 diabetes mellitus | FALSE |
| CCI score | 403438288 | 45769873 | Charlson - DMcx | TRUE | 1 | Traction detachment of retina due to type 1 diabetes mellitus | FALSE |
| CCI score | 403438288 | 45773064 | Charlson - DMcx | TRUE | 1 | Traction detachment of retina due to type 2 diabetes mellitus | FALSE |
| CCI score | 403438288 | 376112 | Charlson - DMcx | TRUE | 1 | Polyneuropathy due to diabetes mellitus | FALSE |
| CCI score | 403438288 | 4048028 | Charlson - DMcx | TRUE | 1 | Diabetic mononeuropathy | FALSE |
| CCI score | 403438288 | 4222415 | Charlson - DMcx | TRUE | 1 | Mononeuropathy due to type 2 diabetes mellitus | FALSE |
| CCI score | 403438288 | 4290822 | Charlson - DMcx | TRUE | 1 | Severe nonproliferative retinopathy with clinically significant macular edema due to diabetes mellitus | FALSE |
| CCI score | 403438288 | 37016179 | Charlson - DMcx | TRUE | 1 | Mild nonproliferative retinopathy due to type 1 diabetes mellitus | FALSE |
| CCI score | 403438288 | 37016768 | Charlson - DMcx | TRUE | 1 | Autonomic neuropathy due to type 2 diabetes mellitus | FALSE |
| CCI score | 403438288 | 37017431 | Charlson - DMcx | TRUE | 1 | Polyneuropathy due to type 1 diabetes mellitus | FALSE |
| CCI score | 403438288 | 42538169 | Charlson - DMcx | TRUE | 1 | Disorder of eye due to type 1 diabetes mellitus | FALSE |
| CCI score | 403438288 | 45770881 | Charlson - DMcx | TRUE | 1 | Moderate nonproliferative retinopathy due to type 2 diabetes mellitus | FALSE |
| CCI score | 403438288 | 377821 | Charlson - DMcx | TRUE | 1 | Disorder of nervous system due to type 1 diabetes mellitus | FALSE |
| CCI score | 403438288 | 4221495 | Charlson - DMcx | TRUE | 1 | Cataract due to diabetes mellitus type 2 | FALSE |
| CCI score | 403438288 | 4226354 | Charlson - DMcx | TRUE | 1 | Gangrene due to diabetes mellitus | FALSE |
| CCI score | 403438288 | 4266637 | Charlson - DMcx | TRUE | 1 | Severe nonproliferative retinopathy without macular edema due to diabetes mellitus | FALSE |
| CCI score | 403438288 | 4338897 | Charlson - DMcx | TRUE | 1 | Combined traction and rhegmatogenous retinal detachment | FALSE |
| CCI score | 403438288 | 37016767 | Charlson - DMcx | TRUE | 1 | Autonomic neuropathy due to type 1 diabetes mellitus | FALSE |
| CCI score | 403438288 | 37017432 | Charlson - DMcx | TRUE | 1 | Polyneuropathy due to type 2 diabetes mellitus | FALSE |
| CCI score | 403438288 | 45770830 | Charlson - DMcx | TRUE | 1 | Macular edema and retinopathy due to type 2 diabetes mellitus | FALSE |
| CCI score | 494981955 | 4046123 | Charlson - LiverMild | TRUE | 1 | Secondary biliary cirrhosis | FALSE |
| CCI score | 494981955 | 4055225 | Charlson - LiverMild | TRUE | 1 | Toxic liver disease with chronic lobular hepatitis | FALSE |
| CCI score | 494981955 | 4059299 | Charlson - LiverMild | TRUE | 1 | Toxic liver disease with chronic active hepatitis | FALSE |
| CCI score | 494981955 | 192675 | Charlson - LiverMild | TRUE | 1 | Biliary cirrhosis | FALSE |
| CCI score | 494981955 | 194984 | Charlson - LiverMild | TRUE | 1 | Disease of liver | FALSE |
| CCI score | 494981955 | 196463 | Charlson - LiverMild | TRUE | 1 | Alcoholic cirrhosis | FALSE |
| CCI score | 494981955 | 4026125 | Charlson - LiverMild | TRUE | 1 | Chronic active hepatitis | FALSE |
| CCI score | 494981955 | 4340383 | Charlson - LiverMild | TRUE | 1 | Alcoholic hepatitis | FALSE |
| CCI score | 494981955 | 42537742 | Charlson - LiverMild | TRUE | 1 | Transplanted liver present | FALSE |
| CCI score | 494981955 | 192240 | Charlson - LiverMild | TRUE | 1 | Chronic viral hepatitis B with hepatitis D | FALSE |
| CCI score | 494981955 | 194417 | Charlson - LiverMild | TRUE | 1 | Hepatic infarction | FALSE |
| CCI score | 494981955 | 4238978 | Charlson - LiverMild | TRUE | 1 | Chronic lobular hepatitis | FALSE |
| CCI score | 494981955 | 46269816 | Charlson - LiverMild | TRUE | 1 | Ascites due to alcoholic cirrhosis | FALSE |
| CCI score | 494981955 | 46269835 | Charlson - LiverMild | TRUE | 1 | Hepatic ascites due to chronic alcoholic hepatitis | FALSE |
| CCI score | 494981955 | 193256 | Charlson - LiverMild | TRUE | 1 | Alcoholic fatty liver | FALSE |
| CCI score | 494981955 | 4059298 | Charlson - LiverMild | TRUE | 1 | Toxic liver disease with chronic persistent hepatitis | FALSE |
| CCI score | 494981955 | 4340385 | Charlson - LiverMild | TRUE | 1 | Alcoholic fibrosis and sclerosis of liver | FALSE |
| CCI score | 494981955 | 46273476 | Charlson - LiverMild | TRUE | 1 | Hepatic ascites co-occurrent with chronic active hepatitis due to toxic liver disease | FALSE |
| CCI score | 494981955 | 198964 | Charlson - LiverMild | TRUE | 1 | Chronic hepatitis C | FALSE |
| CCI score | 494981955 | 4012113 | Charlson - LiverMild | TRUE | 1 | Chronic viral hepatitis | FALSE |
| CCI score | 494981955 | 4135822 | Charlson - LiverMild | TRUE | 1 | Primary biliary cholangitis | FALSE |
| CCI score | 494981955 | 4267417 | Charlson - LiverMild | TRUE | 1 | Hepatic fibrosis | FALSE |
| CCI score | 494981955 | 201612 | Charlson - LiverMild | TRUE | 1 | Alcoholic liver damage | FALSE |
| CCI score | 494981955 | 4058695 | Charlson - LiverMild | TRUE | 1 | Toxic liver disease with fibrosis and cirrhosis of liver | FALSE |
| CCI score | 494981955 | 4058696 | Charlson - LiverMild | TRUE | 1 | Central hemorrhagic necrosis of liver | FALSE |
| CCI score | 494981955 | 4064161 | Charlson - LiverMild | TRUE | 1 | Cirrhosis of liver | FALSE |
| CCI score | 494981955 | 4240725 | Charlson - LiverMild | TRUE | 1 | Peliosis hepatis | FALSE |
| CCI score | 494981955 | 4340394 | Charlson - LiverMild | TRUE | 1 | Hepatic sclerosis | FALSE |
| CCI score | 494981955 | 4340948 | Charlson - LiverMild | TRUE | 1 | Hepatic fibrosis with hepatic sclerosis | FALSE |
| CCI score | 494981955 | 199867 | Charlson - LiverMild | TRUE | 1 | Chronic persistent hepatitis | FALSE |
| CCI score | 494981955 | 200763 | Charlson - LiverMild | TRUE | 1 | Chronic hepatitis | FALSE |
| CCI score | 494981955 | 439674 | Charlson - LiverMild | TRUE | 1 | Chronic viral hepatitis B without delta-agent | FALSE |
| CCI score | 494981955 | 4059290 | Charlson - LiverMild | TRUE | 1 | Steatosis of liver | FALSE |
| CCI score | 494981955 | 4159144 | Charlson - LiverMild | TRUE | 1 | Hepatopulmonary syndrome | FALSE |
| CCI score | 248333963 | 377604 | Charlson - LiverSevere | TRUE | 1 | Hepatic coma | FALSE |
| CCI score | 248333963 | 4277276 | Charlson - LiverSevere | TRUE | 1 | Veno-occlusive disease of the liver | FALSE |
| CCI score | 248333963 | 4340386 | Charlson - LiverSevere | TRUE | 1 | Alcoholic hepatic failure | FALSE |
| CCI score | 248333963 | 192680 | Charlson - LiverSevere | TRUE | 1 | Portal hypertension | FALSE |
| CCI score | 248333963 | 4340390 | Charlson - LiverSevere | TRUE | 1 | Chronic hepatic failure | FALSE |
| CCI score | 248333963 | 46269836 | Charlson - LiverSevere | TRUE | 1 | Hepatic coma due to chronic hepatic failure | FALSE |
| CCI score | 248333963 | 196455 | Charlson - LiverSevere | TRUE | 1 | Hepatorenal syndrome | FALSE |
| CCI score | 248333963 | 22340 | Charlson - LiverSevere | TRUE | 1 | Esophageal varices without bleeding | FALSE |
| CCI score | 248333963 | 24966 | Charlson - LiverSevere | TRUE | 1 | Esophageal varices | FALSE |
| CCI score | 248333963 | 4026136 | Charlson - LiverSevere | TRUE | 1 | Toxic liver disease with hepatic necrosis | FALSE |
| CCI score | 248333963 | 4245975 | Charlson - LiverSevere | TRUE | 1 | Hepatic failure | FALSE |
| CCI score | 248333963 | 46269818 | Charlson - LiverSevere | TRUE | 1 | Hepatic coma due to alcoholic liver failure | FALSE |
| CCI score | 248333963 | 28779 | Charlson - LiverSevere | TRUE | 1 | Bleeding esophageal varices | FALSE |
| CCI score | 248333963 | 4237824 | Charlson - LiverSevere | TRUE | 1 | Gastric varices | FALSE |
| CCI score | 378462283 | 72266 | Charlson - Mets | TRUE | 1 | Secondary malignant neoplasm of pleura | FALSE |
| CCI score | 378462283 | 136354 | Charlson - Mets | TRUE | 1 | Secondary malignant neoplasm of skin | FALSE |
| CCI score | 378462283 | 193144 | Charlson - Mets | TRUE | 1 | Secondary malignant neoplasm of adrenal gland | FALSE |
| CCI score | 378462283 | 198371 | Charlson - Mets | TRUE | 1 | Secondary malignant neoplasm of small intestine | FALSE |
| CCI score | 378462283 | 253717 | Charlson - Mets | TRUE | 1 | Secondary malignant neoplasm of respiratory tract | FALSE |
| CCI score | 378462283 | 432851 | Charlson - Mets | TRUE | 1 | Secondary malignant neoplastic disease | FALSE |
| CCI score | 378462283 | 442182 | Charlson - Mets | TRUE | 1 | Secondary malignant neoplasm of lymph nodes of lower limb | FALSE |
| CCI score | 378462283 | 4246450 | Charlson - Mets | TRUE | 1 | Secondary malignant neoplasm of bone marrow | FALSE |
| CCI score | 378462283 | 4281027 | Charlson - Mets | TRUE | 1 | Secondary malignant neoplasm of left ovary | FALSE |
| CCI score | 378462283 | 4314071 | Charlson - Mets | TRUE | 1 | Secondary malignant neoplasm of trunk | FALSE |
| CCI score | 378462283 | 44806773 | Charlson - Mets | TRUE | 1 | Secondary malignant neoplasm of liver and intrahepatic bile duct | FALSE |
| CCI score | 378462283 | 439751 | Charlson - Mets | TRUE | 1 | Secondary malignant neoplasm of intrathoracic lymph nodes | FALSE |
| CCI score | 378462283 | 4147162 | Charlson - Mets | TRUE | 1 | Secondary malignant neoplasm of respiratory and digestive systems | FALSE |
| CCI score | 378462283 | 4281030 | Charlson - Mets | TRUE | 1 | Secondary malignant neoplasm of right ovary | FALSE |
| CCI score | 378462283 | 46273652 | Charlson - Mets | TRUE | 1 | Secondary malignant neoplasm of left lung | FALSE |
| CCI score | 378462283 | 78987 | Charlson - Mets | TRUE | 1 | Secondary malignant neoplasm of urinary system | FALSE |
| CCI score | 378462283 | 318096 | Charlson - Mets | TRUE | 1 | Secondary malignant neoplasm of lymph node | FALSE |
| CCI score | 378462283 | 443252 | Charlson - Mets | TRUE | 1 | Post-transplant neoplasia | FALSE |
| CCI score | 378462283 | 4158910 | Charlson - Mets | TRUE | 1 | Secondary malignant neoplasm of unknown site | FALSE |
| CCI score | 378462283 | 4312802 | Charlson - Mets | TRUE | 1 | Secondary malignant neoplasm of bladder | FALSE |
| CCI score | 378462283 | 78097 | Charlson - Mets | TRUE | 1 | Secondary malignant neoplasm of bone | FALSE |
| CCI score | 378462283 | 196925 | Charlson - Mets | TRUE | 1 | Secondary malignant neoplasm of retroperitoneum and peritoneum | FALSE |
| CCI score | 378462283 | 434875 | Charlson - Mets | TRUE | 1 | Secondary malignant neoplasm of mediastinum | FALSE |
| CCI score | 378462283 | 4160276 | Charlson - Mets | TRUE | 1 | Malignant neoplasm of genital structure | FALSE |
| CCI score | 378462283 | 4247962 | Charlson - Mets | TRUE | 1 | Secondary malignant neoplasm of gastrointestinal tract | FALSE |
| CCI score | 378462283 | 4312290 | Charlson - Mets | TRUE | 1 | Secondary malignant neoplasm of cerebral meninges | FALSE |
| CCI score | 378462283 | 192568 | Charlson - Mets | TRUE | 1 | Secondary malignant neoplasm of intra-abdominal lymph nodes | FALSE |
| CCI score | 378462283 | 199752 | Charlson - Mets | TRUE | 1 | Secondary malignant neoplasm of ovary | FALSE |
| CCI score | 378462283 | 4246451 | Charlson - Mets | TRUE | 1 | Secondary malignant neoplasm of brain | FALSE |
| CCI score | 378462283 | 4315806 | Charlson - Mets | TRUE | 1 | Secondary malignant neoplasm of retroperitoneum | FALSE |
| CCI score | 378462283 | 200348 | Charlson - Mets | TRUE | 1 | Secondary malignant neoplasm of large intestine | FALSE |
| CCI score | 378462283 | 200959 | Charlson - Mets | TRUE | 1 | Secondary malignant neoplasm of intrapelvic lymph nodes | FALSE |
| CCI score | 378462283 | 373425 | Charlson - Mets | TRUE | 1 | Secondary malignant neoplasm of nervous system | FALSE |
| CCI score | 378462283 | 434298 | Charlson - Mets | TRUE | 1 | Secondary malignant neoplasm of lymph nodes of upper limb | FALSE |
| CCI score | 378462283 | 438701 | Charlson - Mets | TRUE | 1 | Disseminated malignancy of unknown primary | FALSE |
| CCI score | 378462283 | 46270513 | Charlson - Mets | TRUE | 1 | Secondary malignant neoplasm of right lung | FALSE |
| CCI score | 378462283 | 140960 | Charlson - Mets | TRUE | 1 | Secondary malignant neoplasm of female breast | FALSE |
| CCI score | 378462283 | 254591 | Charlson - Mets | TRUE | 1 | Secondary malignant neoplasm of lung | FALSE |
| CCI score | 378462283 | 320342 | Charlson - Mets | TRUE | 1 | Secondary malignant neoplasm of lymph nodes of multiple sites | FALSE |
| CCI score | 378462283 | 443392 | Charlson - Mets | TRUE | 1 | Malignant neoplastic disease | FALSE |
| CCI score | 259495957 | 4108218 | Charlson - MI | TRUE | 1 | Subsequent myocardial infarction of inferior wall | FALSE |
| CCI score | 259495957 | 43020460 | Charlson - MI | TRUE | 1 | Acute ST segment elevation myocardial infarction involving left anterior descending coronary artery | FALSE |
| CCI score | 259495957 | 314666 | Charlson - MI | TRUE | 1 | Old myocardial infarction | FALSE |
| CCI score | 259495957 | 4108217 | Charlson - MI | TRUE | 1 | Subsequent myocardial infarction | FALSE |
| CCI score | 259495957 | 45766114 | Charlson - MI | TRUE | 1 | Subsequent ST segment elevation myocardial infarction | FALSE |
| CCI score | 259495957 | 46270162 | Charlson - MI | TRUE | 1 | Acute ST segment elevation myocardial infarction due to left coronary artery occlusion | FALSE |
| CCI score | 259495957 | 312327 | Charlson - MI | TRUE | 1 | Acute myocardial infarction | FALSE |
| CCI score | 259495957 | 45766075 | Charlson - MI | TRUE | 1 | Acute anterior ST segment elevation myocardial infarction | FALSE |
| CCI score | 259495957 | 45766116 | Charlson - MI | TRUE | 1 | Acute ST segment elevation myocardial infarction of inferior wall | FALSE |
| CCI score | 259495957 | 45766241 | Charlson - MI | TRUE | 1 | Subsequent non-ST segment elevation myocardial infarction | FALSE |
| CCI score | 259495957 | 4108677 | Charlson - MI | TRUE | 1 | Subsequent myocardial infarction of anterior wall | FALSE |
| CCI score | 259495957 | 4329847 | Charlson - MI | TRUE | 1 | Myocardial infarction | FALSE |
| CCI score | 259495957 | 4296653 | Charlson - MI | TRUE | 1 | Acute ST segment elevation myocardial infarction | FALSE |
| CCI score | 259495957 | 46270163 | Charlson - MI | TRUE | 1 | Acute ST segment elevation myocardial infarction due to right coronary artery occlusion | FALSE |
| CCI score | 259495957 | 4270024 | Charlson - MI | TRUE | 1 | Acute non-ST segment elevation myocardial infarction | FALSE |
| CCI score | 489555336 | 372613 | Charlson - Paralysis | TRUE | 1 | Flaccid hemiplegia | FALSE |
| CCI score | 489555336 | 375528 | Charlson - Paralysis | TRUE | 1 | Spastic hemiplegia | FALSE |
| CCI score | 489555336 | 4102342 | Charlson - Paralysis | TRUE | 1 | Cauda equina syndrome | FALSE |
| CCI score | 489555336 | 40480435 | Charlson - Paralysis | TRUE | 1 | Spastic hemiplegia of nondominant side | FALSE |
| CCI score | 489555336 | 43530719 | Charlson - Paralysis | TRUE | 1 | Incomplete paraplegia | FALSE |
| CCI score | 489555336 | 195240 | Charlson - Paralysis | TRUE | 1 | Monoplegia of lower limb | FALSE |
| CCI score | 489555336 | 40480416 | Charlson - Paralysis | TRUE | 1 | Complete tetraplegia due to lesion at C1-C4 level | FALSE |
| CCI score | 489555336 | 43530718 | Charlson - Paralysis | TRUE | 1 | Complete paraplegia | FALSE |
| CCI score | 489555336 | 372880 | Charlson - Paralysis | TRUE | 1 | Diplegia of upper limbs | FALSE |
| CCI score | 489555336 | 374377 | Charlson - Paralysis | TRUE | 1 | Paralytic syndrome | FALSE |
| CCI score | 489555336 | 380393 | Charlson - Paralysis | TRUE | 1 | Monoplegia of upper limb affecting non-dominant side | FALSE |
| CCI score | 489555336 | 381548 | Charlson - Paralysis | TRUE | 1 | Monoplegia | FALSE |
| CCI score | 489555336 | 4144328 | Charlson - Paralysis | TRUE | 1 | Monoplegia of lower limb affecting dominant side | FALSE |
| CCI score | 489555336 | 37019108 | Charlson - Paralysis | TRUE | 1 | Human T-cell lymphotropic virus 1-associated myelopathy | FALSE |
| CCI score | 489555336 | 40480429 | Charlson - Paralysis | TRUE | 1 | Hemiplegia of nondominant side | FALSE |
| CCI score | 489555336 | 40480514 | Charlson - Paralysis | TRUE | 1 | Incomplete tetraplegia due to lesion at C5-C7 level | FALSE |
| CCI score | 489555336 | 40481345 | Charlson - Paralysis | TRUE | 1 | Monoplegia of upper limb of dominant side | FALSE |
| CCI score | 489555336 | 40481757 | Charlson - Paralysis | TRUE | 1 | Flaccid hemiplegia of dominant side | FALSE |
| CCI score | 489555336 | 44806793 | Charlson - Paralysis | TRUE | 1 | Spastic hemiplegic cerebral palsy | FALSE |
| CCI score | 489555336 | 379012 | Charlson - Paralysis | TRUE | 1 | Monoplegia of upper limb | FALSE |
| CCI score | 489555336 | 40480944 | Charlson - Paralysis | TRUE | 1 | Spastic hemiplegia of dominant side | FALSE |
| CCI score | 489555336 | 40481376 | Charlson - Paralysis | TRUE | 1 | Complete tetraplegia due to lesion at C5-C7 level | FALSE |
| CCI score | 489555336 | 40481820 | Charlson - Paralysis | TRUE | 1 | Flaccid hemiplegia of nondominant side | FALSE |
| CCI score | 489555336 | 40482237 | Charlson - Paralysis | TRUE | 1 | Hemiplegia of dominant side | FALSE |
| CCI score | 489555336 | 192606 | Charlson - Paralysis | TRUE | 1 | Paraplegia | FALSE |
| CCI score | 489555336 | 374914 | Charlson - Paralysis | TRUE | 1 | Tetraplegia | FALSE |
| CCI score | 489555336 | 4141654 | Charlson - Paralysis | TRUE | 1 | Monoplegia of lower limb affecting non-dominant side | FALSE |
| CCI score | 489555336 | 132617 | Charlson - Paralysis | TRUE | 1 | Diplegic cerebral palsy | FALSE |
| CCI score | 489555336 | 374022 | Charlson - Paralysis | TRUE | 1 | Hemiplegia | FALSE |
| CCI score | 489555336 | 43531639 | Charlson - Paralysis | TRUE | 1 | Paralytic syndrome on one side of the body | FALSE |
| CCI score | 489555336 | 192901 | Charlson - Paralysis | TRUE | 1 | Hereditary spastic paraplegia | FALSE |
| CCI score | 489555336 | 40480066 | Charlson - Paralysis | TRUE | 1 | Incomplete tetraplegia due to spinal cord lesion at C1-C4 level | FALSE |
| CCI score | 510748896 | 4222896 | Charlson - PUD | TRUE | 1 | Chronic duodenal ulcer without hemorrhage AND without perforation | FALSE |
| CCI score | 510748896 | 4231580 | Charlson - PUD | TRUE | 1 | Acute gastric ulcer with hemorrhage | FALSE |
| CCI score | 510748896 | 433515 | Charlson - PUD | TRUE | 1 | Chronic gastrojejunal ulcer with hemorrhage | FALSE |
| CCI score | 510748896 | 4173408 | Charlson - PUD | TRUE | 1 | Chronic duodenal ulcer with perforation | FALSE |
| CCI score | 510748896 | 4177387 | Charlson - PUD | TRUE | 1 | Chronic gastrojejunal ulcer without hemorrhage AND without perforation | FALSE |
| CCI score | 510748896 | 4198381 | Charlson - PUD | TRUE | 1 | Ulcer of duodenum | FALSE |
| CCI score | 510748896 | 4265479 | Charlson - PUD | TRUE | 1 | Acute duodenal ulcer with perforation | FALSE |
| CCI score | 510748896 | 4289830 | Charlson - PUD | TRUE | 1 | Chronic duodenal ulcer with hemorrhage AND perforation | FALSE |
| CCI score | 510748896 | 4336230 | Charlson - PUD | TRUE | 1 | Acute duodenal ulcer with hemorrhage AND perforation | FALSE |
| CCI score | 510748896 | 4101870 | Charlson - PUD | TRUE | 1 | Chronic gastrojejunal ulcer with perforation | FALSE |
| CCI score | 510748896 | 4146517 | Charlson - PUD | TRUE | 1 | Chronic peptic ulcer with perforation | FALSE |
| CCI score | 510748896 | 4147683 | Charlson - PUD | TRUE | 1 | Acute gastrojejunal ulcer without hemorrhage AND without perforation | FALSE |
| CCI score | 510748896 | 4174044 | Charlson - PUD | TRUE | 1 | Chronic peptic ulcer with hemorrhage | FALSE |
| CCI score | 510748896 | 4247008 | Charlson - PUD | TRUE | 1 | Chronic peptic ulcer with hemorrhage AND perforation | FALSE |
| CCI score | 510748896 | 4248429 | Charlson - PUD | TRUE | 1 | Gastric ulcer without hemorrhage AND without perforation | FALSE |
| CCI score | 510748896 | 4265600 | Charlson - PUD | TRUE | 1 | Gastric ulcer | FALSE |
| CCI score | 510748896 | 4296611 | Charlson - PUD | TRUE | 1 | Chronic gastric ulcer without hemorrhage AND without perforation | FALSE |
| CCI score | 510748896 | 4027729 | Charlson - PUD | TRUE | 1 | Acute duodenal ulcer with hemorrhage | FALSE |
| CCI score | 510748896 | 4057953 | Charlson - PUD | TRUE | 1 | Acute gastric ulcer with perforation | FALSE |
| CCI score | 510748896 | 4059178 | Charlson - PUD | TRUE | 1 | Gastrojejunal ulcer | FALSE |
| CCI score | 510748896 | 4164920 | Charlson - PUD | TRUE | 1 | Chronic gastrojejunal ulcer with hemorrhage and perforation | FALSE |
| CCI score | 510748896 | 4274491 | Charlson - PUD | TRUE | 1 | Acute gastrojejunal ulcer with hemorrhage | FALSE |
| CCI score | 510748896 | 4294973 | Charlson - PUD | TRUE | 1 | Chronic gastric ulcer with hemorrhage and with perforation | FALSE |
| CCI score | 510748896 | 4027663 | Charlson - PUD | TRUE | 1 | Peptic ulcer | FALSE |
| CCI score | 510748896 | 4046500 | Charlson - PUD | TRUE | 1 | Acute peptic ulcer with hemorrhage | FALSE |
| CCI score | 510748896 | 4150681 | Charlson - PUD | TRUE | 1 | Chronic gastric ulcer with perforation | FALSE |
| CCI score | 510748896 | 4169592 | Charlson - PUD | TRUE | 1 | Acute gastric ulcer with hemorrhage and perforation | FALSE |
| CCI score | 510748896 | 4204555 | Charlson - PUD | TRUE | 1 | Chronic peptic ulcer without hemorrhage AND without perforation | FALSE |
| CCI score | 510748896 | 4280942 | Charlson - PUD | TRUE | 1 | Acute gastrojejunal ulcer with perforation | FALSE |
| CCI score | 510748896 | 4006994 | Charlson - PUD | TRUE | 1 | Acute peptic ulcer with hemorrhage and perforation | FALSE |
| CCI score | 510748896 | 4101104 | Charlson - PUD | TRUE | 1 | Gastrojejunal ulcer without hemorrhage AND without perforation | FALSE |
| CCI score | 510748896 | 4194543 | Charlson - PUD | TRUE | 1 | Acute peptic ulcer with perforation | FALSE |
| CCI score | 510748896 | 4195231 | Charlson - PUD | TRUE | 1 | Acute gastric ulcer without hemorrhage AND without perforation | FALSE |
| CCI score | 510748896 | 4211001 | Charlson - PUD | TRUE | 1 | Chronic gastric ulcer with hemorrhage | FALSE |
| CCI score | 510748896 | 4217947 | Charlson - PUD | TRUE | 1 | Acute gastrojejunal ulcer with hemorrhage and perforation | FALSE |
| CCI score | 510748896 | 4138962 | Charlson - PUD | TRUE | 1 | Acute duodenal ulcer without hemorrhage AND without perforation | FALSE |
| CCI score | 510748896 | 4163865 | Charlson - PUD | TRUE | 1 | Acute peptic ulcer without hemorrhage AND without perforation | FALSE |
| CCI score | 510748896 | 4232181 | Charlson - PUD | TRUE | 1 | Chronic duodenal ulcer with hemorrhage | FALSE |
| CCI score | 514953976 | 435298 | Charlson - Pulmonary | TRUE | 1 | Farmers' lung | FALSE |
| CCI score | 514953976 | 4112814 | Charlson - Pulmonary | TRUE | 1 | Chronic drug-induced interstitial lung disorders | FALSE |
| CCI score | 514953976 | 4145356 | Charlson - Pulmonary | TRUE | 1 | Severe persistent asthma | FALSE |
| CCI score | 514953976 | 4167085 | Charlson - Pulmonary | TRUE | 1 | Pulmonary heart disease | FALSE |
| CCI score | 514953976 | 4249010 | Charlson - Pulmonary | TRUE | 1 | Pneumoconiosis due to talc | FALSE |
| CCI score | 514953976 | 45769351 | Charlson - Pulmonary | TRUE | 1 | Acute severe exacerbation of moderate persistent asthma | FALSE |
| CCI score | 514953976 | 255841 | Charlson - Pulmonary | TRUE | 1 | Chronic bronchitis | FALSE |
| CCI score | 514953976 | 261889 | Charlson - Pulmonary | TRUE | 1 | Simple chronic bronchitis | FALSE |
| CCI score | 514953976 | 320136 | Charlson - Pulmonary | TRUE | 1 | Disorder of respiratory system | FALSE |
| CCI score | 514953976 | 434975 | Charlson - Pulmonary | TRUE | 1 | Malt-workers' lung | FALSE |
| CCI score | 514953976 | 439853 | Charlson - Pulmonary | TRUE | 1 | Bird-fanciers' lung | FALSE |
| CCI score | 514953976 | 442125 | Charlson - Pulmonary | TRUE | 1 | Pneumoconiosis due to silica | FALSE |
| CCI score | 514953976 | 4066407 | Charlson - Pulmonary | TRUE | 1 | Graphite fibrosis of lung | FALSE |
| CCI score | 514953976 | 4112673 | Charlson - Pulmonary | TRUE | 1 | Air-conditioner and humidifier lung | FALSE |
| CCI score | 514953976 | 4142738 | Charlson - Pulmonary | TRUE | 1 | Moderate persistent asthma | FALSE |
| CCI score | 514953976 | 4143828 | Charlson - Pulmonary | TRUE | 1 | Mild persistent asthma | FALSE |
| CCI score | 514953976 | 4302900 | Charlson - Pulmonary | TRUE | 1 | Cannabinosis | FALSE |
| CCI score | 514953976 | 45769438 | Charlson - Pulmonary | TRUE | 1 | Acute severe exacerbation of asthma | FALSE |
| CCI score | 514953976 | 252348 | Charlson - Pulmonary | TRUE | 1 | Chronic pulmonary radiation disease | FALSE |
| CCI score | 514953976 | 256146 | Charlson - Pulmonary | TRUE | 1 | Pneumoconiosis due to silicate | FALSE |
| CCI score | 514953976 | 313236 | Charlson - Pulmonary | TRUE | 1 | Cough variant asthma | FALSE |
| CCI score | 514953976 | 316452 | Charlson - Pulmonary | TRUE | 1 | Chronic respiratory condition due to fumes AND/OR vapors | FALSE |
| CCI score | 514953976 | 4032314 | Charlson - Pulmonary | TRUE | 1 | Bauxite fibrosis of lung | FALSE |
| CCI score | 514953976 | 4286497 | Charlson - Pulmonary | TRUE | 1 | Centriacinar emphysema | FALSE |
| CCI score | 514953976 | 4311814 | Charlson - Pulmonary | TRUE | 1 | Byssinosis | FALSE |
| CCI score | 514953976 | 40493243 | Charlson - Pulmonary | TRUE | 1 | Eisenmenger's syndrome | FALSE |
| CCI score | 514953976 | 45768908 | Charlson - Pulmonary | TRUE | 1 | Exercise induced bronchospasm | FALSE |
| CCI score | 514953976 | 45768963 | Charlson - Pulmonary | TRUE | 1 | Uncomplicated mild persistent asthma | FALSE |
| CCI score | 514953976 | 45768965 | Charlson - Pulmonary | TRUE | 1 | Uncomplicated severe persistent asthma | FALSE |
| CCI score | 514953976 | 45771045 | Charlson - Pulmonary | TRUE | 1 | Acute exacerbation of asthma | FALSE |
| CCI score | 514953976 | 46273487 | Charlson - Pulmonary | TRUE | 1 | Acute exacerbation of moderate persistent asthma | FALSE |
| CCI score | 514953976 | 254389 | Charlson - Pulmonary | TRUE | 1 | Pneumoconiosis due to inorganic dust | FALSE |
| CCI score | 514953976 | 256449 | Charlson - Pulmonary | TRUE | 1 | Bronchiectasis | FALSE |
| CCI score | 514953976 | 256450 | Charlson - Pulmonary | TRUE | 1 | Asbestosis | FALSE |
| CCI score | 514953976 | 257004 | Charlson - Pulmonary | TRUE | 1 | Acute exacerbation of chronic obstructive airways disease | FALSE |
| CCI score | 514953976 | 438175 | Charlson - Pulmonary | TRUE | 1 | Maple-bark strippers' lung | FALSE |
| CCI score | 514953976 | 4112676 | Charlson - Pulmonary | TRUE | 1 | Pneumoconiosis associated with tuberculosis | FALSE |
| CCI score | 514953976 | 4138760 | Charlson - Pulmonary | TRUE | 1 | Exacerbation of intermittent asthma | FALSE |
| CCI score | 514953976 | 4163244 | Charlson - Pulmonary | TRUE | 1 | Unilateral emphysema | FALSE |
| CCI score | 514953976 | 4177944 | Charlson - Pulmonary | TRUE | 1 | Panacinar emphysema | FALSE |
| CCI score | 514953976 | 4236624 | Charlson - Pulmonary | TRUE | 1 | Aluminosis of lung | FALSE |
| CCI score | 514953976 | 44782732 | Charlson - Pulmonary | TRUE | 1 | Chronic pulmonary embolism | FALSE |
| CCI score | 514953976 | 45769350 | Charlson - Pulmonary | TRUE | 1 | Acute severe exacerbation of severe persistent asthma | FALSE |
| CCI score | 514953976 | 256451 | Charlson - Pulmonary | TRUE | 1 | Bronchitis | FALSE |
| CCI score | 514953976 | 261325 | Charlson - Pulmonary | TRUE | 1 | Pulmonary emphysema | FALSE |
| CCI score | 514953976 | 443890 | Charlson - Pulmonary | TRUE | 1 | Suberosis | FALSE |
| CCI score | 514953976 | 4110056 | Charlson - Pulmonary | TRUE | 1 | Chronic obstructive pulmonary disease with acute lower respiratory infection | FALSE |
| CCI score | 514953976 | 4112826 | Charlson - Pulmonary | TRUE | 1 | Mixed simple and mucopurulent chronic bronchitis | FALSE |
| CCI score | 514953976 | 4146581 | Charlson - Pulmonary | TRUE | 1 | Mild intermittent asthma | FALSE |
| CCI score | 514953976 | 4266525 | Charlson - Pulmonary | TRUE | 1 | Pulmonary siderosis | FALSE |
| CCI score | 514953976 | 45768964 | Charlson - Pulmonary | TRUE | 1 | Uncomplicated moderate persistent asthma | FALSE |
| CCI score | 514953976 | 45769352 | Charlson - Pulmonary | TRUE | 1 | Acute severe exacerbation of mild persistent asthma | FALSE |
| CCI score | 514953976 | 255573 | Charlson - Pulmonary | TRUE | 1 | Chronic obstructive lung disease | FALSE |
| CCI score | 514953976 | 317009 | Charlson - Pulmonary | TRUE | 1 | Asthma | FALSE |
| CCI score | 514953976 | 433233 | Charlson - Pulmonary | TRUE | 1 | Mushroom workers' lung | FALSE |
| CCI score | 514953976 | 444084 | Charlson - Pulmonary | TRUE | 1 | Extrinsic allergic alveolitis | FALSE |
| CCI score | 514953976 | 4027669 | Charlson - Pulmonary | TRUE | 1 | Flax-dressers' disease | FALSE |
| CCI score | 514953976 | 4195892 | Charlson - Pulmonary | TRUE | 1 | Chronic cor pulmonale | FALSE |
| CCI score | 514953976 | 4221139 | Charlson - Pulmonary | TRUE | 1 | Berylliosis | FALSE |
| CCI score | 514953976 | 46270082 | Charlson - Pulmonary | TRUE | 1 | Acute exacerbation of mild persistent asthma | FALSE |
| CCI score | 514953976 | 252946 | Charlson - Pulmonary | TRUE | 1 | Coal workers' pneumoconiosis | FALSE |
| CCI score | 514953976 | 257905 | Charlson - Pulmonary | TRUE | 1 | Mucopurulent chronic bronchitis | FALSE |
| CCI score | 514953976 | 259044 | Charlson - Pulmonary | TRUE | 1 | Pneumoconiosis | FALSE |
| CCI score | 514953976 | 437588 | Charlson - Pulmonary | TRUE | 1 | Bagassosis | FALSE |
| CCI score | 514953976 | 4196950 | Charlson - Pulmonary | TRUE | 1 | Stannosis | FALSE |
| CCI score | 514953976 | 37116845 | Charlson - Pulmonary | TRUE | 1 | Acute severe refractory exacerbation of asthma | FALSE |
| CCI score | 514953976 | 40483342 | Charlson - Pulmonary | TRUE | 1 | Acute exacerbation of bronchiectasis | FALSE |
| CCI score | 514953976 | 45768910 | Charlson - Pulmonary | TRUE | 1 | Uncomplicated asthma | FALSE |
| CCI score | 376881697 | 199064 | Charlson - PVD | TRUE | 1 | Chronic vascular insufficiency of intestine | FALSE |
| CCI score | 376881697 | 312343 | Charlson - PVD | TRUE | 1 | Ruptured aortic aneurysm | FALSE |
| CCI score | 376881697 | 317305 | Charlson - PVD | TRUE | 1 | Stricture of artery | FALSE |
| CCI score | 376881697 | 317585 | Charlson - PVD | TRUE | 1 | Aortic aneurysm | FALSE |
| CCI score | 376881697 | 441875 | Charlson - PVD | TRUE | 1 | Thoracic aortic aneurysm which has ruptured | FALSE |
| CCI score | 376881697 | 443358 | Charlson - PVD | TRUE | 1 | Ulcer of heel | FALSE |
| CCI score | 376881697 | 4177703 | Charlson - PVD | TRUE | 1 | Ulcer | FALSE |
| CCI score | 376881697 | 36717279 | Charlson - PVD | TRUE | 1 | Gangrene of right lower limb due to atherosclerosis | FALSE |
| CCI score | 376881697 | 37110250 | Charlson - PVD | TRUE | 1 | Atherosclerosis of artery of lower limb | FALSE |
| CCI score | 376881697 | 40479625 | Charlson - PVD | TRUE | 1 | Atherosclerosis of artery | FALSE |
| CCI score | 376881697 | 46271459 | Charlson - PVD | TRUE | 1 | Atherosclerosis of bypass graft of lower limb | FALSE |
| CCI score | 376881697 | 195834 | Charlson - PVD | TRUE | 1 | Atherosclerosis of renal artery | FALSE |
| CCI score | 376881697 | 312939 | Charlson - PVD | TRUE | 1 | Thromboangiitis obliterans | FALSE |
| CCI score | 376881697 | 433222 | Charlson - PVD | TRUE | 1 | Thoracoabdominal aortic aneurysm, ruptured | FALSE |
| CCI score | 376881697 | 4325344 | Charlson - PVD | TRUE | 1 | Pain at rest due to peripheral vascular disease | FALSE |
| CCI score | 376881697 | 36712963 | Charlson - PVD | TRUE | 1 | Gangrene of left lower limb due to atherosclerosis | FALSE |
| CCI score | 376881697 | 37016882 | Charlson - PVD | TRUE | 1 | Dissection of thoracoabdominal aorta | FALSE |
| CCI score | 376881697 | 44782819 | Charlson - PVD | TRUE | 1 | Chronic occlusion of artery of extremity | FALSE |
| CCI score | 376881697 | 138525 | Charlson - PVD | TRUE | 1 | Pain in limb | FALSE |
| CCI score | 376881697 | 197304 | Charlson - PVD | TRUE | 1 | Ulcer of lower extremity | FALSE |
| CCI score | 376881697 | 315558 | Charlson - PVD | TRUE | 1 | Atherosclerosis of arteries of the extremities | FALSE |
| CCI score | 376881697 | 321052 | Charlson - PVD | TRUE | 1 | Peripheral vascular disease | FALSE |
| CCI score | 376881697 | 442287 | Charlson - PVD | TRUE | 1 | Rest pain | FALSE |
| CCI score | 376881697 | 4134603 | Charlson - PVD | TRUE | 1 | Vascular disorder of intestine | FALSE |
| CCI score | 376881697 | 4316222 | Charlson - PVD | TRUE | 1 | Venous intermittent claudication | FALSE |
| CCI score | 376881697 | 36717006 | Charlson - PVD | TRUE | 1 | Intermittent claudication of bilateral lower limbs co-occurrent and due to atherosclerosis | FALSE |
| CCI score | 376881697 | 36717286 | Charlson - PVD | TRUE | 1 | Intermittent claudication of left lower limb co-occurrent and due to atherosclerosis | FALSE |
| CCI score | 376881697 | 37312529 | Charlson - PVD | TRUE | 1 | Intermittent claudication due to atherosclerosis of artery of limb | FALSE |
| CCI score | 376881697 | 37312531 | Charlson - PVD | TRUE | 1 | Gangrene of limb due to atherosclerosis of artery of limb | FALSE |
| CCI score | 376881697 | 40484541 | Charlson - PVD | TRUE | 1 | Atherosclerosis of autologous vein bypass graft of limb | FALSE |
| CCI score | 376881697 | 74719 | Charlson - PVD | TRUE | 1 | Ulcer of foot | FALSE |
| CCI score | 376881697 | 134380 | Charlson - PVD | TRUE | 1 | Erythromelalgia | FALSE |
| CCI score | 376881697 | 317577 | Charlson - PVD | TRUE | 1 | Arteriosclerotic gangrene | FALSE |
| CCI score | 376881697 | 321882 | Charlson - PVD | TRUE | 1 | Generalized atherosclerosis | FALSE |
| CCI score | 376881697 | 443593 | Charlson - PVD | TRUE | 1 | Ulcer of thigh | FALSE |
| CCI score | 376881697 | 4323360 | Charlson - PVD | TRUE | 1 | History of cardiovascular surgery | FALSE |
| CCI score | 376881697 | 35615028 | Charlson - PVD | TRUE | 1 | Bilateral atherosclerosis of lower limbs with gangrene | FALSE |
| CCI score | 376881697 | 36712806 | Charlson - PVD | TRUE | 1 | Intermittent claudication of right lower limb co-occurrent and due to atherosclerosis | FALSE |
| CCI score | 376881697 | 36712807 | Charlson - PVD | TRUE | 1 | Pain at rest of right lower limb co-occurrent and due to atherosclerosis | FALSE |
| CCI score | 376881697 | 37312520 | Charlson - PVD | TRUE | 1 | Ischemic foot ulcer due to atherosclerosis of artery of lower limb | FALSE |
| CCI score | 376881697 | 40484551 | Charlson - PVD | TRUE | 1 | Atherosclerosis of nonautologous biological bypass graft of limb | FALSE |
| CCI score | 376881697 | 195559 | Charlson - PVD | TRUE | 1 | Ruptured abdominal aortic aneurysm | FALSE |
| CCI score | 376881697 | 318443 | Charlson - PVD | TRUE | 1 | Arteriosclerotic vascular disease | FALSE |
| CCI score | 376881697 | 321314 | Charlson - PVD | TRUE | 1 | Abdominal aortic aneurysm without rupture | FALSE |
| CCI score | 376881697 | 436136 | Charlson - PVD | TRUE | 1 | Dissection of thoracic aorta | FALSE |
| CCI score | 376881697 | 4215685 | Charlson - PVD | TRUE | 1 | Past history of procedure | FALSE |
| CCI score | 376881697 | 4256889 | Charlson - PVD | TRUE | 1 | Dissection of abdominal aorta | FALSE |
| CCI score | 376881697 | 37312519 | Charlson - PVD | TRUE | 1 | Ulcer of calf due to atherosclerosis of artery of lower limb | FALSE |
| CCI score | 376881697 | 40483538 | Charlson - PVD | TRUE | 1 | Atherosclerosis of bypass graft of limb | FALSE |
| CCI score | 376881697 | 312934 | Charlson - PVD | TRUE | 1 | Atherosclerosis of aorta | FALSE |
| CCI score | 376881697 | 320739 | Charlson - PVD | TRUE | 1 | Dissection of aorta | FALSE |
| CCI score | 376881697 | 441051 | Charlson - PVD | TRUE | 1 | Thoracic aortic aneurysm without rupture | FALSE |
| CCI score | 376881697 | 4171556 | Charlson - PVD | TRUE | 1 | Ankle ulcer | FALSE |
| CCI score | 376881697 | 4187240 | Charlson - PVD | TRUE | 1 | Disorder of aorta | FALSE |
| CCI score | 376881697 | 35611566 | Charlson - PVD | TRUE | 1 | Bilateral lower limb atherosclerosis pain at rest co-occurrent and due to atherosclerosis | FALSE |
| CCI score | 376881697 | 37312524 | Charlson - PVD | TRUE | 1 | Ulcer of ankle due to atherosclerosis of artery of lower limb | FALSE |
| CCI score | 376881697 | 436996 | Charlson - PVD | TRUE | 1 | Thoracoabdominal aortic aneurysm | FALSE |
| CCI score | 376881697 | 442774 | Charlson - PVD | TRUE | 1 | Intermittent claudication | FALSE |
| CCI score | 376881697 | 36712805 | Charlson - PVD | TRUE | 1 | Pain at rest of left lower limb co-occurrent and due to atherosclerosis | FALSE |
| CCI score | 220495690 | 4019967 | Charlson - Renal | TRUE | 1 | Dependence on renal dialysis | FALSE |
| CCI score | 220495690 | 42539502 | Charlson - Renal | TRUE | 1 | Transplanted kidney present | FALSE |
| CCI score | 220495690 | 44784621 | Charlson - Renal | TRUE | 1 | Hypertensive heart and chronic kidney disease | FALSE |
| CCI score | 220495690 | 192359 | Charlson - Renal | TRUE | 1 | Renal failure syndrome | FALSE |
| CCI score | 220495690 | 197921 | Charlson - Renal | TRUE | 1 | Renal osteodystrophy | FALSE |
| CCI score | 220495690 | 443601 | Charlson - Renal | TRUE | 1 | Chronic kidney disease stage 2 | FALSE |
| CCI score | 220495690 | 443614 | Charlson - Renal | TRUE | 1 | Chronic kidney disease stage 1 | FALSE |
| CCI score | 220495690 | 4056480 | Charlson - Renal | TRUE | 1 | Chronic nephritic syndrome, diffuse crescentic glomerulonephritis | FALSE |
| CCI score | 220495690 | 4146996 | Charlson - Renal | TRUE | 1 | Mesangial proliferative glomerulonephritis | FALSE |
| CCI score | 220495690 | 4298809 | Charlson - Renal | TRUE | 1 | Nephritic syndrome | FALSE |
| CCI score | 220495690 | 252365 | Charlson - Renal | TRUE | 1 | Membranous glomerulonephritis | FALSE |
| CCI score | 220495690 | 443597 | Charlson - Renal | TRUE | 1 | Chronic kidney disease stage 3 | FALSE |
| CCI score | 220495690 | 443611 | Charlson - Renal | TRUE | 1 | Chronic kidney disease stage 5 | FALSE |
| CCI score | 220495690 | 4056462 | Charlson - Renal | TRUE | 1 | Chronic mesangial proliferative glomerulonephritis | FALSE |
| CCI score | 220495690 | 4059463 | Charlson - Renal | TRUE | 1 | Chronic nephritic syndrome, diffuse endocapillary proliferative glomerulonephritis | FALSE |
| CCI score | 220495690 | 4284491 | Charlson - Renal | TRUE | 1 | Diffuse endocapillary proliferative glomerulonephritis | FALSE |
| CCI score | 220495690 | 443612 | Charlson - Renal | TRUE | 1 | Chronic kidney disease stage 4 | FALSE |
| CCI score | 220495690 | 4055899 | Charlson - Renal | TRUE | 1 | Chronic nephritic syndrome, dense deposit disease | FALSE |
| CCI score | 220495690 | 4059584 | Charlson - Renal | TRUE | 1 | Chronic nephritic syndrome, diffuse mesangiocapillary glomerulonephritis | FALSE |
| CCI score | 220495690 | 4272486 | Charlson - Renal | TRUE | 1 | Diffuse crescentic glomerulonephritis | FALSE |
| CCI score | 220495690 | 4301680 | Charlson - Renal | TRUE | 1 | Dialysis care | FALSE |
| CCI score | 220495690 | 36717583 | Charlson - Renal | TRUE | 1 | Dense deposit disease | FALSE |
| CCI score | 220495690 | 443919 | Charlson - Renal | TRUE | 1 | Hypertensive renal failure | FALSE |
| CCI score | 220495690 | 4203722 | Charlson - Renal | TRUE | 1 | Patient encounter procedure | FALSE |
| CCI score | 220495690 | 46270347 | Charlson - Renal | TRUE | 1 | Chronic nephritic syndrome with membranous glomerulonephritis | FALSE |
| CCI score | 220495690 | 193782 | Charlson - Renal | TRUE | 1 | End-stage renal disease | FALSE |
| CCI score | 220495690 | 433257 | Charlson - Renal | TRUE | 1 | Mesangiocapillary glomerulonephritis | FALSE |
| CCI score | 220495690 | 46271022 | Charlson - Renal | TRUE | 1 | Chronic kidney disease | FALSE |
| CCI score | 765004404 | 134453 | Charlson - Rheumatic | TRUE | 1 | Bursitis | FALSE |
| CCI score | 765004404 | 255348 | Charlson - Rheumatic | TRUE | 1 | Polymyalgia rheumatica | FALSE |
| CCI score | 765004404 | 4055369 | Charlson - Rheumatic | TRUE | 1 | Lung disease with polymyositis | FALSE |
| CCI score | 765004404 | 4055640 | Charlson - Rheumatic | TRUE | 1 | Lung disease with systemic lupus erythematosus | FALSE |
| CCI score | 765004404 | 4081250 | Charlson - Rheumatic | TRUE | 1 | Dermatomyositis sine myositis | FALSE |
| CCI score | 765004404 | 4271003 | Charlson - Rheumatic | TRUE | 1 | Rheumatoid vasculitis | FALSE |
| CCI score | 765004404 | 4344166 | Charlson - Rheumatic | TRUE | 1 | Adult onset Still's disease | FALSE |
| CCI score | 765004404 | 42534834 | Charlson - Rheumatic | TRUE | 1 | Rheumatoid arthritis of left hand | FALSE |
| CCI score | 765004404 | 42534837 | Charlson - Rheumatic | TRUE | 1 | Rheumatoid arthritis of right hip | FALSE |
| CCI score | 765004404 | 46270482 | Charlson - Rheumatic | TRUE | 1 | Polyneuropathy due to systemic sclerosis | FALSE |
| CCI score | 765004404 | 80800 | Charlson - Rheumatic | TRUE | 1 | Polymyositis | FALSE |
| CCI score | 765004404 | 4035611 | Charlson - Rheumatic | TRUE | 1 | Seropositive rheumatoid arthritis | FALSE |
| CCI score | 765004404 | 4079978 | Charlson - Rheumatic | TRUE | 1 | Overlap syndrome | FALSE |
| CCI score | 765004404 | 4117687 | Charlson - Rheumatic | TRUE | 1 | Rheumatoid arthritis - ankle and/or foot | FALSE |
| CCI score | 765004404 | 4135937 | Charlson - Rheumatic | TRUE | 1 | CREST syndrome | FALSE |
| CCI score | 765004404 | 4216972 | Charlson - Rheumatic | TRUE | 1 | Bursitis of hip | FALSE |
| CCI score | 765004404 | 4285717 | Charlson - Rheumatic | TRUE | 1 | SLE glomerulonephritis syndrome | FALSE |
| CCI score | 765004404 | 37108590 | Charlson - Rheumatic | TRUE | 1 | Rheumatoid arthritis of left knee | FALSE |
| CCI score | 765004404 | 40485046 | Charlson - Rheumatic | TRUE | 1 | Progressive systemic sclerosis | FALSE |
| CCI score | 765004404 | 74125 | Charlson - Rheumatic | TRUE | 1 | Inflammatory polyarthropathy | FALSE |
| CCI score | 765004404 | 81097 | Charlson - Rheumatic | TRUE | 1 | Felty's syndrome | FALSE |
| CCI score | 765004404 | 134442 | Charlson - Rheumatic | TRUE | 1 | Systemic sclerosis | FALSE |
| CCI score | 765004404 | 255304 | Charlson - Rheumatic | TRUE | 1 | Lung disease with systemic sclerosis | FALSE |
| CCI score | 765004404 | 4063581 | Charlson - Rheumatic | TRUE | 1 | Drug-induced systemic lupus erythematosus | FALSE |
| CCI score | 765004404 | 4107913 | Charlson - Rheumatic | TRUE | 1 | Myopathy due to rheumatoid arthritis | FALSE |
| CCI score | 765004404 | 4116150 | Charlson - Rheumatic | TRUE | 1 | Rheumatoid arthritis of hip | FALSE |
| CCI score | 765004404 | 4117689 | Charlson - Rheumatic | TRUE | 1 | Bursitis of bursa of ankle and/or foot | FALSE |
| CCI score | 765004404 | 4184657 | Charlson - Rheumatic | TRUE | 1 | Bursitis of wrist | FALSE |
| CCI score | 765004404 | 4344158 | Charlson - Rheumatic | TRUE | 1 | Systemic lupus erythematosus with organ/system involvement | FALSE |
| CCI score | 765004404 | 35609009 | Charlson - Rheumatic | TRUE | 1 | Rheumatoid arthritis of left shoulder | FALSE |
| CCI score | 765004404 | 37108591 | Charlson - Rheumatic | TRUE | 1 | Rheumatoid arthritis of right knee | FALSE |
| CCI score | 765004404 | 37108607 | Charlson - Rheumatic | TRUE | 1 | Arthritis of right wrist | FALSE |
| CCI score | 765004404 | 37395588 | Charlson - Rheumatic | TRUE | 1 | Juvenile dermatomyositis co-occurrent with respiratory involvement | FALSE |
| CCI score | 765004404 | 77630 | Charlson - Rheumatic | TRUE | 1 | Disorder of shoulder | FALSE |
| CCI score | 765004404 | 4118008 | Charlson - Rheumatic | TRUE | 1 | Bursitis of multiple bursa | FALSE |
| CCI score | 765004404 | 4145240 | Charlson - Rheumatic | TRUE | 1 | Renal tubulo-interstitial disorder in systemic lupus erythematosus | FALSE |
| CCI score | 765004404 | 4211842 | Charlson - Rheumatic | TRUE | 1 | Bursitis of hand | FALSE |
| CCI score | 765004404 | 35609010 | Charlson - Rheumatic | TRUE | 1 | Rheumatoid arthritis of right shoulder | FALSE |
| CCI score | 765004404 | 46273162 | Charlson - Rheumatic | TRUE | 1 | Bursitis of elbow | FALSE |
| CCI score | 765004404 | 46273369 | Charlson - Rheumatic | TRUE | 1 | Endocarditis due to systemic lupus erythematosus | FALSE |
| CCI score | 765004404 | 80182 | Charlson - Rheumatic | TRUE | 1 | Dermatomyositis | FALSE |
| CCI score | 765004404 | 80809 | Charlson - Rheumatic | TRUE | 1 | Rheumatoid arthritis | FALSE |
| CCI score | 765004404 | 319825 | Charlson - Rheumatic | TRUE | 1 | Rheumatic heart disease | FALSE |
| CCI score | 765004404 | 4005037 | Charlson - Rheumatic | TRUE | 1 | Childhood type dermatomyositis | FALSE |
| CCI score | 765004404 | 4102493 | Charlson - Rheumatic | TRUE | 1 | Polyneuropathy in rheumatoid arthritis | FALSE |
| CCI score | 765004404 | 4115161 | Charlson - Rheumatic | TRUE | 1 | Rheumatoid arthritis - hand joint | FALSE |
| CCI score | 765004404 | 4142899 | Charlson - Rheumatic | TRUE | 1 | Subcutaneous rheumatoid nodule | FALSE |
| CCI score | 765004404 | 4343935 | Charlson - Rheumatic | TRUE | 1 | Giant cell arteritis with polymyalgia rheumatica | FALSE |
| CCI score | 765004404 | 4344161 | Charlson - Rheumatic | TRUE | 1 | Dermatomyositis with malignant disease | FALSE |
| CCI score | 765004404 | 4344258 | Charlson - Rheumatic | TRUE | 1 | Bursitis of shoulder | FALSE |
| CCI score | 765004404 | 36685020 | Charlson - Rheumatic | TRUE | 1 | Rheumatoid arthritis of left wrist | FALSE |
| CCI score | 765004404 | 36685024 | Charlson - Rheumatic | TRUE | 1 | Rheumatoid arthritis of right wrist | FALSE |
| CCI score | 765004404 | 37108605 | Charlson - Rheumatic | TRUE | 1 | Arthritis of left elbow | FALSE |
| CCI score | 765004404 | 37108606 | Charlson - Rheumatic | TRUE | 1 | Arthritis of left wrist | FALSE |
| CCI score | 765004404 | 37119224 | Charlson - Rheumatic | TRUE | 1 | Arthritis of right elbow | FALSE |
| CCI score | 765004404 | 42534836 | Charlson - Rheumatic | TRUE | 1 | Rheumatoid arthritis of right hand | FALSE |
| CCI score | 765004404 | 762446 | Charlson - Rheumatic | TRUE | 1 | Arthropathy of multiple joints | FALSE |
| CCI score | 765004404 | 4009619 | Charlson - Rheumatic | TRUE | 1 | Bursitis of knee | FALSE |
| CCI score | 765004404 | 4083556 | Charlson - Rheumatic | TRUE | 1 | Seronegative rheumatoid arthritis | FALSE |
| CCI score | 765004404 | 4105026 | Charlson - Rheumatic | TRUE | 1 | Myopathy due to systemic sclerosis | FALSE |
| CCI score | 765004404 | 4116151 | Charlson - Rheumatic | TRUE | 1 | Rheumatoid arthritis of knee | FALSE |
| CCI score | 765004404 | 4116440 | Charlson - Rheumatic | TRUE | 1 | Rheumatoid arthritis of elbow | FALSE |
| CCI score | 765004404 | 4116441 | Charlson - Rheumatic | TRUE | 1 | Rheumatoid arthritis of wrist | FALSE |
| CCI score | 765004404 | 4149913 | Charlson - Rheumatic | TRUE | 1 | Systemic lupus erythematosus with pericarditis | FALSE |
| CCI score | 765004404 | 36685022 | Charlson - Rheumatic | TRUE | 1 | Rheumatoid arthritis of right elbow | FALSE |
| CCI score | 765004404 | 42534835 | Charlson - Rheumatic | TRUE | 1 | Rheumatoid arthritis of left hip | FALSE |
| CCI score | 765004404 | 257628 | Charlson - Rheumatic | TRUE | 1 | Systemic lupus erythematosus | FALSE |
| CCI score | 765004404 | 4063582 | Charlson - Rheumatic | TRUE | 1 | Systemic sclerosis induced by drugs and chemicals | FALSE |
| CCI score | 765004404 | 4114439 | Charlson - Rheumatic | TRUE | 1 | Rheumatoid arthritis of shoulder | FALSE |
| CCI score | 765004404 | 4117686 | Charlson - Rheumatic | TRUE | 1 | Rheumatoid arthritis of multiple joints | FALSE |
| CCI score | 765004404 | 4305027 | Charlson - Rheumatic | TRUE | 1 | Finding of ankle or foot | FALSE |
| CCI score | 765004404 | 37108614 | Charlson - Rheumatic | TRUE | 1 | Arthritis of right ankle | FALSE |
| CCI score | 765004404 | 37395590 | Charlson - Rheumatic | TRUE | 1 | Rheumatoid lung disease with rheumatoid arthritis | FALSE |
| CCI score | 652711186 | 381591 | Charlson - Stroke | TRUE | 1 | Cerebrovascular disease | FALSE |
| CCI score | 652711186 | 434657 | Charlson - Stroke | TRUE | 1 | Weakness of face muscles | FALSE |
| CCI score | 652711186 | 436430 | Charlson - Stroke | TRUE | 1 | Nontraumatic extradural hemorrhage | FALSE |
| CCI score | 652711186 | 443551 | Charlson - Stroke | TRUE | 1 | Apraxia due to cerebrovascular accident | FALSE |
| CCI score | 652711186 | 443599 | Charlson - Stroke | TRUE | 1 | Paralytic syndrome of nondominant side as late effect of stroke | FALSE |
| CCI score | 652711186 | 443916 | Charlson - Stroke | TRUE | 1 | Hereditary disease | FALSE |
| CCI score | 652711186 | 4027461 | Charlson - Stroke | TRUE | 1 | Leukoencephalopathy | FALSE |
| CCI score | 652711186 | 4110192 | Charlson - Stroke | TRUE | 1 | Cerebral infarction due to thrombosis of cerebral arteries | FALSE |
| CCI score | 652711186 | 4110194 | Charlson - Stroke | TRUE | 1 | Middle cerebral artery syndrome | FALSE |
| CCI score | 652711186 | 4111708 | Charlson - Stroke | TRUE | 1 | Subarachnoid hemorrhage from vertebral artery | FALSE |
| CCI score | 652711186 | 4164092 | Charlson - Stroke | TRUE | 1 | Acute cerebrovascular insufficiency | FALSE |
| CCI score | 652711186 | 4190891 | Charlson - Stroke | TRUE | 1 | Cerebral autosomal dominant arteriopathy with subcortical infarcts and leukoencephalopathy | FALSE |
| CCI score | 652711186 | 40479575 | Charlson - Stroke | TRUE | 1 | Dysphasia as late effect of cerebrovascular disease | FALSE |
| CCI score | 652711186 | 40480002 | Charlson - Stroke | TRUE | 1 | Aphasia as late effect of cerebrovascular disease | FALSE |
| CCI score | 652711186 | 43530727 | Charlson - Stroke | TRUE | 1 | Spontaneous cerebral hemorrhage | FALSE |
| CCI score | 652711186 | 43530742 | Charlson - Stroke | TRUE | 1 | Paralytic syndrome on one side of the body as late effect of cerebrovascular accident | FALSE |
| CCI score | 652711186 | 43530744 | Charlson - Stroke | TRUE | 1 | Weakness of face muscles as sequela of stroke | FALSE |
| CCI score | 652711186 | 45772786 | Charlson - Stroke | TRUE | 1 | Cerebral infarction due to embolism of middle cerebral artery | FALSE |
| CCI score | 652711186 | 45773220 | Charlson - Stroke | TRUE | 1 | Reversible cerebral vasoconstriction syndrome | FALSE |
| CCI score | 652711186 | 197303 | Charlson - Stroke | TRUE | 1 | Monoplegia of dominant lower limb as a late effect of cerebrovascular accident | FALSE |
| CCI score | 652711186 | 314667 | Charlson - Stroke | TRUE | 1 | Nonpyogenic thrombosis of intracranial venous sinus | FALSE |
| CCI score | 652711186 | 372654 | Charlson - Stroke | TRUE | 1 | Paralytic syndrome as late effect of stroke | FALSE |
| CCI score | 652711186 | 374055 | Charlson - Stroke | TRUE | 1 | Basilar artery syndrome | FALSE |
| CCI score | 652711186 | 374384 | Charlson - Stroke | TRUE | 1 | Cerebral ischemia | FALSE |
| CCI score | 652711186 | 432923 | Charlson - Stroke | TRUE | 1 | Subarachnoid hemorrhage | FALSE |
| CCI score | 652711186 | 4043731 | Charlson - Stroke | TRUE | 1 | Infarction - precerebral | FALSE |
| CCI score | 652711186 | 4045738 | Charlson - Stroke | TRUE | 1 | Pure sensory lacunar infarction | FALSE |
| CCI score | 652711186 | 4111717 | Charlson - Stroke | TRUE | 1 | Occlusion and stenosis of posterior cerebral artery | FALSE |
| CCI score | 652711186 | 4111720 | Charlson - Stroke | TRUE | 1 | Sequelae of subarachnoid hemorrhage | FALSE |
| CCI score | 652711186 | 4153380 | Charlson - Stroke | TRUE | 1 | Disorder of carotid artery | FALSE |
| CCI score | 652711186 | 40481842 | Charlson - Stroke | TRUE | 1 | Monoplegia of upper limb as late effect of cerebrovascular disease | FALSE |
| CCI score | 652711186 | 312938 | Charlson - Stroke | TRUE | 1 | Hypertensive encephalopathy | FALSE |
| CCI score | 652711186 | 443454 | Charlson - Stroke | TRUE | 1 | Cerebral infarction | FALSE |
| CCI score | 652711186 | 443525 | Charlson - Stroke | TRUE | 1 | Monoplegia of dominant upper limb as a late effect of cerebrovascular accident | FALSE |
| CCI score | 652711186 | 4045737 | Charlson - Stroke | TRUE | 1 | Pure motor lacunar infarction | FALSE |
| CCI score | 652711186 | 4045749 | Charlson - Stroke | TRUE | 1 | Cerebral amyloid angiopathy | FALSE |
| CCI score | 652711186 | 4049659 | Charlson - Stroke | TRUE | 1 | Subcortical hemorrhage | FALSE |
| CCI score | 652711186 | 4110186 | Charlson - Stroke | TRUE | 1 | Intracerebral hemorrhage, multiple localized | FALSE |
| CCI score | 652711186 | 4110190 | Charlson - Stroke | TRUE | 1 | Cerebral infarction due to embolism of precerebral arteries | FALSE |
| CCI score | 652711186 | 4110195 | Charlson - Stroke | TRUE | 1 | Posterior cerebral artery syndrome | FALSE |
| CCI score | 652711186 | 4353709 | Charlson - Stroke | TRUE | 1 | Intracerebral vascular finding | FALSE |
| CCI score | 652711186 | 40480946 | Charlson - Stroke | TRUE | 1 | Monoplegia of nondominant lower limb as a late effect of cerebrovascular accident | FALSE |
| CCI score | 652711186 | 43022059 | Charlson - Stroke | TRUE | 1 | Disease of non-coronary systemic artery | FALSE |
| CCI score | 652711186 | 43530851 | Charlson - Stroke | TRUE | 1 | Chronic non-traumatic intracranial subdural hemorrhage | FALSE |
| CCI score | 652711186 | 321887 | Charlson - Stroke | TRUE | 1 | Disorder of artery | FALSE |
| CCI score | 652711186 | 380747 | Charlson - Stroke | TRUE | 1 | Cerebral arteritis | FALSE |
| CCI score | 652711186 | 381036 | Charlson - Stroke | TRUE | 1 | Multiple AND bilateral precerebral artery stenosis | FALSE |
| CCI score | 652711186 | 433195 | Charlson - Stroke | TRUE | 1 | Transient arterial retinal occlusion | FALSE |
| CCI score | 652711186 | 434056 | Charlson - Stroke | TRUE | 1 | Late effects of cerebrovascular disease | FALSE |
| CCI score | 652711186 | 437306 | Charlson - Stroke | TRUE | 1 | Transient global amnesia | FALSE |
| CCI score | 652711186 | 443465 | Charlson - Stroke | TRUE | 1 | Dysphagia as a late effect of cerebrovascular accident | FALSE |
| CCI score | 652711186 | 4111709 | Charlson - Stroke | TRUE | 1 | Non-traumatic subdural hemorrhage | FALSE |
| CCI score | 652711186 | 4111711 | Charlson - Stroke | TRUE | 1 | Cerebellar stroke syndrome | FALSE |
| CCI score | 652711186 | 4111715 | Charlson - Stroke | TRUE | 1 | Occlusion and stenosis of cerebral arteries, not resulting in cerebral infarction | FALSE |
| CCI score | 652711186 | 4111716 | Charlson - Stroke | TRUE | 1 | Occlusion and stenosis of anterior cerebral artery | FALSE |
| CCI score | 652711186 | 4112023 | Charlson - Stroke | TRUE | 1 | Occlusion and stenosis of middle cerebral artery | FALSE |
| CCI score | 652711186 | 4112024 | Charlson - Stroke | TRUE | 1 | Occlusion and stenosis of cerebellar arteries | FALSE |
| CCI score | 652711186 | 4144154 | Charlson - Stroke | TRUE | 1 | Non-traumatic intracerebral ventricular hemorrhage | FALSE |
| CCI score | 652711186 | 4148906 | Charlson - Stroke | TRUE | 1 | Spontaneous subarachnoid hemorrhage | FALSE |
| CCI score | 652711186 | 4319328 | Charlson - Stroke | TRUE | 1 | Brain stem hemorrhage | FALSE |
| CCI score | 652711186 | 4332246 | Charlson - Stroke | TRUE | 1 | Aneurysm | FALSE |
| CCI score | 652711186 | 40480938 | Charlson - Stroke | TRUE | 1 | Monoplegia of lower limb as late effect of cerebrovascular disease | FALSE |
| CCI score | 652711186 | 40481354 | Charlson - Stroke | TRUE | 1 | Speech and language deficit as late effect of cerebrovascular accident | FALSE |
| CCI score | 652711186 | 42535425 | Charlson - Stroke | TRUE | 1 | Spontaneous hemorrhage of cerebral hemisphere | FALSE |
| CCI score | 652711186 | 43530674 | Charlson - Stroke | TRUE | 1 | Spontaneous cerebellar hemorrhage | FALSE |
| CCI score | 652711186 | 46273649 | Charlson - Stroke | TRUE | 1 | Cerebral infarction due to occlusion of basilar artery | FALSE |
| CCI score | 652711186 | 373503 | Charlson - Stroke | TRUE | 1 | Transient cerebral ischemia | FALSE |
| CCI score | 652711186 | 378774 | Charlson - Stroke | TRUE | 1 | Moyamoya disease | FALSE |
| CCI score | 652711186 | 443609 | Charlson - Stroke | TRUE | 1 | Paralytic syndrome of dominant side as late effect of stroke | FALSE |
| CCI score | 652711186 | 4006294 | Charlson - Stroke | TRUE | 1 | Basilar artery embolism | FALSE |
| CCI score | 652711186 | 4046360 | Charlson - Stroke | TRUE | 1 | Lacunar infarction | FALSE |
| CCI score | 652711186 | 4111710 | Charlson - Stroke | TRUE | 1 | Brainstem stroke syndrome | FALSE |
| CCI score | 652711186 | 4112020 | Charlson - Stroke | TRUE | 1 | Carotid artery syndrome hemispheric | FALSE |
| CCI score | 652711186 | 4112026 | Charlson - Stroke | TRUE | 1 | Sequelae of cerebral infarction | FALSE |
| CCI score | 652711186 | 4180158 | Charlson - Stroke | TRUE | 1 | Hereditary disorder by system | FALSE |
| CCI score | 652711186 | 4274969 | Charlson - Stroke | TRUE | 1 | Vertebral artery embolism | FALSE |
| CCI score | 652711186 | 4338227 | Charlson - Stroke | TRUE | 1 | Basilar artery thrombosis | FALSE |
| CCI score | 652711186 | 37016924 | Charlson - Stroke | TRUE | 1 | Dissection of cerebral artery | FALSE |
| CCI score | 652711186 | 40482266 | Charlson - Stroke | TRUE | 1 | Monoplegia of nondominant upper limb as a late effect of cerebrovascular accident | FALSE |
| CCI score | 652711186 | 42872891 | Charlson - Stroke | TRUE | 1 | Posterior reversible encephalopathy syndrome | FALSE |
| CCI score | 652711186 | 43530687 | Charlson - Stroke | TRUE | 1 | Dysarthria as late effects of cerebrovascular disease | FALSE |
| CCI score | 652711186 | 43530688 | Charlson - Stroke | TRUE | 1 | Fluency disorder as sequela of cerebrovascular disease | FALSE |
| CCI score | 652711186 | 43531622 | Charlson - Stroke | TRUE | 1 | Ataxia as sequela of cerebrovascular disease | FALSE |
| CCI score | 652711186 | 45767658 | Charlson - Stroke | TRUE | 1 | Cerebral infarction due to thrombosis of middle cerebral artery | FALSE |
| CCI score | 652711186 | 255919 | Charlson - Stroke | TRUE | 1 | Finding of head and neck region | FALSE |
| CCI score | 652711186 | 313226 | Charlson - Stroke | TRUE | 1 | Carotid artery occlusion | FALSE |
| CCI score | 652711186 | 316437 | Charlson - Stroke | TRUE | 1 | Cerebral atherosclerosis | FALSE |
| CCI score | 652711186 | 4108360 | Charlson - Stroke | TRUE | 1 | Anterior cerebral artery syndrome | FALSE |
| CCI score | 652711186 | 4108952 | Charlson - Stroke | TRUE | 1 | Subarachnoid hemorrhage from carotid siphon and bifurcation | FALSE |
| CCI score | 652711186 | 4110189 | Charlson - Stroke | TRUE | 1 | Cerebral infarct due to thrombosis of precerebral arteries | FALSE |
| CCI score | 652711186 | 4111714 | Charlson - Stroke | TRUE | 1 | Cerebral infarction due to cerebral venous thrombosis, non-pyogenic | FALSE |
| CCI score | 652711186 | 4111721 | Charlson - Stroke | TRUE | 1 | Sequelae of intracerebral hemorrhage | FALSE |
| CCI score | 652711186 | 4159164 | Charlson - Stroke | TRUE | 1 | Disorder of basilar artery | FALSE |
| CCI score | 652711186 | 4176892 | Charlson - Stroke | TRUE | 1 | Cortical hemorrhage | FALSE |
| CCI score | 652711186 | 4213731 | Charlson - Stroke | TRUE | 1 | Carotid artery embolism | FALSE |
| CCI score | 652711186 | 4273526 | Charlson - Stroke | TRUE | 1 | Vertebral artery thrombosis | FALSE |
| CCI score | 652711186 | 4311124 | Charlson - Stroke | TRUE | 1 | Carotid artery thrombosis | FALSE |
| CCI score | 652711186 | 40482301 | Charlson - Stroke | TRUE | 1 | Residual cognitive deficit as late effect of cerebrovascular accident | FALSE |
| CCI score | 652711186 | 44782781 | Charlson - Stroke | TRUE | 1 | Hemiplegia and/or hemiparesis following stroke | FALSE |
| CCI score | 652711186 | 45766121 | Charlson - Stroke | TRUE | 1 | Cerebral vasoconstriction syndrome | FALSE |
| CCI score | 652711186 | 46270031 | Charlson - Stroke | TRUE | 1 | Cerebral infarction due to occlusion of precerebral artery | FALSE |
| CCI score | 652711186 | 372924 | Charlson - Stroke | TRUE | 1 | Cerebral artery occlusion | FALSE |
| CCI score | 652711186 | 443239 | Charlson - Stroke | TRUE | 1 | Precerebral arterial occlusion | FALSE |
| CCI score | 652711186 | 4108356 | Charlson - Stroke | TRUE | 1 | Cerebral infarction due to embolism of cerebral arteries | FALSE |
| CCI score | 652711186 | 4288310 | Charlson - Stroke | TRUE | 1 | Carotid artery obstruction | FALSE |
| CCI score | 652711186 | 4338523 | Charlson - Stroke | TRUE | 1 | Amaurosis fugax | FALSE |
| CCI score | 652711186 | 44782753 | Charlson - Stroke | TRUE | 1 | Weakness as a late effect of stroke | FALSE |
| CCI score | 382527336 | 439727 | hiv infection | TRUE | 2 | Human immunodeficiency virus infection | FALSE |
| CCI score | 382527336 | 4013106 | hiv infection | TRUE | 2 | HIV positive | FALSE |
| CCI score | 382527336 | 4047624 | hiv infection | TRUE | 2 | Human immunodeficiency virus leukoencephalopathy | FALSE |
| CCI score | 382527336 | 4087603 | hiv infection | TRUE | 2 | Human immunodeficiency virus infection constitutional disease | FALSE |
| CCI score | 382527336 | 4172009 | hiv infection | TRUE | 2 | HIV infection with infectious mononucleosis-like syndrome | FALSE |
| CCI score | 382527336 | 4201627 | hiv infection | TRUE | 2 | HIV infection with aseptic meningitis | FALSE |
| CCI score | 382527336 | 4262297 | hiv infection | TRUE | 2 | Human immunodeficiency virus encephalopathy | FALSE |
| CCI score | 382527336 | 36674253 | hiv infection | TRUE | 2 | Infection caused by Toxoplasma gondii co-occurrent with acquired immunodeficiency syndrome | FALSE |
| CCI score | 382527336 | 37017092 | hiv infection | TRUE | 2 | Candidiasis of esophagus co-occurrent with human immunodeficiency virus infection | FALSE |
| CCI score | 382527336 | 37017124 | hiv infection | TRUE | 2 | Infection caused by Strongyloides co-occurrent with human immunodeficiency virus infection | FALSE |
| CCI score | 382527336 | 37017260 | hiv infection | TRUE | 2 | Disorder of kidney co-occurrent with human immunodeficiency virus infection | FALSE |
| CCI score | 382527336 | 37017261 | hiv infection | TRUE | 2 | Gastrointestinal malabsorption syndrome co-occurrent with human immunodeficiency virus infection | FALSE |
| CCI score | 382527336 | 37017276 | hiv infection | TRUE | 2 | Cardiomyopathy co-occurrent with human immunodeficiency virus infection | FALSE |
| CCI score | 382527336 | 37017283 | hiv infection | TRUE | 2 | Visual impairment co-occurrent with human immunodeficiency virus infection | FALSE |
| CCI score | 382527336 | 37017320 | hiv infection | TRUE | 2 | Malignant neoplastic disease co-occurrent with human immunodeficiency virus infection | FALSE |
| CCI score | 382527336 | 37017424 | hiv infection | TRUE | 2 | Nephrotic syndrome co-occurrent with human immunodeficiency virus infection | FALSE |
| CCI score | 382527336 | 37017446 | hiv infection | TRUE | 2 | Infection caused by Cytomegalovirus co-occurrent with human immunodeficiency virus infection | FALSE |
| CCI score | 382527336 | 37017457 | hiv infection | TRUE | 2 | Infection caused by herpes zoster virus co-occurrent with human immunodeficiency virus infection | FALSE |
| CCI score | 382527336 | 37017549 | hiv infection | TRUE | 2 | Dementia co-occurrent with human immunodeficiency virus infection | FALSE |
| CCI score | 382527336 | 37019058 | hiv infection | TRUE | 2 | Bacterial pneumonia co-occurrent with human immunodeficiency virus infection | FALSE |
| CCI score | 382527336 | 42536595 | hiv infection | TRUE | 2 | Human immunodeficiency virus World Health Organization 2007 stage 4 co-occurrent with tuberculosis | FALSE |
| CCI score | 382527336 | 42538959 | hiv infection | TRUE | 2 | Asymptomatic human immunodeficiency virus A1 infection | FALSE |
| CCI score | 382527336 | 44783356 | hiv infection | TRUE | 2 | Human immunodeficiency virus carrier | FALSE |
| CCI score | 382527336 | 45757102 | hiv infection | TRUE | 2 | Acquired immune deficiency syndrome complicating childbirth | FALSE |
| CCI score | 382527336 | 432554 | hiv infection | TRUE | 2 | Human immunodeficiency virus II infection | FALSE |
| CCI score | 382527336 | 4008081 | hiv infection | TRUE | 2 | Acute HIV infection | FALSE |
| CCI score | 382527336 | 36674254 | hiv infection | TRUE | 2 | Infection caused by Isospora co-occurrent with acquired immunodeficiency syndrome | FALSE |
| CCI score | 382527336 | 36687122 | hiv infection | TRUE | 2 | Human immunodeficiency virus infection with cognitive impairment | FALSE |
| CCI score | 382527336 | 37017071 | hiv infection | TRUE | 2 | Subacute adenoviral encephalitis co-occurrent with human immunodeficiency virus infection | FALSE |
| CCI score | 382527336 | 37017082 | hiv infection | TRUE | 2 | Splenomegaly co-occurrent with human immunodeficiency virus infection | FALSE |
| CCI score | 382527336 | 37017093 | hiv infection | TRUE | 2 | Heart disease co-occurrent with human immunodeficiency virus infection | FALSE |
| CCI score | 382527336 | 37017108 | hiv infection | TRUE | 2 | Myocarditis co-occurrent with human immunodeficiency virus infection | FALSE |
| CCI score | 382527336 | 37017249 | hiv infection | TRUE | 2 | Organic brain syndrome co-occurrent with human immunodeficiency virus infection | FALSE |
| CCI score | 382527336 | 37017279 | hiv infection | TRUE | 2 | Disorder of peripheral nervous system co-occurrent with human immunodeficiency virus infection | FALSE |
| CCI score | 382527336 | 37017284 | hiv infection | TRUE | 2 | Infective arthritis co-occurrent with human immunodeficiency virus infection | FALSE |
| CCI score | 382527336 | 37017285 | hiv infection | TRUE | 2 | Acquired hemolytic anemia co-occurrent with human immunodeficiency virus infection | FALSE |
| CCI score | 382527336 | 37017295 | hiv infection | TRUE | 2 | Infection caused by Nocardia co-occurrent with human immunodeficiency virus infection | FALSE |
| CCI score | 382527336 | 37017454 | hiv infection | TRUE | 2 | Infection caused by herpes simplex virus co-occurrent with human immunodeficiency virus infection | FALSE |
| CCI score | 382527336 | 37017455 | hiv infection | TRUE | 2 | Pyrexia of unknown origin co-occurrent with human immunodeficiency virus infection | FALSE |
| CCI score | 382527336 | 37017456 | hiv infection | TRUE | 2 | Aspergillosis co-occurrent with human immunodeficiency virus infection | FALSE |
| CCI score | 382527336 | 37017579 | hiv infection | TRUE | 2 | Opportunistic mycosis co-occurrent with human immunodeficiency virus infection | FALSE |
| CCI score | 382527336 | 37017586 | hiv infection | TRUE | 2 | Focal segmental glomerulosclerosis co-occurrent with human immunodeficiency virus infection | FALSE |
| CCI score | 382527336 | 37017655 | hiv infection | TRUE | 2 | Disseminated atypical infection caused by Mycobacterium co-occurrent with human immunodeficiency virus infection | FALSE |
| CCI score | 382527336 | 37018711 | hiv infection | TRUE | 2 | Disorder of eye proper co-occurrent with human immunodeficiency virus infection | FALSE |
| CCI score | 382527336 | 37018935 | hiv infection | TRUE | 2 | Dermatophytosis co-occurrent with human immunodeficiency virus infection | FALSE |
| CCI score | 382527336 | 42538960 | hiv infection | TRUE | 2 | Asymptomatic human immunodeficiency virus A2 infection | FALSE |
| CCI score | 382527336 | 42539031 | hiv infection | TRUE | 2 | Human immunodeficiency virus World Health Organization 2007 stage 3 co-occurrent with tuberculosis | FALSE |
| CCI score | 382527336 | 43531586 | hiv infection | TRUE | 2 | Symptomatic human immunodeficiency virus infection | FALSE |
| CCI score | 382527336 | 4180254 | hiv infection | TRUE | 2 | Congenital human immunodeficiency virus infection | FALSE |
| CCI score | 382527336 | 4239722 | hiv infection | TRUE | 2 | Asymptomatic human immunodeficiency virus infection in pregnancy | FALSE |
| CCI score | 382527336 | 36674252 | hiv infection | TRUE | 2 | Infection caused by Coccidia co-occurrent with acquired immunodeficiency syndrome | FALSE |
| CCI score | 382527336 | 36715476 | hiv infection | TRUE | 2 | Human immunodeficiency virus complicating pregnancy childbirth and the puerperium | FALSE |
| CCI score | 382527336 | 36716524 | hiv infection | TRUE | 2 | Parkinsonism due to human immunodeficiency virus infection | FALSE |
| CCI score | 382527336 | 37017126 | hiv infection | TRUE | 2 | Myelitis co-occurrent with human immunodeficiency virus infection | FALSE |
| CCI score | 382527336 | 37017132 | hiv infection | TRUE | 2 | Anemia co-occurrent with human immunodeficiency virus infection | FALSE |
| CCI score | 382527336 | 37017247 | hiv infection | TRUE | 2 | Presenile dementia co-occurrent with human immunodeficiency virus infection | FALSE |
| CCI score | 382527336 | 37017248 | hiv infection | TRUE | 2 | Infection caused by Pneumocystis co-occurrent with human immunodeficiency virus infection | FALSE |
| CCI score | 382527336 | 37017259 | hiv infection | TRUE | 2 | Disorder of spinal cord co-occurrent with human immunodeficiency virus infection | FALSE |
| CCI score | 382527336 | 37018063 | hiv infection | TRUE | 2 | Immune reconstitution inflammatory syndrome caused by human immunodeficiency virus infection | FALSE |
| CCI score | 382527336 | 37018714 | hiv infection | TRUE | 2 | Radiculitis co-occurrent with human immunodeficiency virus infection | FALSE |
| CCI score | 382527336 | 37019034 | hiv infection | TRUE | 2 | Neuralgia co-occurrent with human immunodeficiency virus infection | FALSE |
| CCI score | 382527336 | 37116830 | hiv infection | TRUE | 2 | Invasive carcinoma of uterine cervix co-occurrent with human immunodeficiency virus infection | FALSE |
| CCI score | 382527336 | 42536594 | hiv infection | TRUE | 2 | Human immunodeficiency virus World Health Organization 2007 stage 3 co-occurrent with malaria | FALSE |
| CCI score | 382527336 | 42536596 | hiv infection | TRUE | 2 | Human immunodeficiency virus World Health Organization 2007 stage 4 co-occurrent with malaria | FALSE |
| CCI score | 382527336 | 45769839 | hiv infection | TRUE | 2 | Human immunodeficiency virus (HIV) II infection category B1 | FALSE |
| CCI score | 382527336 | 45769856 | hiv infection | TRUE | 2 | Human immunodeficiency virus (HIV) infection category B2 | FALSE |
| CCI score | 382527336 | 46284256 | hiv infection | TRUE | 2 | HIV (human immunodeficiency virus) disease resulting in haematological and immunological abnormalities | FALSE |
| CCI score | 382527336 | 4048033 | hiv infection | TRUE | 2 | Neuropathy due to human immunodeficiency virus | FALSE |
| CCI score | 382527336 | 4124361 | hiv infection | TRUE | 2 | Human immunodeficiency virus-associated periodontitis | FALSE |
| CCI score | 382527336 | 37017125 | hiv infection | TRUE | 2 | Disorder of skin co-occurrent with human immunodeficiency virus infection | FALSE |
| CCI score | 382527336 | 37017209 | hiv infection | TRUE | 2 | Hemophagocytic syndrome co-occurrent with human immunodeficiency virus infection | FALSE |
| CCI score | 382527336 | 37017246 | hiv infection | TRUE | 2 | Progressive multifocal leukoencephalopathy co-occurrent with human immunodeficiency virus infection | FALSE |
| CCI score | 382527336 | 37017266 | hiv infection | TRUE | 2 | Acute endocarditis co-occurrent with human immunodeficiency virus infection | FALSE |
| CCI score | 382527336 | 37017282 | hiv infection | TRUE | 2 | Agranulocytosis co-occurrent with human immunodeficiency virus infection | FALSE |
| CCI score | 382527336 | 37017318 | hiv infection | TRUE | 2 | Infectious gastroenteritis co-occurrent with human immunodeficiency virus infection | FALSE |
| CCI score | 382527336 | 37017442 | hiv infection | TRUE | 2 | Diffuse non-Hodgkin immunoblastic lymphoma co-occurrent with human immunodeficiency virus infection | FALSE |
| CCI score | 382527336 | 37017453 | hiv infection | TRUE | 2 | Coccidiosis co-occurrent with human immunodeficiency virus infection | FALSE |
| CCI score | 382527336 | 37017652 | hiv infection | TRUE | 2 | Multidermatomal infection caused by Herpes zoster co-occurrent with human immunodeficiency virus infection | FALSE |
| CCI score | 382527336 | 37019055 | hiv infection | TRUE | 2 | Aplastic anemia co-occurrent with human immunodeficiency virus infection | FALSE |
| CCI score | 382527336 | 37116831 | hiv infection | TRUE | 2 | Extrapulmonary tuberculosis co-occurrent with human immunodeficiency virus infection | FALSE |
| CCI score | 382527336 | 42536592 | hiv infection | TRUE | 2 | Human immunodeficiency virus World Health Organization 2007 stage 2 co-occurrent with tuberculosis | FALSE |
| CCI score | 382527336 | 4087604 | hiv infection | TRUE | 2 | Human immunodeficiency virus infection with neurological disease | FALSE |
| CCI score | 382527336 | 4092686 | hiv infection | TRUE | 2 | Human immunodeficiency virus infection with secondary clinical infectious disease | FALSE |
| CCI score | 382527336 | 4161950 | hiv infection | TRUE | 2 | Human immunodeficiency virus encephalitis | FALSE |
| CCI score | 382527336 | 4171124 | hiv infection | TRUE | 2 | Congenital acquired immune deficiency syndrome | FALSE |
| CCI score | 382527336 | 4236860 | hiv infection | TRUE | 2 | Pediatric human immunodeficiency virus infection | FALSE |
| CCI score | 382527336 | 36714339 | hiv infection | TRUE | 2 | Candidiasis of upper respiratory tract co-occurrent with human immunodeficiency virus infection | FALSE |
| CCI score | 382527336 | 37017094 | hiv infection | TRUE | 2 | Disorder of gastrointestinal tract co-occurrent with human immunodeficiency virus infection | FALSE |
| CCI score | 382527336 | 37017106 | hiv infection | TRUE | 2 | Eruption of skin co-occurrent with human immunodeficiency virus infection | FALSE |
| CCI score | 382527336 | 37017112 | hiv infection | TRUE | 2 | Primary cerebral lymphoma co-occurrent with human immunodeficiency virus infection | FALSE |
| CCI score | 382527336 | 37017210 | hiv infection | TRUE | 2 | Chronic infection caused by herpes simplex virus co-occurrent with human immunodeficiency virus infection | FALSE |
| CCI score | 382527336 | 37017243 | hiv infection | TRUE | 2 | Reticulosarcoma co-occurrent with human immunodeficiency virus infection | FALSE |
| CCI score | 382527336 | 37017265 | hiv infection | TRUE | 2 | Enlargement of liver co-occurrent with human immunodeficiency virus infection | FALSE |
| CCI score | 382527336 | 37017294 | hiv infection | TRUE | 2 | Demyelinating disease of central nervous system co-occurrent with human immunodeficiency virus infection | FALSE |
| CCI score | 382527336 | 37017296 | hiv infection | TRUE | 2 | Isosporiasis co-occurrent with human immunodeficiency virus infection | FALSE |
| CCI score | 382527336 | 37017465 | hiv infection | TRUE | 2 | Human immunodeficiency virus antibody positive | FALSE |
| CCI score | 382527336 | 37017580 | hiv infection | TRUE | 2 | Microsporidiosis co-occurrent with human immunodeficiency virus infection | FALSE |
| CCI score | 382527336 | 37017595 | hiv infection | TRUE | 2 | Burkitt lymphoma co-occurrent with human immunodeficiency virus infection | FALSE |
| CCI score | 382527336 | 42536590 | hiv infection | TRUE | 2 | Human immunodeficiency virus World Health Organization 2007 stage 1 co-occurrent with tuberculosis | FALSE |
| CCI score | 382527336 | 45757132 | hiv infection | TRUE | 2 | Human immunodeficiency virus in mother complicating childbirth | FALSE |
| CCI score | 382527336 | 45769855 | hiv infection | TRUE | 2 | Human immunodeficiency virus (HIV) infection category B1 | FALSE |
| CCI score | 382527336 | 45773072 | hiv infection | TRUE | 2 | Human immunodeficiency virus (HIV) II infection category B2 | FALSE |
| CCI score | 382527336 | 4128060 | hiv infection | TRUE | 2 | Acquired immune deficiency syndrome-related nephropathy | FALSE |
| CCI score | 382527336 | 4225193 | hiv infection | TRUE | 2 | Microsporidiosis associated with acquired immunodeficiency syndrome | FALSE |
| CCI score | 382527336 | 4253472 | hiv infection | TRUE | 2 | Human immunodeficiency virus I infection | FALSE |
| CCI score | 382527336 | 4314426 | hiv infection | TRUE | 2 | HIV infection with acute lymphadenitis | FALSE |
| CCI score | 382527336 | 4340791 | hiv infection | TRUE | 2 | Human immunodeficiency virus enteropathy | FALSE |
| CCI score | 382527336 | 37017244 | hiv infection | TRUE | 2 | Disorder of respiratory system co-occurrent with human immunodeficiency virus infection | FALSE |
| CCI score | 382527336 | 37017278 | hiv infection | TRUE | 2 | Recurrent bacterial pneumonia co-occurrent with human immunodeficiency virus infection | FALSE |
| CCI score | 382527336 | 37017319 | hiv infection | TRUE | 2 | Disorder of central nervous system co-occurrent with human immunodeficiency virus infection | FALSE |
| CCI score | 382527336 | 37017425 | hiv infection | TRUE | 2 | Renal failure syndrome co-occurrent with human immunodeficiency virus infection | FALSE |
| CCI score | 382527336 | 37017550 | hiv infection | TRUE | 2 | Infection caused by Cryptosporidium co-occurrent with human immunodeficiency virus infection | FALSE |
| CCI score | 382527336 | 37018721 | hiv infection | TRUE | 2 | Polyneuropathy co-occurrent with human immunodeficiency virus infection | FALSE |
| CCI score | 382527336 | 37018755 | hiv infection | TRUE | 2 | Recurrent salmonella sepsis co-occurrent with human immunodeficiency virus infection | FALSE |
| CCI score | 382527336 | 37019042 | hiv infection | TRUE | 2 | Infection caused by Salmonella co-occurrent with human immunodeficiency virus infection | FALSE |
| CCI score | 382527336 | 37019052 | hiv infection | TRUE | 2 | Disseminated infection caused by Strongyloides co-occurrent with human immunodeficiency virus infection | FALSE |
| CCI score | 382527336 | 42536593 | hiv infection | TRUE | 2 | Human immunodeficiency virus World Health Organization 2007 stage 2 co-occurrent with malaria | FALSE |
| CCI score | 382527336 | 45769864 | hiv infection | TRUE | 2 | Symptomatic human immunodeficiency virus (HIV) I infection | FALSE |
| CCI score | 382527336 | 4171125 | hiv infection | TRUE | 2 | Congenital human immunodeficiency virus positive status syndrome | FALSE |
| CCI score | 382527336 | 4241530 | hiv infection | TRUE | 2 | Asymptomatic human immunodeficiency virus infection | FALSE |
| CCI score | 382527336 | 4267414 | hiv infection | TRUE | 2 | AIDS | FALSE |
| CCI score | 382527336 | 4320032 | hiv infection | TRUE | 2 | Persistent generalized lymphadenopathy | FALSE |
| CCI score | 382527336 | 4347288 | hiv infection | TRUE | 2 | Human immunodeficiency virus myopathy | FALSE |
| CCI score | 382527336 | 37017254 | hiv infection | TRUE | 2 | Candidiasis of mouth co-occurrent with human immunodeficiency virus infection | FALSE |
| CCI score | 382527336 | 37017262 | hiv infection | TRUE | 2 | Neuritis co-occurrent with human immunodeficiency virus infection | FALSE |
| CCI score | 382527336 | 37017263 | hiv infection | TRUE | 2 | Lymphadenopathy co-occurrent with human immunodeficiency virus infection | FALSE |
| CCI score | 382527336 | 42536591 | hiv infection | TRUE | 2 | Human immunodeficiency virus World Health Organization 2007 stage 1 co-occurrent with malaria | FALSE |

Abbreviations: CCI: Charlson Comorbidity Index

Appendix A2: Qualifying Cognitive Symptoms and OMOP Codes

Table of qualifying Cognitive Long COVID symptoms

| phenotype_id | phenotype_label | omop_concept_id | omop_concept_label | omop_concept_code | omop_standard | omop_domain | omop_vocabulary |
| --- | --- | --- | --- | --- | --- | --- | --- |
| HP_0001289 | confusion | 4164633 | Clouded consciousness | 40917007 | S | Conditions | SNOMED |
| HP_0001289 | confusion | 381273 | Confusional state | 286933003 | S | Conditions | SNOMED |
| HP_0001289 | confusion | 4268911 | Disorientated | 62476001 | S | Conditions | SNOMED |
| HP_0001289 | confusion | 4056930 | Disorientated in time | 19657006 | S | Conditions | SNOMED |
| HP_0001289 | confusion | 4091700 | Dissociative confusion | 280943007 | S | Conditions | SNOMED |
| HP_0001289 | confusion | 441540 | Reactive confusion | 191678001 | S | Conditions | SNOMED |
| HP_0002354 | memory impairment | 439147 | Amnesia | 48167000 | S | Conditions | SNOMED |
| HP_0002354 | memory impairment | 4206332 | Forgetful | 55533009 | S | Conditions | SNOMED |
| HP_0002354 | memory impairment | 4304008 | Memory impairment | 386807006 | S | Conditions | SNOMED |
| HP_0002354 | memory impairment | 4229448 | Anterograde amnesia | 88822006 | S | Conditions | SNOMED |
| HP_0002354 | memory impairment | 4198081 | Retrograde amnesia | 51921000 | S | Conditions | SNOMED |
| HP_0002354 | memory impairment | 4012209 | Temporary loss of memory | 162200009 | S | Conditions | SNOMED |
| HP_0002354 | memory impairment | 4085496 | Poor long-term memory | 247588002 | S | Conditions | SNOMED |
| HP_0002354 | memory impairment | 4084412 | Poor short-term memory | 247592009 | S | Conditions | SNOMED |
| HP_0002354 | memory impairment | 372608 | Amnestic disorder | 3298001 | S | Conditions | SNOMED |
| HP_0002354 | memory impairment | 36712678 | Amnestic disorder associated with general medical condition | 1.05451E+14 | S | Conditions | SNOMED |
| HP_0002354 | memory impairment | 435795 | Fugue | 80490004 | S | Conditions | SNOMED |
| HP_0002354 | memory impairment | 4099961 | Mild memory disturbance | 192071009 | S | Conditions | SNOMED |
| HP_0002354 | memory impairment | 4155096 | Poor historian | 272054000 | S | Conditions | SNOMED |
| HP_0002354 | memory impairment | 444259 | Psychogenic amnesia | 84209002 | S | Conditions | SNOMED |
| HP_0002354 | memory impairment | 4145069 | Transient memory loss | 307413004 | S | Conditions | SNOMED |
| HP_0031987 | diminished ability to concentrate | 4096147 | Poor concentration | 26329005 | S | Conditions | SNOMED |
| HP_0031987 | diminished ability to concentrate | 4243064 | Unable to concentrate | 60032008 | S | Conditions | SNOMED |

Appendix A3: Qualifying Fatigue Symptoms and OMOP Codes

Table of qualifying Fatigue Long COVID symptoms

| phenotype_id | phenotype_label | omop_concept_id | omop_concept_label | omop_concept_code | omop_standard | omop_domain | omop_vocabulary |
| --- | --- | --- | --- | --- | --- | --- | --- |
| HP_0000716 | depressivity | 440383 | Depressive disorder | 35489007 | S | Conditions | SNOMED |
| HP_0000716 | depressivity | 433440 | Dysthymia | 78667006 | S | Conditions | SNOMED |
| HP_0000716 | depressivity | 4191716 | Symptoms of depression | 394924000 | S | Conditions | SNOMED |
| HP_0000716 | depressivity | 37016718 | Acute depression | 712823008 | S | Conditions | SNOMED |
| HP_0000716 | depressivity | 4103574 | Chronic depression | 192080009 | S | Conditions | SNOMED |
| HP_0000716 | depressivity | 40546087 | Depressed mood | 366979004 | S | Conditions | SNOMED |
| HP_0000716 | depressivity | 4149320 | Mild depression | 310495003 | S | Conditions | SNOMED |
| HP_0000716 | depressivity | 4336957 | Mild major depression | 87512008 | S | Conditions | SNOMED |
| HP_0000716 | depressivity | 4151170 | Moderate depression | 310496002 | S | Conditions | SNOMED |
| HP_0000716 | depressivity | 4307111 | Moderate major depression | 832007 | S | Conditions | SNOMED |
| HP_0000716 | depressivity | 36717092 | Moderately severe depression | 719593009 | S | Conditions | SNOMED |
| HP_0000716 | depressivity | 36714389 | Moderately severe major depression | 719592004 | S | Conditions | SNOMED |
| HP_0000716 | depressivity | 4149321 | Severe depression | 310497006 | S | Conditions | SNOMED |
| HP_0000716 | depressivity | 42872722 | Severe major depression | 450714000 | S | Conditions | SNOMED |
| HP_0000739 | anxiety | 441542 | Anxiety | 48694002 | S | Conditions | SNOMED |
| HP_0000739 | anxiety | 4087190 | Performance anxiety | 279622009 | S | Conditions | SNOMED |
| HP_0000739 | anxiety | 442077 | Anxiety disorder | 197480006 | S | Conditions | SNOMED |
| HP_0000739 | anxiety | 4199892 | Anxiety disorder due to a general medical condition | 52910006 | S | Conditions | SNOMED |
| HP_0000739 | anxiety | 381537 | Organic anxiety disorder | 17496003 | S | Conditions | SNOMED |
| HP_0000739 | anxiety | 4193634 | Worried | 79015004 | S | Conditions | SNOMED |
| HP_0000739 | anxiety | 440083 | Acute stress disorder | 67195008 | S | Conditions | SNOMED |
| HP_0000739 | anxiety | 4098314 | Chronic anxiety | 191708009 | S | Conditions | SNOMED |
| HP_0000739 | anxiety | 4138454 | Chronic stress disorder | 426174008 | S | Conditions | SNOMED |
| HP_0000739 | anxiety | 4239819 | Distress | 69328002 | S | Conditions | SNOMED |
| HP_0000739 | anxiety | 4142455 | Feeling hopeless | 307077003 | S | Conditions | SNOMED |
| HP_0000739 | anxiety | 436817 | Feeling nervous | 424196004 | S | Conditions | SNOMED |
| HP_0000739 | anxiety | 434613 | Generalized anxiety disorder | 21897009 | S | Conditions | SNOMED |
| HP_0000739 | anxiety | 4322025 | Mild anxiety | 70997004 | S | Conditions | SNOMED |
| HP_0000739 | anxiety | 4263429 | Moderate anxiety | 61387006 | S | Conditions | SNOMED |
| HP_0000739 | anxiety | 4103273 | Recurrent anxiety | 191709001 | S | Conditions | SNOMED |
| HP_0000739 | anxiety | 4214746 | Severe anxiety | 80583007 | S | Conditions | SNOMED |
| HP_0000739 | anxiety | 4216670 | Worried well | 81302005 | S | Conditions | SNOMED |
| HP_0002027 | abdominal pain | 4183041 | Abdominal discomfort | 43364001 | S | Conditions | SNOMED |
| HP_0002027 | abdominal pain | 200219 | Abdominal pain | 21522001 | S | Conditions | SNOMED |
| HP_0002027 | abdominal pain | 4152197 | Stomach ache | 271681002 | S | Conditions | SNOMED |
| HP_0002027 | abdominal pain | 4012224 | Upset stomach | 162059005 | S | Conditions | SNOMED |
| HP_0002027 | abdominal pain | 4200114 | Abdominal pain - cause unknown | 314212008 | S | Conditions | SNOMED |
| HP_0002027 | abdominal pain | 4263576 | Abdominal wind pain | 45979003 | S | Conditions | SNOMED |
| HP_0002027 | abdominal pain | 4002232 | Epigastric discomfort | 119416008 | S | Conditions | SNOMED |
| HP_0002027 | abdominal pain | 761102 | Intractable abdominal pain | 1.47E+16 | S | Conditions | SNOMED |
| HP_0002027 | abdominal pain | 4152353 | Loin pain | 271857006 | S | Conditions | SNOMED |
| HP_0002027 | abdominal pain | 4182562 | Lower abdominal pain | 54586004 | S | Conditions | SNOMED |
| HP_0002027 | abdominal pain | 4306292 | Upper abdominal pain | 83132003 | S | Conditions | SNOMED |
| HP_0002027 | abdominal pain | 4203705 | Visceral abdominal pain | 438506002 | S | Conditions | SNOMED |
| HP_0002027 | abdominal pain | 4241033 | Acute abdomen | 9209005 | S | Conditions | SNOMED |
| HP_0002027 | abdominal pain | 42710018 | Functional abdominal pain syndrome | 449890002 | S | Conditions | SNOMED |
| HP_0002027 | abdominal pain | 4012222 | Central abdominal pain | 162046002 | S | Conditions | SNOMED |
| HP_0002027 | abdominal pain | 4008102 | Chronic abdominal pain | 111985007 | S | Conditions | SNOMED |
| HP_0002027 | abdominal pain | 4218793 | Colicky pain | 73063007 | S | Conditions | SNOMED |
| HP_0002027 | abdominal pain | 201418 | Flatulence, eructation and gas pain | 271832001 | S | Conditions | SNOMED |
| HP_0002027 | abdominal pain | 197988 | Generalized abdominal pain | 102614006 | S | Conditions | SNOMED |
| HP_0002027 | abdominal pain | 193317 | Generalized abdominal rigidity | 440122009 | S | Conditions | SNOMED |
| HP_0002027 | abdominal pain | 443629 | Generalized abdominal tenderness | 440220007 | S | Conditions | SNOMED |
| HP_0002027 | abdominal pain | 195083 | Left lower quadrant pain | 301716002 | S | Conditions | SNOMED |
| HP_0002027 | abdominal pain | 4109084 | Left sided abdominal pain | 285387005 | S | Conditions | SNOMED |
| HP_0002027 | abdominal pain | 194175 | Left upper quadrant pain | 301715003 | S | Conditions | SNOMED |
| HP_0002027 | abdominal pain | 4128083 | Nonspecific abdominal pain | 304542004 | S | Conditions | SNOMED |
| HP_0002027 | abdominal pain | 4115547 | Rebound tenderness of left hypochondrium | 301414000 | S | Conditions | SNOMED |
| HP_0002027 | abdominal pain | 4118277 | Rebound tenderness of right hypochondrium | 301413006 | S | Conditions | SNOMED |
| HP_0002027 | abdominal pain | 4258543 | Recurrent abdominal pain | 439469002 | S | Conditions | SNOMED |
| HP_0002027 | abdominal pain | 193322 | Right lower quadrant pain | 301754002 | S | Conditions | SNOMED |
| HP_0002027 | abdominal pain | 4109085 | Right sided abdominal pain | 285388000 | S | Conditions | SNOMED |
| HP_0002027 | abdominal pain | 198263 | Right upper quadrant pain | 301717006 | S | Conditions | SNOMED |
| HP_0002355 | difficulty walking | 36714126 | Difficulty walking | 719232003 | S | Conditions | SNOMED |
| HP_0002355 | difficulty walking | 439405 | Walking disability | 228158008 | S | Conditions | SNOMED |
| HP_0002355 | difficulty walking | 4107172 | Difficulty in starting and stopping walking spontaneously | 282220004 | S | Conditions | SNOMED |
| HP_0002355 | difficulty walking | 4107168 | Difficulty in stopping walking | 282213006 | S | Conditions | SNOMED |
| HP_0002355 | difficulty walking | 4084106 | Difficulty initiating walking | 282207002 | S | Conditions | SNOMED |
| HP_0002355 | difficulty walking | 4107166 | Does not initiate walking | 282206006 | S | Conditions | SNOMED |
| HP_0002355 | difficulty walking | 4116707 | Unable to weight-bear | 285674009 | S | Conditions | SNOMED |
| HP_0003546 | exercise intolerance | 4147189 | Impaired exercise tolerance | 267044007 | S | Conditions | SNOMED |
| HP_0003546 | exercise intolerance | 4299640 | Activity intolerance | 77427003 | S | Conditions | SNOMED |
| HP_0003546 | exercise intolerance | 4000163 | Cough on exercise | 19282004 | S | Conditions | SNOMED |
| HP_0012378 | fatigue | 4223659 | Fatigue | 84229001 | S | Conditions | SNOMED |
| HP_0012378 | fatigue | 4309912 | Generally unwell | 213257006 | S | Conditions | SNOMED |
| HP_0012378 | fatigue | 4087481 | Lack of energy | 248274002 | S | Conditions | SNOMED |
| HP_0012378 | fatigue | 4272240 | Malaise | 367391008 | S | Conditions | SNOMED |
| HP_0012378 | fatigue | 4074624 | Tired | 224960004 | S | Conditions | SNOMED |
| HP_0012378 | fatigue | 4086975 | Undifferentiated illness: Vague ill health | 248282002 | S | Conditions | SNOMED |
| HP_0012378 | fatigue | 4152509 | C/O - "tired all the time" | 272062008 | S | Conditions | SNOMED |
| HP_0012378 | fatigue | 4060222 | Chronic sick | 161901003 | S | Conditions | SNOMED |
| HP_0012378 | fatigue | 4158498 | Fatigue - symptom | 272060000 | S | Conditions | SNOMED |
| HP_0012378 | fatigue | 4198140 | Feeling tired | 314109004 | S | Conditions | SNOMED |
| HP_0012378 | fatigue | 4060217 | Heavy feeling | 161874006 | S | Conditions | SNOMED |
| HP_0012378 | fatigue | 439926 | Malaise and fatigue | 271795006 | S | Conditions | SNOMED |
| HP_0012378 | fatigue | 37017316 | Occasionally tired | 713568000 | S | Conditions | SNOMED |
| HP_0012378 | fatigue | 4092860 | Rapid fatigue of gait | 250002000 | S | Conditions | SNOMED |
| HP_0012378 | fatigue | 4149857 | Tired all the time | 267032009 | S | Conditions | SNOMED |
| HP_0012378 | fatigue | 4086973 | Tired on least exertion | 248269005 | S | Conditions | SNOMED |
| HP_0012378 | fatigue | 4147184 | Tiredness symptom | 267031002 | S | Conditions | SNOMED |
| HP_0012378 | fatigue | 4246495 | Exhaustion | 60119000 | S | Conditions | SNOMED |

Appendix A4: Qualifying Respiratory Symptoms and OMOP Codes

Table of qualifying Respiratory Long COVID symptoms

| phenotype_id | phenotype_label | omop_concept_id | omop_concept_label | omop_concept_code | omop_standard | omop_domain | omop_vocabulary |
| --- | --- | --- | --- | --- | --- | --- | --- |
| HP_0002094 | dyspnea | 4305080 | Abnormal breathing | 386813002 | S | Conditions | SNOMED |
| HP_0002094 | dyspnea | 312437 | Dyspnea | 267036007 | S | Conditions | SNOMED |
| HP_0002094 | dyspnea | 4060052 | Dyspnea at rest | 161941007 | S | Conditions | SNOMED |
| HP_0002094 | dyspnea | 43020567 | Air leaking from lung | 471283005 | S | Conditions | SNOMED |
| HP_0002094 | dyspnea | 4094132 | Nocturnal dyspnea | 248548009 | S | Conditions | SNOMED |
| HP_0002110 | bronchiectasis | 256449 | Bronchiectasis | 12295008 | S | Conditions | SNOMED |
| HP_0002110 | bronchiectasis | 4052549 | Acquired bronchiectasis | 233628009 | S | Conditions | SNOMED |
| HP_0002110 | bronchiectasis | 4052550 | Idiopathic bronchiectasis | 233629001 | S | Conditions | SNOMED |
| HP_0002110 | bronchiectasis | 40483342 | Acute exacerbation of bronchiectasis | 445378003 | S | Conditions | SNOMED |
| HP_0002110 | bronchiectasis | 4027836 | Fusiform bronchiectasis | 13217005 | S | Conditions | SNOMED |
| HP_0002875 | exertional dyspnea | 4263848 | Dyspnea on exertion | 60845006 | S | Conditions | SNOMED |
| HP_0006530 | abnormal pulmonary interstitial morphology | 4119786 | Interstitial lung disease | 233703007 | S | Conditions | SNOMED |
| HP_0006530 | abnormal pulmonary interstitial morphology | 438791 | Pulmonary congestion and hypostasis | 196115007 | S | Conditions | SNOMED |
| HP_0006530 | abnormal pulmonary interstitial morphology | 45768996 | Pulmonary interstitial glycogenosis | 707551007 | S | Conditions | SNOMED |
| HP_0006530 | abnormal pulmonary interstitial morphology | 762964 | Chronic interstitial lung disease | 4.34301E+14 | S | Conditions | SNOMED |
| HP_0006530 | abnormal pulmonary interstitial morphology | 4045227 | Respiratory bronchiolitis associated interstitial lung disease | 129451001 | S | Conditions | SNOMED |
| HP_0012735 | cough | 4158493 | C/O - cough | 272039006 | S | Conditions | SNOMED |
| HP_0012735 | cough | 254761 | Cough | 49727002 | S | Conditions | SNOMED |
| HP_0012735 | cough | 4195384 | Chronic cough | 68154008 | S | Conditions | SNOMED |
| HP_0012735 | cough | 4086814 | Clearing throat - hawking | 248589007 | S | Conditions | SNOMED |
| HP_0012735 | cough | 4048098 | Cough with fever | 135883003 | S | Conditions | SNOMED |
| HP_0012735 | cough | 4269800 | Nocturnal cough | 62548007 | S | Conditions | SNOMED |
| HP_0012735 | cough | 4144596 | Pain provoked by coughing | 427146008 | S | Conditions | SNOMED |
| HP_0012735 | cough | 4090569 | Painful cough | 247410004 | S | Conditions | SNOMED |
| HP_0012735 | cough | 4109381 | Persistent cough | 284523002 | S | Conditions | SNOMED |
| HP_0012735 | cough | 4099940 | Psychogenic cough | 191954008 | S | Conditions | SNOMED |
| HP_0025179 | Ground-glass opacification on pulmonary HRCT | 4318404 | Lung consolidation | 95436008 | S | Conditions | SNOMED |
| HP_0025179 | Ground-glass opacification on pulmonary HRCT | 4306177 | On examination - lung consolidation | 419449001 | S | Conditions | SNOMED |
| HP_0025390 | Reticular pattern on pulmonary HRCT | 4318404 | Lung consolidation | 95436008 | S | Conditions | SNOMED |
| HP_0025390 | Reticular pattern on pulmonary HRCT | 4306177 | On examination - lung consolidation | 419449001 | S | Conditions | SNOMED |
| HP_0030877 | Reduced FEV1/FVC ratio | 4064741 | FEV1/FVC ratio abnormal | 165044007 | S | Conditions | SNOMED |
| HP_0030879 | Interlobular septal thickening on pulmonary HRCT | 4318404 | Lung consolidation | 95436008 | S | Conditions | SNOMED |
| HP_0030879 | Interlobular septal thickening on pulmonary HRCT | 4306177 | On examination - lung consolidation | 419449001 | S | Conditions | SNOMED |
| HP_0031245 | productive cough | 4102774 | Productive cough | 28743005 | S | Conditions | SNOMED |
| HP_0031246 | nonproductive cough | 4038519 | Dry cough | 11833005 | S | Conditions | SNOMED |
| HP_0031983 | Abnormal pulmonary thoracic imaging finding | 4318404 | Lung consolidation | 95436008 | S | Conditions | SNOMED |
| HP_0031983 | Abnormal pulmonary thoracic imaging finding | 4306177 | On examination - lung consolidation | 419449001 | S | Conditions | SNOMED |
| HP_0032177 | Parenchymal consolidation | 4318404 | Lung consolidation | 95436008 | S | Conditions | SNOMED |
| HP_0032177 | Parenchymal consolidation | 4306177 | On examination - lung consolidation | 419449001 | S | Conditions | SNOMED |
| HP_0032341 | Reduced forced vital capacity | 4065408 | FVC - forced vital capacity abnormal | 165040003 | S | Conditions | SNOMED |
| HP_0032342 | Reduced forced expiratory volume in one second | 4151447 | FEV1/FVC < 70% of predicted | 314472003 | S | Conditions | SNOMED |
| HP_0033106 | Elevated D-dimers | 42709958 | D-dimer above reference range | 449830004 | S | Conditions | SNOMED |
| HP_0033169 | Reduced total lung capacity | 4267216 | Decreased total lung capacity | 36317006 | S | Conditions | SNOMED |
| HP_0045051 | Decreased DLCO | 4142621 | Decreased diffusion capacity of lung | 34627006 | S | Conditions | SNOMED |
